## Supplementary Material for "Estimating the effects of non-pharmaceutical interventions on the number of new infections with COVID-19 during the first epidemic wave"

**Abstract**

The supplementary material contains (i) a detailed description of the method, (ii) further descriptives, (iii) detailed estimation results and model checks, (iv) results from the sensitivity analysis, (v) a visual inspection of the model fit, and (vi) data on non-pharmaceutical interventions.

References 128

#### List of Tables

1 Start and end of non-modeling and modeling phase by country. . . . . 14
2 Prior choices for model parameters. . . . . 15

3 Distance (in days) to the next NPIs in time per contry and NPI. “Average” is the mean distance over all countries. Computation: For each NPI, the distance is measured as the absolute difference (in days) to the NPI that was implemented closest before or after, which is then averaged across countries. For countries where NPIs were implemented at the regional level, the absolute differences were first averaged over all regions. If an NPI was not implemented, then the absolute difference is omitted from computation. . . . . 17

4 Pairwise average distance (in days) between the implementation of NPIs across countries. Computation: For each pair of NPIs, the distance is measured as the absolute difference (in days) between the implementation dates of NPIs, which is then averaged across countries. For countries where NPIs were implemented at the regional level, the absolute differences were first averaged over all regions. If an NPI was not implemented, then the absolute difference is omitted from computation. . . . . 18

5 Estimation results . . . . . 20

6 Sources for policies in Switzerland, Austria, Belgium, Denmark, Finland, France, United Kingdom, Greece, Ireland, Luxembourg, Netherlands, Norway, Portugal, Sweden, United States of America, Germany, Italy, Spain, Canada, and Australia . . . . 57

7 Sources for policies implemented across different US States . . . . . 80

8 Sources for policies implemented across different German regions . . . . . 86

9 Sources for policies implemented across different Spanish regions . . . . . 97

10 Sources for policies implemented across different Canadian regions . . . . . 102

11 Sources for policies implemented across different Australian regions . . . . . 106

12 Sources for policies implemented across different Italian regions . . . . . 116

13 Sources for policies implemented across different Swiss regions . . . . . 127

### 1 Method

#### 1.1 Notation

| | |
|---|---|
| $j$ | country |
| $t$ | days since start of observation period |
| $N_{jt}$ | number of reported new cases in country $j$ at day $t$ |
| $I_{jt}$ | number of new infections (transmissions) in country $j$ at day $t$ (unobserved) |
| $C_{jt}$ | number of contagious subjects in country $j$ at day $t$ (unobserved) |

To model the impact of a non-pharmaceutical intervention (NPI) in a specific country, we have to take into account that some countries introduced the NPIs only in specific subregions or at different timepoints in different subregions. Hence, with respect to the NPIs, we introduce the following notation:

| | |
|---|---|
| $m$ | NPI (numbered from 1 to $M$) |
| $r$ | subregion $r$ of country $j$ ($r = 1,...,R_j$) |
| $T_{mrjt}$ | number of days since NPI $m$ took effect in region $r$ of country $j$ at day $t$ (counting the first day at which the NPI could affect the number of new cases as $t = 1$) |
| $p_{rj}$ | the share of region's $r$ population of the total population of country $j$ |

Note that, also in countries with several regions, only the overall number of new cases is analyzed, not region specific counts.

#### 1.2 Overall approach

The overall aim is to assess the impact of NPI $m$ on the number of new infections in the days after the NPI becomes active. We approach this by modeling the number of new infections as a function of the number of contagious subjects and the presence of active measures. That is, we consider a model linking two unobserved quantities. To obtain a model in the observed quantities $N_{jt}$, we link this number to the number of new infections in the previous days. Similarly, we link the number of contagious subjects to the number of new infections in the previous days. The overall model is then fitted using a fully Bayesian approach, which requires the specification of prior distributions for all model parameters.

In the following, we present the three submodels and describe to which degree external knowledge is incorporated to justify choices.

##### 1.3 Relating the number of new infections to the number of contagious subjects and the presence of NPIs

Let $\theta_m$ denote the relative reduction of new infections when NPI $m$ is fully implemented, i.e. the fraction of avoided new infections compared to the situation without this NPI. Let $f(t)$ denote a function taking values between 0 and 1 describing the degree of implementation as a function of the time since start of the NPI. In a country $j$ with only one region, $\theta_m f(T_{m1jt})$ describes the fraction of new infections that are avoided in this country at day $t$. If regions are exposed to NPIs to a varying degree, the corresponding fraction is $\theta_m \sum_{r=1}^{R_j} p_{rj}\, f(T_{mrjt})$.

The overall fraction of *un*-avoided new infections is then given by

$$\prod_{m=1}^{M}\left(1-\theta_m \sum_{r=1}^{R_j} p_{rj}\, f(T_{mrjt})\right). \tag{1}$$

In the absence of any measure, the expected value of the number of new infections $\mu^{I_{jt}}$ would be determined by the number of contagious subjects $C_{jt}$ and the country-specific daily transmission rate $\delta_j$, i.e., $\mu^{I_{jt}} = C_{jt}\ \delta_j$. In the presence of NPIs, we have to multiply this with the fraction of un-avoided infections, resulting in

$$\mu^{I_{jt}} = C_{jt}\ \delta_j \prod_{m=1}^{M}\left(1-\theta_m \sum_{r=1}^{R_j} p_{rj}\, f(T_{mrjt})\right). \tag{2}$$

It would be natural to model the number of new infections as a negative binomial distribution. However, the software used in this work does not allow integer values for unobserved variables. Therefore, a normal distribution was used instead, with the mean and standard deviation analogous to a negative binomial distribution:

$$I_{jt} \sim \mathrm{Log-normal}\left(\mu^{I_{jt}}, \sigma^{I_{jt}}\right)\ , \tag{3}$$

with $\sigma^{I_{jt}} = \sqrt{\mu^{I_{jt}} \left(1 + \frac{\mu^{I_{jt}}}{\phi^I}\right)}$, and an overdispersion parameter $\phi^I$.

##### 1.4 Relating the number of observed cases to the number of new infections in the previous days

The expected number $\mu^{N_{jt}}$ of new cases $N_{jt}$ in country $j$ at day $t$ can be derived from the number of new infections in the previous days as

$$\mu^{N_{jt}} = \sum_{s<t} I_{js} \cdot p_{\text{IN}}(t-s) \ , \tag{4}$$

where $p_{\text{IN}}(t)$ denotes the probability that a new infected subject is reported at day $t$ after the infection, i.e., becomes a new case. It is assumed that $p_{\text{IN}}(t)$ is a discretized version of a log-normal distribution. This distribution is estimated from our data as part of fitting the overall model using a weakly informative prior, as described later.

The observed number of new cases are modeled to follow a negative binomial distribution (NB), i.e.,

$$N_{jt} \sim \text{NB}\left(\mu^{N_{jt}}, \sigma^{N_{jt}}\right) \tag{5}$$

with mean $\mu^{N_{jt}}$, standard deviation $\sigma^{N_{jt}} = \sqrt{\mu^{N_{jt}} \left(1 + \frac{\mu^{N_{jt}}}{\phi^N}\right)}$, and an overdispersion parameter $\phi^N$.

##### 1.5 Relating the number of contagious subjects to the number of new infections in the previous days

The expected number $\mu^{C_{jt}}$ of contagious subjects in country $j$ at day $t$ can be derived from the number of new infections in the previous days as

$$\mu^{C_{jt}} = \sum_{s<t} I_{js} p_{\text{IC}}(t-s) \ , \tag{6}$$

where $p_{\text{IC}}(t)$ denotes the probability that a new infected subject is contagious at day $t$ after the infection. Since our data does not include information about these probabilities, we make a choice based on external information and considerations presented in the next section. Moreover, we regard the relation between $I_{jt}$ and $C_{jt}$ as a deterministic one, i.e., $C_{jt} = \mu^{C_{jt}}$.

##### 1.6 Relating the probability to be contagious to the generation time distribution

Let $\gamma$ denote the probability that a randomly chosen contagious subject infects another randomly chosen subject within one day. Considering the probabilities $q(t)$ that an infected subject infects another randomly chosen subject at day $t$ after his/her own infection, there is the simple relation

$$q(t) = p_{\mathrm{IC}}(t)\,\gamma\ . \tag{7}$$

Now, let $p_{\mathrm{G}}(t)$ denote the density of the generation time distribution. That is, given a subject has infected another subject, $p_{\mathrm{G}}(t)$ is the probability that this has happened $t$ days after his/her infection. It follows that there is a simple relation

$$p_{\mathrm{G}}(t) = \frac{q(t)}{\sum_t q(t)} = \frac{p_{\mathrm{IC}}(t)\,\gamma}{\sum_t p_{\mathrm{IC}}(t)\,\gamma} = \frac{p_{\mathrm{IC}}(t)}{\sum_t p_{\mathrm{IC}}(t)} \tag{8}$$

showing that $p_{IC}(t)$ is proportional to the generation time distribution. Consequently, an estimate for the generation time distribution is needed. The proportionality factor can be omitted because $\mu^{I_{jt}}$ is multiplicative in $C_{jt}$, and thus the omitted factor can be subsumed into $\delta_j$.

Approximating $p_{\mathrm{IC}}$ by the generation time distribution has been also done by Cori et al[1]. A more rigorous discussion about the validity of this approximation is given by Fraser et al[2]. Since we model the ratio between the expected number of new infections and the number of contagious subjects, our approach can be also interpreted as modeling the course of the reproduction number over time.

##### 1.7 Choice of priors for the parameters of the distribution of the time from infection to reporting

The time from infection to reporting of a new case can be written as a sum of the incubation period and the time from symptom onset to reporting (i.e., the reporting delay). Estimates for these individual distributions can be found in the literature. A rapid systematic review and meta analysis of observational research on the incubation period[3] arrived at a Log-normal distribution for the incubation period and reported estimates of $\mu = 1.63$ for the mean and of $\sigma = 0.50$ for the standard deviation of the natural logarithm. This translates into an average incubation period of about six days. A study on the COVID-19 outbreak in Italy[4] arrived at a Gamma distribution for the reporting delay and reported estimates of $\alpha = 1.88$ for the shape and $\beta = 0.26$ for the inverse scale. This translates into an average reporting delay of about seven days, which is in line with estimates from other countries[5–8].

It is reasonable to assume that the incubation period and the reporting delay are independent. Following this assumption, the distribution for the time from infection to reporting of a new case is the sum of i.i.d. random variables and thus we can compute the mean and variance as the sum of the individual means and variances. Assuming for the sum again a log-normal distribution hence leads to the choice of a $\mathrm{Log-normal}(\mu = 2.47, \sigma = 0.45)$. This translates into an average delay for the time from infection to reporting of a new case of about thirteen days with a standard deviation of six days. However, a fixed choice for $p_{\mathrm{IN}}$ would neglect the uncertainty about the shape of the distribution and thus the parameters of the Log-normal distribution were instead estimated from our data as part of fitting the overall model. To do so, weakly informative priors are chosen for the mean $\mu \sim \mathrm{Normal}(2.47, 0.50)$ and standard deviation $\sigma \sim \mathrm{Gamma}(2.00, 4.48)$ (since $\frac{2.00}{4.448} \approx 0.45$) of the natural logarithm. Our choices reflect prior knowledge about the delay's mean and standard deviation, while taking uncertainty about their estimates into account.

The prior distributions for $\mu$ and $\sigma$ as well as the resulting prior distributions for the discrete probabilities $p_{\mathrm{IN}}(t)$ are shown in Fig. 1. Note that $p_{\mathrm{IN}}$ is discretized via $p_{\mathrm{IN}}(s) = \int_0^{0.5} p_{\mathrm{IN}}(\tau)\, \mathrm{d}\tau$ for $s = 0$ and $p_{\mathrm{IN}}(s) = \int_{s-0.5}^{s+0.5} p_{\mathrm{IN}}(\tau)\, \mathrm{d}\tau$ for $s = 1, 2, \ldots$, where $p_{\mathrm{IN}}(\tau) \sim \mathrm{Log-normal}(\mu, \sigma)$ is the density of the Log-normal distribution with mean $\mu$ and standard deviation $\sigma$.

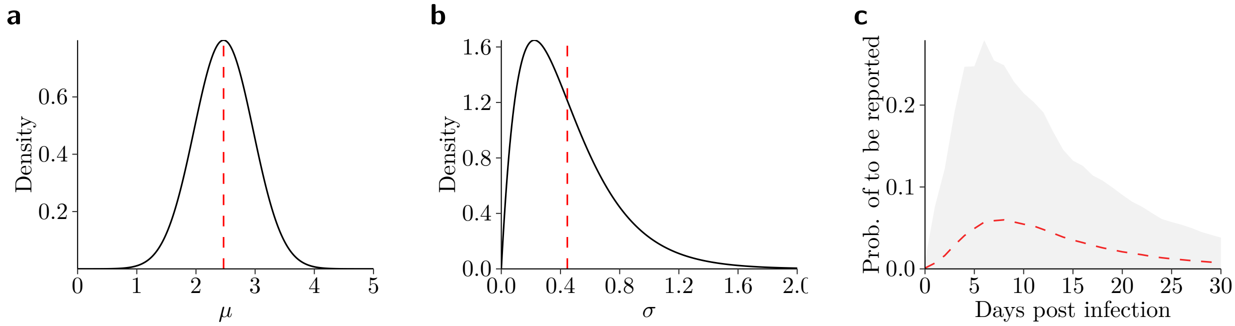

**Figure 1.** Prior choice for the distribution of the time from infection to reporting of a new case. **(a)** Log mean $\mu$ (prior mean as dashed red line). **(b)** Log standard deviation $\sigma$ (prior mean as dashed red line). **(c)** Distribution of $p_{\mathrm{IN}}(t)$ for $t = 0, 1, \ldots, 30$ (prior mean as dashed red line with 95 % range as shaded area, based on 4,000 independent draws from the distributions for $\mu$ and $\sigma$).

#### 1.8 Choice of estimate for the generation time distribution

The generation time distribution is often approximated by the serial interval distribution (e.g.,[9]). However, it was recently pointed out that this approximation is based on the assumption that the incubation period and the infectiousness profile are independent[10], which is questionable. Other studies estimated the generation time distribution from data on transmission pairs[11,12], thereby making the implicit assumption that the generation time is independent of the incubation period[10], which is also questionable.

By leveraging data on the exposure for both index and secondary cases, a recent study inferred the generation distribution more accurately[13] without having to make the above assumptions. This study suggests to use a Weibull(3.28, 6.12) distribution, which has a mean of 5.49 days and a standard deviation of 1.84 days.

The discrete form of this distribution is depicted in Fig. 2. Again, the distribution is discretized via $p_\mathrm{G}(s) = \int_0^{1.5} p_\mathrm{G}(\tau)\,\mathrm{d}\tau$ for $s = 1$ and $p_\mathrm{G}(s) = \int_{s-0.5}^{s+0.5} p_\mathrm{G}(\tau)\,\mathrm{d}\tau$ for $s = 2, 3, \ldots$, where $p_\mathrm{G}(\tau) \sim$ Weibull(3.28, 6.12) is the density of the Weibull distribution with shape $\alpha$ and scale $\kappa$. Note that we explicitly set $p_\mathrm{IC}(0)$ to 0, avoiding the challenge to include new infections at one day in the number of contagious subjects the same day.

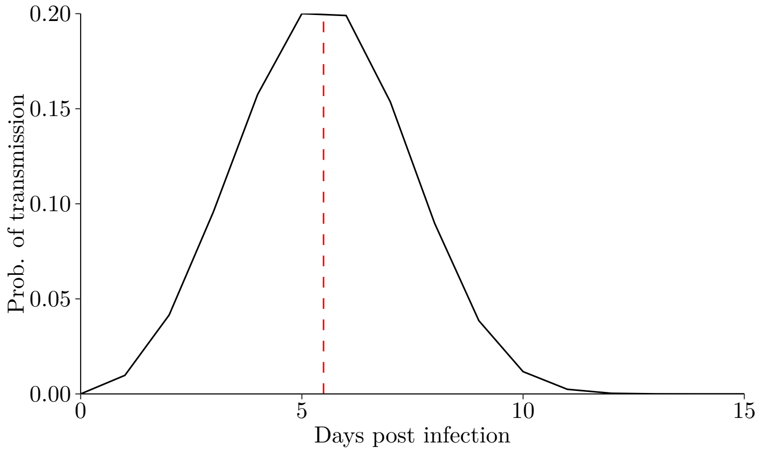

**Figure 2.** Prior choice for the generation time distribution $p_\mathrm{G}(t)$.

#### 1.9 Choice of the functional form of the time-delayed response function

We model the time-delayed response function with a first-order spline

$$f(t) = \begin{cases} 0, & \text{if } t \le t_0, \\ \frac{t-t_0}{t_1-t_0}, & \text{if } t_0 < t < t_1, \\ 1, & \text{if } t \ge t_1. \end{cases} \tag{9}$$

This means that the effect of an NPI increases linearly after its implementation until $t_1$. We set $t_0 = 0$ days and $t_1 = 3$ days, reflecting a three day period until people fully respond to the implemented NPI.

#### 1.10 Modeling and non-modeling phase

It took some time for countries to set up reporting practices so that case numbers at the very beginning of the epidemic are often missing or irregular. That is why, in each country, modeling of the number of new cases starts after 100 cumulative cases were reported, i.e., the first day at which the number of cumulative cases exceeds 100 is set as $t = 1$. Before that, the number of new cases is seeded similar to Ref.[9]. That is, in the non-modeling phase, the modeling of $N_{jt}$ is ignored, but the other components are still used and, in particular, $\mu^{I_{jt}}$ and $\mu^{C_{jt}}$ are computed. The start of the non-modeling phase is determined as the day at which we would expect the first case when reaching 100 cumulative cases at day $t = 1$, given the reproduction number, mean generation time and reporting delay as specified. That is,

$$t = 1 - \frac{\log 100}{\log 3.28} \cdot 5.49 + 13.01 \approx -33, \tag{10}$$

where $R_0 = 3.28$ is the basic reproductive number taken from a meta analysis across countries[14], 5.49 days is the mean generation time and 13.01 days is the mean time from infection to reporting. For $I_{j\,-33}$, we replace the recursive formula of our model with

$$I_{j\,-33} \sim \text{Exponential}\left(\frac{1}{\lambda}\right) \tag{11}$$

and choose $\lambda \sim \text{Exponential}(1)$ as a prior distribution. Thereby, it is expected that one person is infected at day $t = -33$, but the estimated number can vary substantially by country. Using the recursive formula, the number of contagious subjects at the start of the non-modeling phase are computed from the initially infected seven days before the non-modeling phase. The initially infected are computed by assuming an exponential growth rate of $\frac{\log 3.28}{5.49}$, i.e.,

$$I_{jt} = I_{j\,-33} \cdot \exp\left(\frac{\log 3.28}{5.49} \cdot (-33 - t)\right),\ t = -40, -39, \ldots, -34.$$

Note that the initially infected are only computed to smooth the number of infected and contagious subjects at the start of the non-modeling phase.

The end of the modeling phase is, in each country, set to 28 days after the last NPI was implemented in any of the country's regions. This provides sufficient time for the effects of NPIs to show up in the number of new cases, thereby concluding the first wave of the epidemic, as most countries saw their case numbers reverting back after all NPIs were in place. The start of the non-modeling phase as well as the start and end of the modeling phase are shown in Tbl. 1. We made only two adjustments. In Australia, Queensland closed schools on April 20, while all other regions implemented their NPIs before April 2, thus we set April 2 plus 28 days for the end of the modeling phase in Australia. Similarly, in Canada, Northwest Territories canceled gatherings on April 11 while all other territories implemented their NPIs before April 1, thus we set April 1 plus 28 days for the end of the modeling phase in Canada.

##### 1.11 Choice of prior distributions for the effect of NPIs

Our primary goal is to infer the effect of NPIs. These policy measures were implemented to reduce new infections via social distancing. We wanted to construct a prior for the effect of NPIs that (1) has support $\theta_m \in (-\infty, 1)$, thereby not precluding a negative effect corresponding to an increase in new infections from NPIs but also allowing a single NPI to account for a complete elimination of transmissions, (2) gives higher probability to positive effects than to negative effects , and (3) is little informative with respect to the magnitude of a positive effect.

The desired properties were accomplished with a mixture of a half normal distribution for negative effects and a uniform distribution for positive effects. The formal specification

$$\theta_m \sim \text{Mixture}(w) = \begin{cases} \text{Normal}^{-}(0, \sigma) & \text{with probability } w, \\ \text{Uniform}(0, 1) & \text{with probability } (1 - w). \end{cases} \tag{12}$$

| Country | Non-modeling phase Start | Modeling phase Start | End |
|---|---|---|---|
| Australia | Feb 11 | Mar 10 | Apr 30 |
| Austria | Feb 09 | Mar 08 | Apr 16 |
| Belgium | Feb 07 | Mar 06 | Apr 17 |
| Canada | Feb 12 | Mar 11 | Apr 29 |
| Denmark | Feb 11 | Mar 10 | Apr 15 |
| Finland | Feb 14 | Mar 13 | Apr 16 |
| France | Feb 01 | Feb 29 | Apr 14 |
| Germany | Feb 02 | Mar 01 | Apr 20 |
| Greece | Feb 14 | Mar 13 | Apr 20 |
| Ireland | Feb 15 | Mar 14 | Apr 25 |
| Italy | Jan 26 | Feb 23 | Apr 23 |
| Luxembourg | Feb 18 | Mar 17 | Apr 15 |
| Netherlands | Feb 07 | Mar 06 | Apr 20 |
| Norway | Feb 07 | Mar 06 | Apr 13 |
| Portugal | Feb 14 | Mar 13 | Apr 19 |
| Spain | Feb 03 | Mar 02 | Apr 27 |
| Sweden | Feb 07 | Mar 06 | Apr 24 |
| Switzerland | Feb 06 | Mar 05 | Apr 22 |
| United Kingdom | Feb 03 | Mar 02 | Apr 20 |
| United States | Feb 05 | Mar 04 | May 05 |

**Table 1.** Start and end of non-modeling and modeling phase by country.

where $w \in [0,1]$ is the mixing ratio. For efficient sampling, the prior density should rather be continuous and thus $\sigma$ is chosen based on $w$ such that this is the case, i. e.,

$$\sigma = \frac{w}{(1-w)\sqrt{2\pi}} \,. \tag{13}$$

As a default, a mixing ratio of $w = 0.1$ is chosen, which implies a 10% probability that NPIs can lead to an increase in new infections, resulting in $\sigma = 0.04$. The density of this prior is shown in Fig 3.

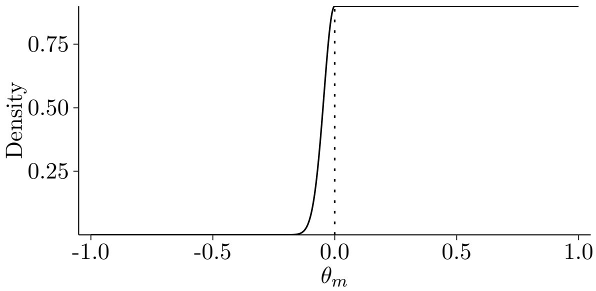

**Figure 3.** Prior choice for the effects of non-pharmaceutical interventions $\theta_m$.

| Parameter | Notation | (Hyper-)Prior |
|---|---|---|
| NPIs | $\theta_m$ | $\text{Mixture}(0.1) = \begin{cases} \text{Normal}^{-}(0, 0.04) & \text{with prob. } 0.1 \\ \text{Uniform}(0, 1) & \text{with prob. } 0.9 \end{cases}$ |
| Country-specific daily transmission rate | $\delta_j$ | $\delta_j = \exp(\alpha + \alpha_j)$ |
| | $\alpha$ | $\text{Student}-\text{t}(\nu = 7, \mu = 0, \sigma = 10)$ |
| | $\alpha_j$ | $\text{Normal}(0, \tau)$ |
| | $\tau$ | $\text{Student}-\text{t}^{+}(\nu = 4, \mu = 0, \sigma = 1)$ |
| Overdispersion | $\phi^N$ | $\phi^N = \left(\frac{1}{\xi^N}\right)^2$ |
| | $\xi^N$ | $\text{Normal}^{+}(\mu = 0, \sigma = 1)$ |
| | $\phi^I$ | $\phi^I = \left(\frac{1}{\xi^I}\right)^2$ |
| | $\xi^I$ | $\text{Normal}^{+}(\mu = 0, \sigma = 1)$ |
| Time from infection to new case | $p_{\text{IN}}$ | $\text{Log}-\text{normal}(\mu, \sigma)$ |
| | $\mu$ | $\text{Normal}(\mu = 2.47, \sigma = 0.45)$ |
| | $\sigma$ | $\text{Gamma}(\alpha = 2.00, \beta = 4.48)$ |
| Generation time | $p_{\text{G}}$ | $\text{Weibull}(\alpha = 3.28, \kappa = 6.12)$ |
| Initially infected subjects | $I_{j-33}$ | $\text{Exponential}\left(\frac{1}{\lambda}\right)$ |
| | $\lambda$ | $\text{Exponential}(1)$ |

**Table 2.** Prior choices for model parameters.

#### 1.12 Choice of prior distributions (summary)

Tbl. 2 provides an overview of the model parameters together with the choice of priors. If not tailored to the specifics of our model, the choice of priors are informed by recommendations on the choice of priors from the Stan Development Team[15].

#### 1.13 Model parameter estimation

Model parameters are estimated with a Bayesian approach. Specifically, Markov chain Monte Carlo (MCMC) sampling is used as implemented by the Hamiltonian Monte Carlo algorithm with the No-U-Turn Sampler (NUTS) from Stan 2.19.2[16]. If not stated otherwise, we report posterior means and credible intervals (CrIs) based on the 2.5% and 97.5% quantile of the posterior samples.

Each model is estimated with 4 Markov chains and 2,000 iterations of which the first 1,000 iterations are discarded as part of the warm-up. Estimation power is evaluated via the ratio of the effective sample size ($\hat{n}_{\text{eff}}/N$), and convergence of the Markov chains is assessed with the Gelman-Rubin convergence diagnostic ($\hat{R}$). Further checks pertain to the detection of influential observations and correlations between the parameters of interest.

#### 1.14 Ignoring undetected infections

In our considerations, we ignore that probably many infected subjects remain undetected as we implicitly assume that all infected case are detected ($p_{\text{IN}}(t)$ sums up to 1). Formally, we could try to take this into account by introducing the probability of an infected case to be observed. However, this would act mainly on the global intercept $\alpha'$ (except that the shape of a negative binomial distribution depends slightly on the actual sample size). Hence it seems to be safe to ignore this.

#### 2 Timing of non-pharmaceutical interventions

| | Event ban | School closure | Venue closure | Gathering ban | Border closure | Stay-at-home order | Work ban |
|---:|---:|---:|---:|---:|---:|---:|---:|
| Australia | 4.0 | 7.0 | 3.0 | 4.1 | 3.0 | 3.2 | |
| Austria | 5.0 | 3.0 | 3.0 | 3.0 | 3.0 | 3.0 | |
| Belgium | 4.0 | 2.0 | 2.0 | 2.0 | 2.0 | 2.0 | |
| Canada | 2.9 | 2.5 | 2.5 | 5.0 | 2.5 | 5.0 | 6.0 |
| Denmark | 2.0 | 2.0 | 2.0 | 2.0 | 2.0 | | |
| Finland | 3.0 | 3.0 | 3.0 | 3.0 | 3.0 | | |
| France | 1.0 | 1.0 | 1.0 | 1.0 | 1.0 | 1.0 | 1.0 |
| Germany | 3.9 | 3.1 | 3.0 | 6.1 | | 4.0 | |
| Greece | 1.0 | 2.0 | 1.0 | 4.0 | | 5.0 | |
| Ireland | 3.0 | 3.0 | 3.0 | 13.0 | | 13.0 | 13.0 |
| Italy | 4.0 | 4.0 | 4.9 | 4.0 | 4.0 | 4.9 | 4.0 |
| Luxembourg | 5.0 | 1.0 | 1.0 | 1.0 | | 1.0 | 1.0 |
| Netherlands | 4.0 | 4.0 | 4.0 | 7.0 | | | |
| Norway | 1.0 | 1.0 | 1.0 | | 3.0 | | |
| Portugal | 1.0 | 1.0 | 6.0 | 6.0 | | 6.0 | |
| Spain | 1.7 | 1.2 | 1.4 | 1.3 | 1.4 | 1.3 | 13.0 |
| Sweden | 16.0 | | | 16.0 | | | |
| Switzerland | 16.9 | 1.5 | 1.5 | 3.3 | 5.2 | | 2.0 |
| United Kingdom | 4.0 | 3.0 | 3.0 | 3.0 | | 3.0 | |
| United States | 1.8 | 2.0 | 3.0 | 2.0 | 2.3 | 8.2 | 7.1 |
| Average | 4.3 | 2.5 | 2.6 | 4.6 | 2.7 | 4.3 | 5.9 |

**Table 3.** Distance (in days) to the next NPIs in time per contry and NPI. "Average" is the mean distance over all countries. Computation: For each NPI, the distance is measured as the absolute difference (in days) to the NPI that was implemented closest before or after, which is then averaged across countries. For countries where NPIs were implemented at the regional level, the absolute differences were first averaged over all regions. If an NPI was not implemented, then the absolute difference is omitted from computation.

| | Event ban | School closure | Venue closure | Gathering ban | Border closure | Stay-at-home order | Work ban |
|---|---|---|---|---|---|---|---|
| Event ban | 0.0 | 4.1 | 4.3 | 7.4 | 7.6 | 9.1 | 12.4 |
| School closure | 4.1 | 0.0 | 2.5 | 4.4 | 5.4 | 6.4 | 8.9 |
| Venue closure | 4.3 | 2.5 | 0.0 | 3.4 | 4.8 | 4.9 | 7.1 |
| Gathering ban | 7.4 | 4.4 | 3.4 | 0.0 | 5.2 | 2.7 | 6.9 |
| Border closure | 7.6 | 5.4 | 4.8 | 5.2 | 0.0 | 6.8 | 7.8 |
| Stay-at-home order | 9.1 | 6.4 | 4.9 | 2.7 | 6.8 | 0.0 | 4.7 |
| Work ban | 12.4 | 8.9 | 7.1 | 6.9 | 7.8 | 4.7 | 0.0 |

**Table 4.** Pairwise average distance (in days) between the implementation of NPIs across countries. Computation: For each pair of NPIs, the distance is measured as the absolute difference (in days) between the implementation dates of NPIs, which is then averaged across countries. For countries where NPIs were implemented at the regional level, the absolute differences were first averaged over all regions. If an NPI was not implemented, then the absolute difference is omitted from computation.

### 3 Estimation results

#### 3.1 Estimated model parameters

Tbl. 5 presents posterior means and credible intervals for all model parameters. See the main paper for a discussion of the effects of NPIs. Here we briefly discuss some of the additional model parameters.

The ratio of the effective sample size ($\hat{n}_{\text{eff}}/N$) and the Gelman-Rubin convergence diagnostic ($\hat{R}$) indicates good estimation power. It further suggests that the Markov chains converged.

There is a country-specific variation in the intercept parameter ($\alpha_j$), reflecting differences in the rate of new cases – i.e., spread of the disease – in the absence of any NPI. Australia, Norway and Sweden are among the countries with lowest estimated rate and Canada, Ireland, and the US are among the countries with highest estimated rate.

The expected number of new infections at start of the non-modeling phase is around 4, but the exact number varies between countries, with around 11 infected in Australia and 1 infected in the US.

The overdispersion parameter ($\phi$) can be rather precisely estimated to be in the magnitude of 4 for the number of new cases and 8 for the number of new infections, i.e., we are far away from the case of no overdispersion ($\phi = \infty$). This coincides with the empirical observation of rather unsmooth trajectories in each country (Section 5).

The posterior distribution of the parameters of the log normal distribution describing the time from infection to reporting suggest that we can estimate them with a rather high precision (Fig. 4a-b). The posterior mean of $\mu^{p_{\text{IN}}} = 2.62$ and $\sigma^{p_{\text{IN}}} = 0.18$ corresponds to a mean delay of about 15 days, which is slightly higher when compared to the prior. The posterior distribution of sigma is placed distinctly below the prior mean value of 0.4. Both together implies a rather precise posterior knowledge about the distribution of the time from infection to reporting with a range from roughly 8 to 25 days.

| | Mean | Lower CrI | Upper CrI | $\hat{n}_{\text{eff}}/N$ | $\hat{R}$ |
|---|---|---|---|---|---|
| $\alpha$ | 0.83 | 0.73 | 0.94 | 0.24 | 1.00 |
| $\tau$ | 0.20 | 0.13 | 0.29 | 0.67 | 1.00 |
| $\alpha_1$ | −0.32 | −0.48 | −0.17 | 0.35 | 1.00 |
| $\alpha_2$ | −0.11 | −0.27 | 0.04 | 0.36 | 1.00 |
| $\alpha_3$ | 0.18 | 0.04 | 0.33 | 0.32 | 1.00 |
| $\alpha_4$ | 0.24 | 0.10 | 0.38 | 0.27 | 1.00 |
| $\alpha_5$ | −0.12 | −0.28 | 0.04 | 0.33 | 1.00 |
| $\alpha_6$ | −0.05 | −0.21 | 0.12 | 0.26 | 1.00 |
| $\alpha_7$ | 0.16 | 0.02 | 0.32 | 0.30 | 1.00 |
| $\alpha_8$ | 0.02 | −0.12 | 0.15 | 0.35 | 1.00 |
| $\alpha_9$ | −0.13 | −0.29 | 0.02 | 0.34 | 1.00 |
| $\alpha_{10}$ | 0.20 | 0.05 | 0.36 | 0.38 | 1.00 |
| $\alpha_{11}$ | 0.06 | −0.06 | 0.19 | 0.32 | 1.00 |
| $\alpha_{12}$ | −0.18 | −0.35 | −0.01 | 0.32 | 1.00 |
| $\alpha_{13}$ | 0.01 | −0.13 | 0.16 | 0.33 | 1.00 |
| $\alpha_{14}$ | −0.25 | −0.41 | −0.10 | 0.35 | 1.00 |
| $\alpha_{15}$ | 0.01 | −0.14 | 0.16 | 0.33 | 1.00 |
| $\alpha_{16}$ | 0.12 | −0.02 | 0.26 | 0.31 | 1.00 |
| $\alpha_{17}$ | −0.18 | −0.35 | 0.00 | 0.19 | 1.01 |
| $\alpha_{18}$ | 0.00 | −0.15 | 0.16 | 0.21 | 1.01 |
| $\alpha_{19}$ | 0.04 | −0.10 | 0.17 | 0.37 | 1.00 |
| $\alpha_{20}$ | 0.33 | 0.20 | 0.47 | 0.30 | 1.00 |
| $\theta_1$ | 0.17 | −0.02 | 0.36 | 0.45 | 1.00 |
| $\theta_2$ | 0.10 | −0.02 | 0.21 | 0.30 | 1.00 |
| $\theta_3$ | 0.37 | 0.21 | 0.50 | 0.09 | 1.02 |
| $\theta_4$ | 0.09 | −0.04 | 0.23 | 0.36 | 1.00 |
| $\theta_5$ | 0.18 | −0.04 | 0.40 | 0.16 | 1.01 |
| $\theta_6$ | 0.04 | −0.06 | 0.17 | 0.43 | 1.00 |
| $\theta_7$ | 0.01 | −0.08 | 0.12 | 0.64 | 1.00 |
| $\lambda$ | 4.41 | 2.39 | 7.32 | 0.14 | 1.00 |
| $I_{1-33}$ | 11.02 | 3.08 | 24.93 | 0.18 | 1.01 |
| $I_{2-33}$ | 7.20 | 1.84 | 17.59 | 0.22 | 1.00 |
| $I_{3-33}$ | 1.22 | 0.14 | 3.59 | 0.34 | 1.00 |
| $I_{4-33}$ | 1.04 | 0.10 | 3.28 | 0.27 | 1.00 |
| $I_{5-33}$ | 7.94 | 2.07 | 18.97 | 0.19 | 1.01 |
| $I_{6-33}$ | 2.83 | 0.42 | 8.02 | 0.21 | 1.00 |
| $I_{7-33}$ | 1.90 | 0.30 | 5.45 | 0.25 | 1.00 |
| $I_{8-33}$ | 5.07 | 1.30 | 12.67 | 0.20 | 1.01 |
| $I_{9-33}$ | 3.99 | 0.80 | 10.17 | 0.23 | 1.00 |
| $I_{10-33}$ | 1.28 | 0.15 | 3.99 | 0.41 | 1.00 |
| $I_{11-33}$ | 5.29 | 1.38 | 12.73 | 0.18 | 1.01 |
| $I_{12-33}$ | 10.50 | 2.73 | 25.32 | 0.21 | 1.00 |
| $I_{13-33}$ | 3.62 | 0.76 | 9.27 | 0.29 | 1.00 |
| $I_{14-33}$ | 10.46 | 3.01 | 23.95 | 0.17 | 1.00 |
| $I_{15-33}$ | 4.78 | 1.06 | 12.39 | 0.22 | 1.01 |
| $I_{16-33}$ | 3.78 | 0.78 | 9.49 | 0.23 | 1.00 |
| $I_{17-33}$ | 7.19 | 1.85 | 17.26 | 0.36 | 1.00 |
| $I_{18-33}$ | 6.31 | 1.61 | 15.45 | 0.26 | 1.00 |
| $I_{19-33}$ | 3.36 | 0.71 | 8.49 | 0.25 | 1.00 |
| $I_{20-33}$ | 1.28 | 0.16 | 3.82 | 0.18 | 1.01 |
| $\phi^N$ | 4.24 | 3.80 | 4.71 | 0.72 | 1.00 |
| $\phi^I$ | 8.66 | 6.70 | 11.44 | 0.05 | 1.02 |
| $\mu^{p_{IN}}$ | 2.62 | 2.51 | 2.73 | 0.05 | 1.03 |
| $\sigma^{p_{IN}}$ | 0.18 | 0.11 | 0.27 | 0.29 | 1.00 |

**Table 5.** Estimation results for the main analysis. The ratio $\hat{n}_{\text{eff}}$ / $N$ is the effective sample size ($\hat{n}_{\text{eff}}$) divided by the total sample size ($N$). Generally, ratios above 0.5 correspond to high, between 0.1 and 0.5 to medium and below 0.1 to low estimation power[17]. $\hat{R}$ is the Gelman-Rubin convergence diagnostic[18]. A $\hat{R} \approx 1.00$ indicates good convergence, while a $\hat{R} > 1.10$ indicates bad convergence[19].

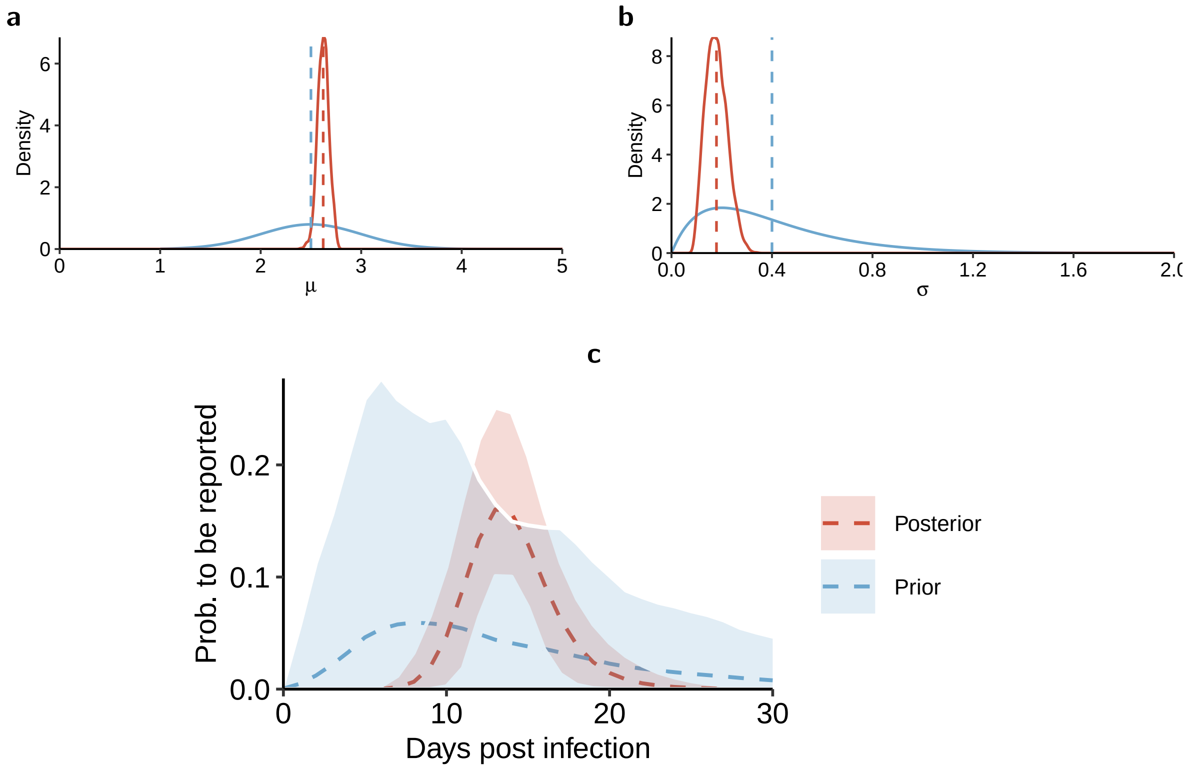

**Figure 4.** Distribution of the time from infection to reporting of a new case. **(a)** Log mean $\mu$ (prior and posterior mean as dashed lines) **(b)** Log standard deviation $\sigma$ (prior and posterior mean as dashed lines). **(c)** Posterior distribution of $p_{\mathrm{IN}}(t)$ for $t = 0, 1, \ldots, 30$ (prior and posterior mean as dashed lines with 95 % range and 95 % credible interval as shaded area, based on 4,000 draws from the prior and posterior distributions for $\mu$ and $\sigma$, respectively).

##### 3.2 Checking for correlations between parameters

A similar timing could make it difficult to distinguish the individual effects of NPIs. To investigate this issue, Fig. 5 depicts the pairwise bivariate posterior distributions of the parameters of the NPIs. We observe a tendency towards negative correlations, reflecting the difficulty to distinguish the effects of NPIs, which were often introduced close in time. That is, highest negative correlations were observed for the pair of NPIs with the smallest average distance in implementation (Fig. 4).

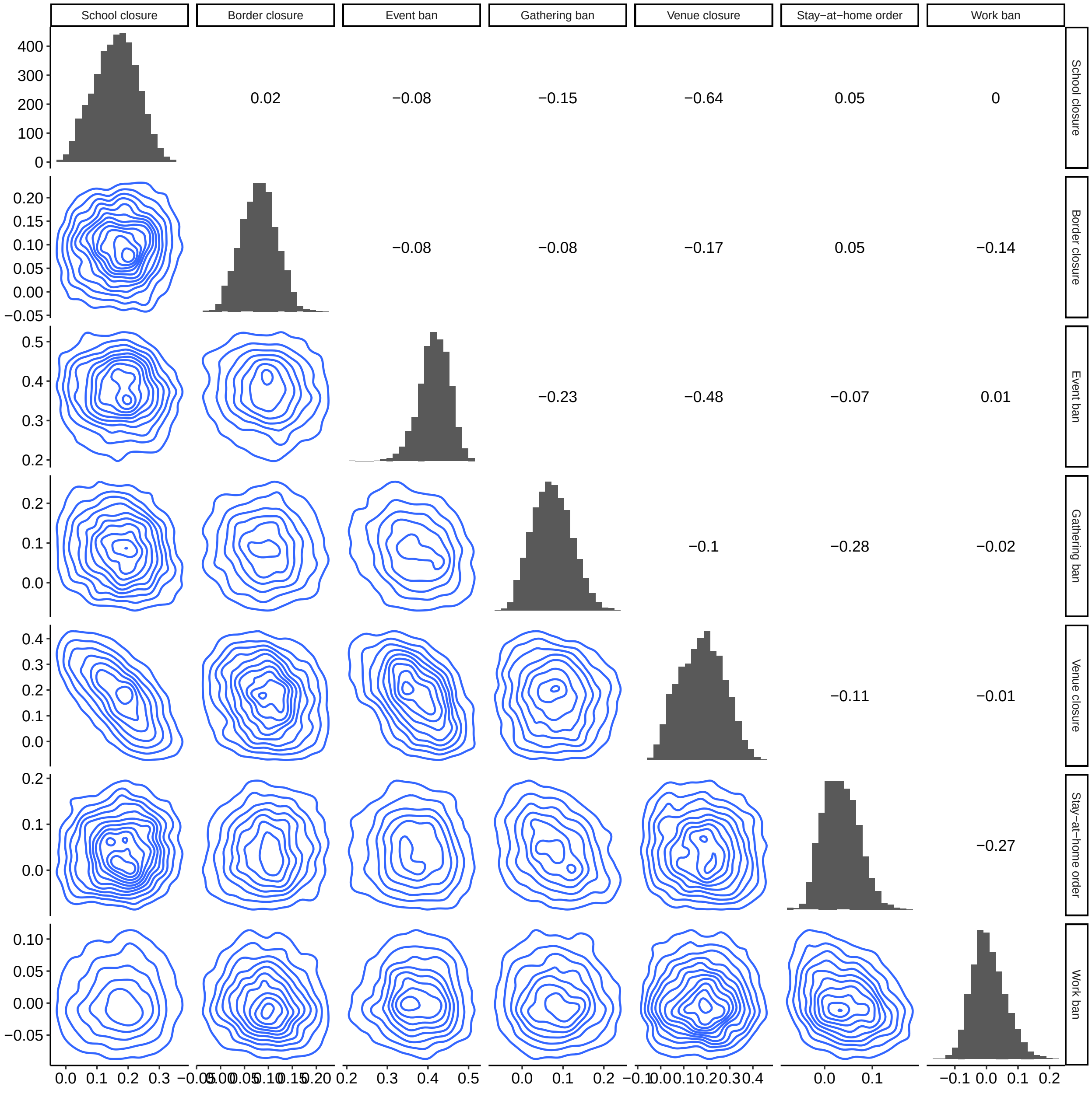

**Figure 5.** Bivariate posterior distributions visualized by contour plots (lower diagonal matrix) from the MCMC sample for the parameters of non-pharmaceutical interventions (NPIs). Pearson's $r$ is shown in the upper diagonal matrix, and the marginal distribution is depicted as a histogram on the diagonal of the matrix.

##### 3.3 Checking for influential observations

Influential observations can affect parameter estimates. To check for influential observations, Fig. 6 shows the tail shape parameter $k$ from approximate leave-one-out (LOO) cross-validation using Pareto smoothed importance sampling[20]. Only few observations seem to be highly influential.

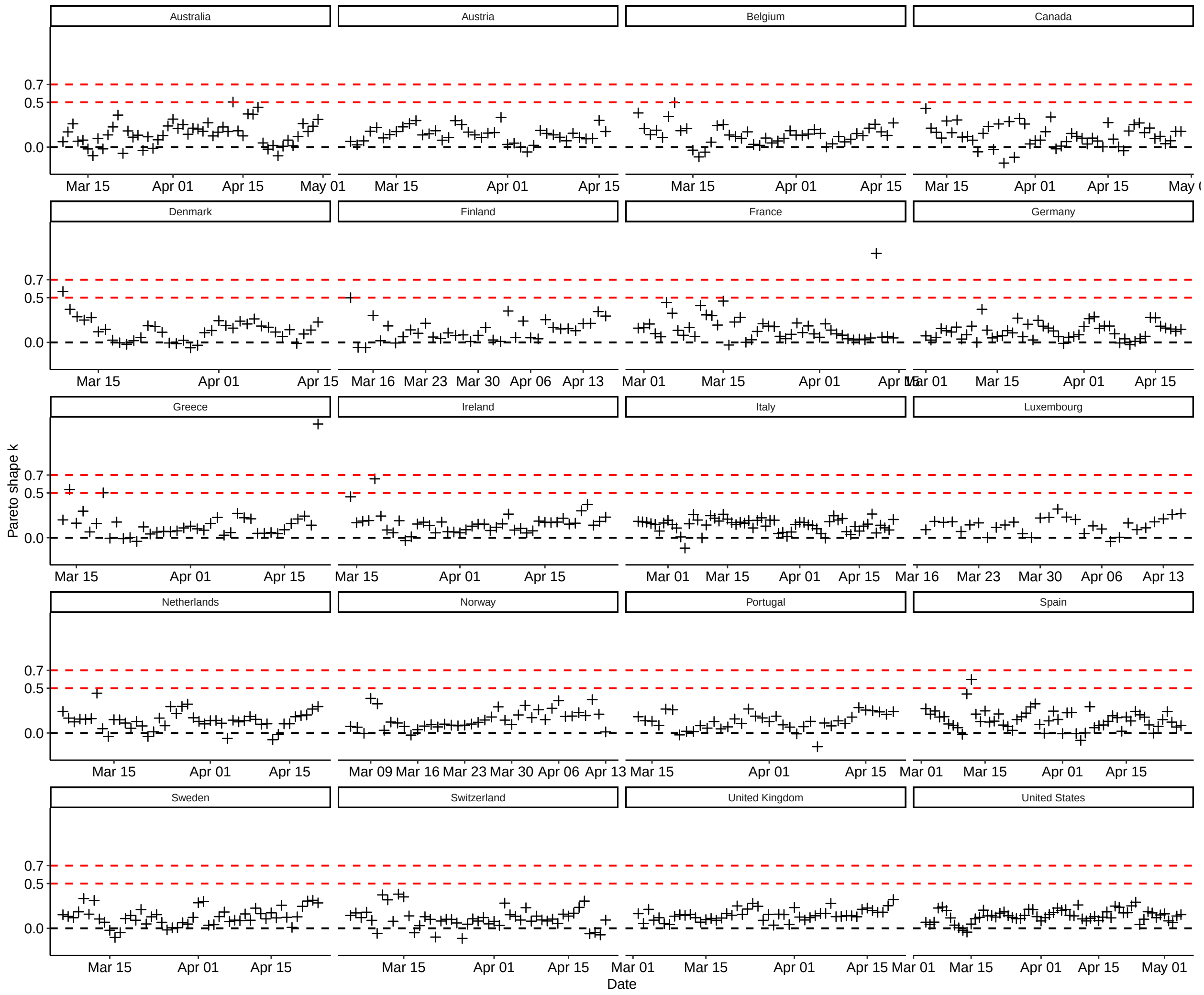

**Figure 6.** Model diagnostics based on influential observations. Shown is the tail shape parameter $k$ of the generalized Pareto distribution from approximate leave-one-out (LOO) cross-validation using Pareto smoothed importance sampling for each observation by country and time. Values below 0.5 indicate that the observation is not influential; values between 0.5 and 0.7 indicate that the observation might be influential but the model is usually still robust; values above 0.7 indicate the observation is influential and that the model may not be robust[20].

#### 4 Sensitivity analysis

##### 4.1 Modeling phase starting from 10 cumulative cases onward

In the main model, the modeling phase starts after 100 cumulative cases were reported by a country. This is because countries needed time to set up documentation practices and thus the early reported cases numbers were very variable. Fig. 7 shows the estimated NPI effects when modeling starts after 10 cumulative cases were reported. Overall, NPI effects are not sensitive to a start of the modeling phase after 50 or 100 cumulative cases were observed. The effects of event ban, venue closure, and gathering ban are sensitive to a very early start of the modeling phase after 10 cumulative cases, but it should be noted that case numbers were highly variable in the very beginning.

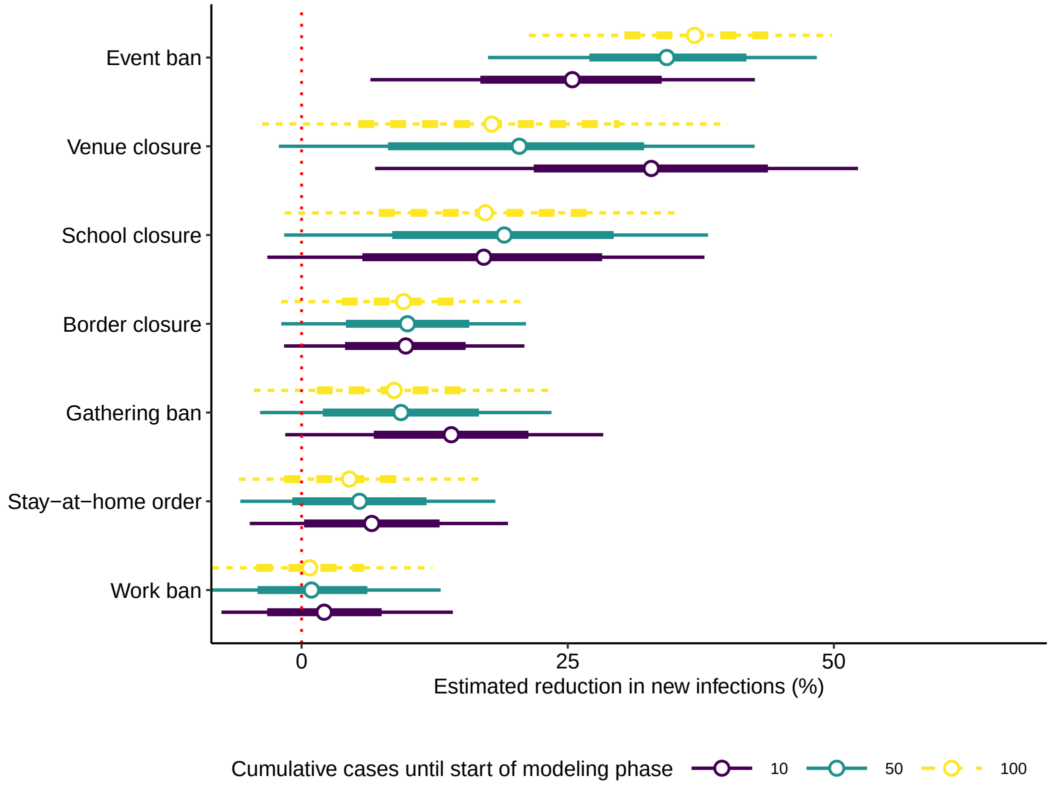

**Figure 7.** Reduction (posterior mean as dots with 80% and 95% credible interval as thick and thin lines, respectively) in the number of new infections (in %) for each non-pharmaceutical intervention (NPI) when varying the start of the modeling phase (default in main model as dashed yellow line).

#### 4.2 Modeling phase ending 21 to 35 days after last NPI was implemented

In the main model, the modeling phase ends 28 days after the last NPI was implemented. This should provide enough time for the effect of NPIs to show up in the number of reported cases, thereby concluding the first wave of the epidemic. Fig. 8 shows the estimated NPI effects when the end of the modeling phase is varied from 21 to 25 days after the last NPI was implemented within a country. NPI effects are a little sensitive to an earlier end of the modeling phase. The wider credible intervals for border closure, gathering ban, stay-at-home order and work ban could indicate that ending the modeling phase 21 days after the last NPI was implemented is a bit too early.

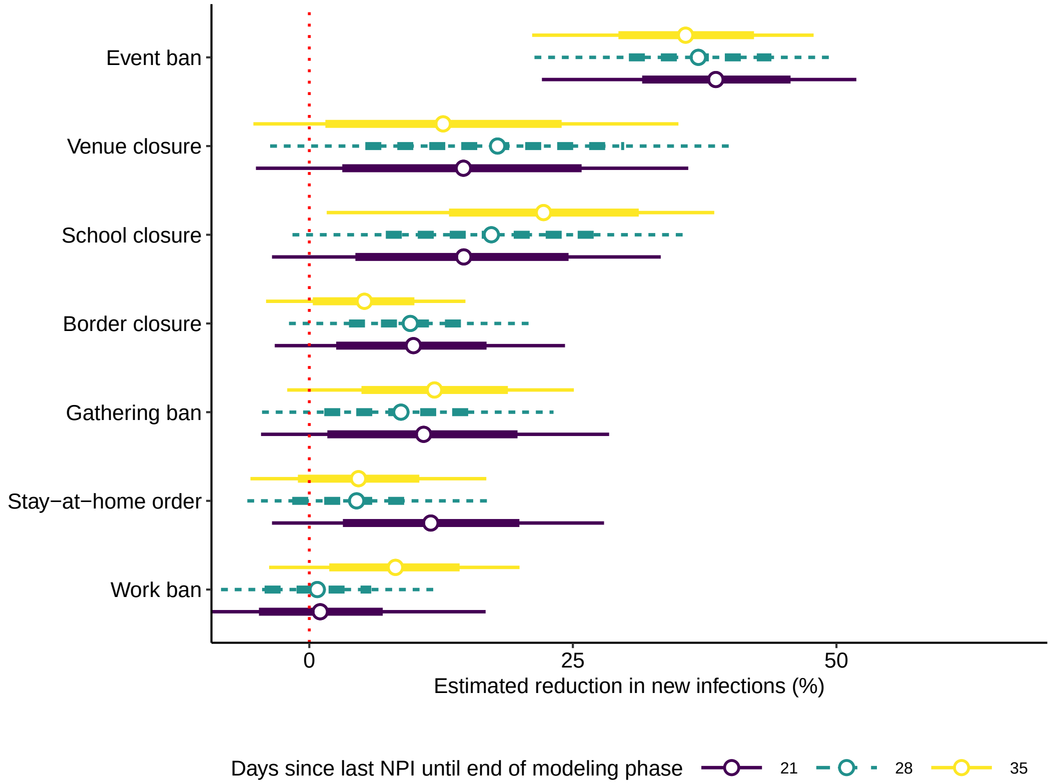

**Figure 8.** Reduction (posterior mean as dots with 80% and 95% credible interval as thick and thin lines, respectively) in the number of new infections (in %) for each non-pharmaceutical intervention (NPI) when varying the end of the modeling phase (default in main model as dashed turquoise line).

##### 4.3 Varying the time-delayed response functions

In the main model, a time delayed response function was considered where the effect of an NPI increases linearly between $t_0 = 0$ days and $t_1 = 3$ days after their implementation. Fig. 9 shows the estimated NPI effects when varying $t_0$ and $t_1$ considering the following alternatives for the first-order spline FOS($t_0$,$t_1$): a proactive response FOS($-1$,0), a quicker response FOS(0,1), and a more delayed response FOS(0,5) as compared to the main model. Overall, the results are not sensitive to the choice of $t_0$ and $t_1$ in the FOS.

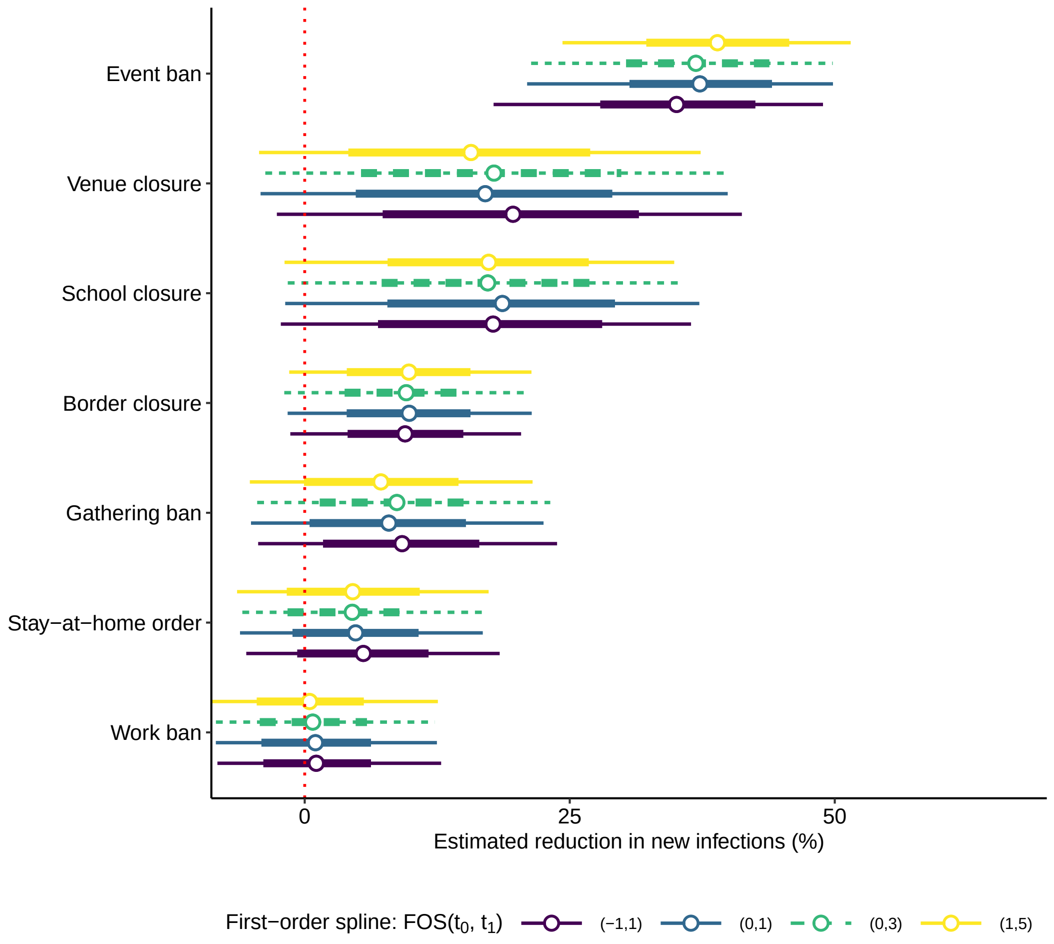

**Figure 9.** Reduction (posterior mean as dots with 80% and 95% credible interval as thick and thin lines, respectively) in the number of new infections (in %) for each non-pharmaceutical intervention (NPI) when varying time delayed response function (default in main model as dashed turquoise line).

##### 4.4 Varying the prior distribution for the effects of non-pharmaceutical interventions

In the main model, a mixture prior for the NPI effects was constructed where the probability of a negative effect (i.e., NPIs leading to an increase in the number of new cases) is 10 %. Fig. 10 shows the estimated NPI effects when the prior probability of a negative effect of NPIs is alternatively 30 % or 50 %. The ranking of the posterior mean effects does not depend on the prior, but the range of effects for venue closure and work ban would include larger negative effects when increasing the prior probability for a negative effect.

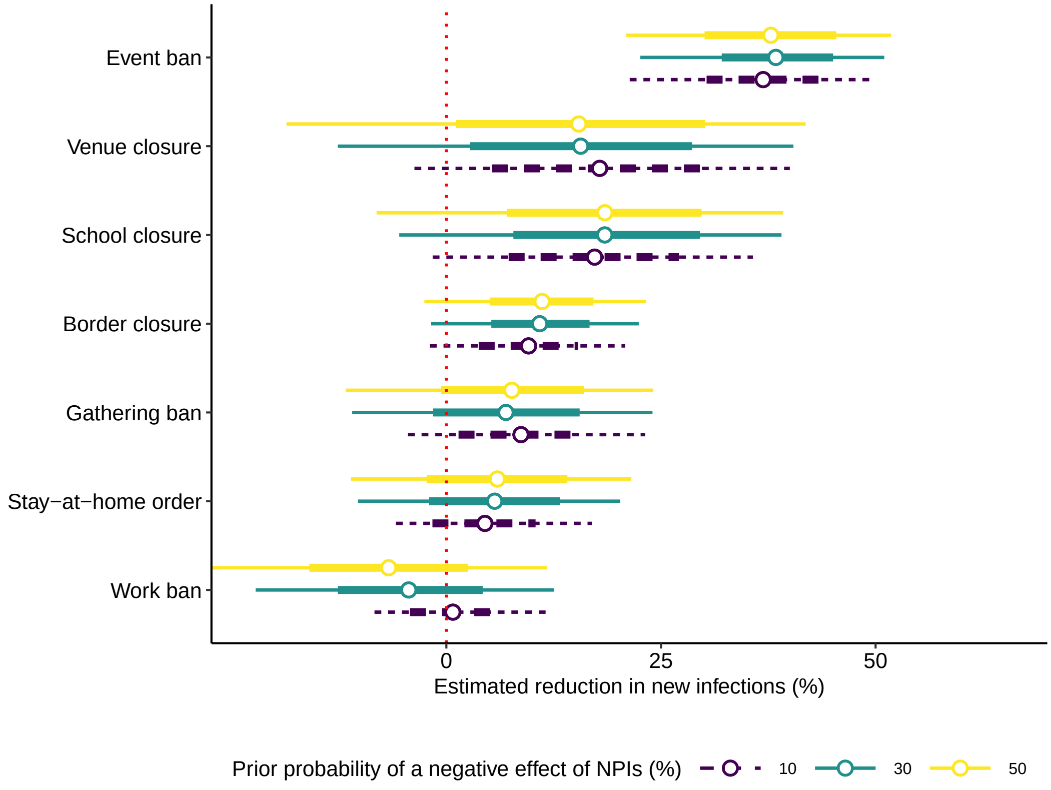

**Figure 10.** Reduction (posterior mean as dots with 80% and 95% credible interval as thick and thin lines, respectively) in the number of new infections (in %) for each non-pharmaceutical intervention (NPI) when varying the probability of a negative effect (in %) in the prior distribution for the effects of non-pharmaceutical interventions (default in main model as dashed purple line).

##### 4.5 Varying the prior distribution for the time from infection to reporting of a new case

In the main model, the probability distribution for the time from infection to reporting of a new case $p_{\mathrm{IN}}(t)$ was inferred by estimating the log mean $\mu$ and the log standard deviation $\sigma$ of the assumed Log-normal distribution. The specified priors in the main model correspond to a $\mathrm{Normal}(\mu_0 = 2.47, \sigma_0 = 0.45)$ for the log mean and $\mathrm{Gamma}(\alpha_0 = 2, = \beta_0 = 4)$ for the log standard deviation, such that $\mu = \mu_0 = 2.47$ and $\sigma = \frac{\alpha_0}{\beta_0} = \frac{2.00}{4.48} \approx 0.45$ correspond to the prior means of a $\mathrm{Log-normal}(\mu = 2.47, \sigma = 0.45)$ that was obtained based on prior knowledge. For this sensitivity check, $\mu_0$ and $\beta_0$ were varied in the Normal prior for the log mean $\mu$ and the Gamma prior for the log standard deviation $\sigma$, resulting in alternative probability distributions for $p_{\mathrm{IN}}(t)$ (Fig. 11). Fig. 12 shows the estimated NPI effects for this sensitivity check. Overall, NPI effects are not sensitive to the choice of priors for the parameters of $p_{\mathrm{IN}}(t)$.

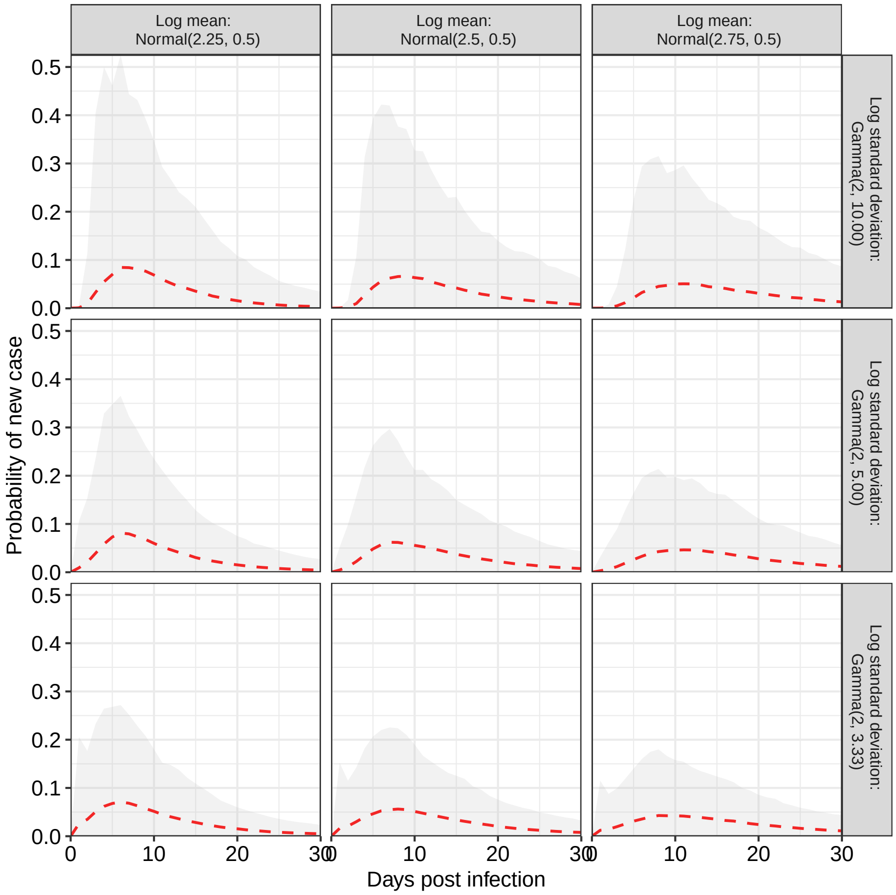

**Figure 11.** Prior choices for the distribution of the time from infection to reporting of a new case $p_{\mathrm{IN}}(t)$ depending on log mean $\mu$ and log standard deviation $\sigma$ (prior mean as dashed red line with 95 % range as shaded area, based on 4,000 independent draws from the distributions for the log mean $\mu$ (column) in the sensitivity analysis (default in main model as middle tile).

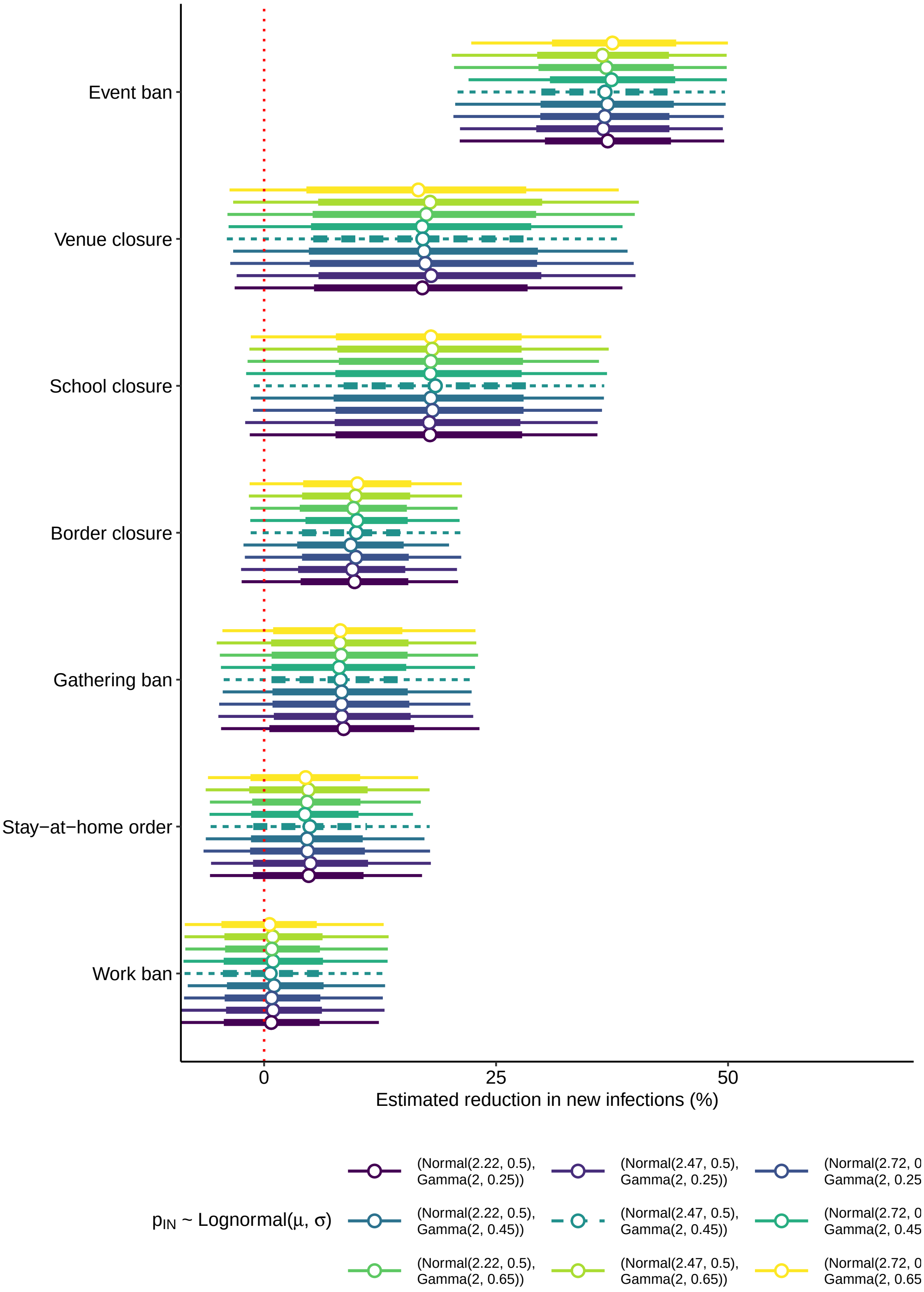

**Figure 12.** Reduction (posterior mean as dots with 80% and 95% credible interval as thick and thin lines, respectively) in the number of new infections (in %) for each non-pharmaceutical intervention (NPI) when varying the prior choices for distribution of the time from infection to reporting of a new case (default in main model as dashed turquoise line).

#### 4.6 Varying the generation time distribution

In the main model, the generation time distribution $p_G(t)$ was assumed to be a Weibull($\alpha = 3.28, \beta = 6.12$)[13]. For this sensitivity check, the shape parameter $\alpha$ and inverse scale parameter $\beta$ were varied, resulting in the alternative probability distributions for $p_G(t)$ (Fig. 13). Fig. 14 shows the estimated NPI effects for this sensitivity check. Overall, NPI effects are not very sensitive to the choice of $p_G(t)$, except that the estimated effects tend to increase for longer generation times.

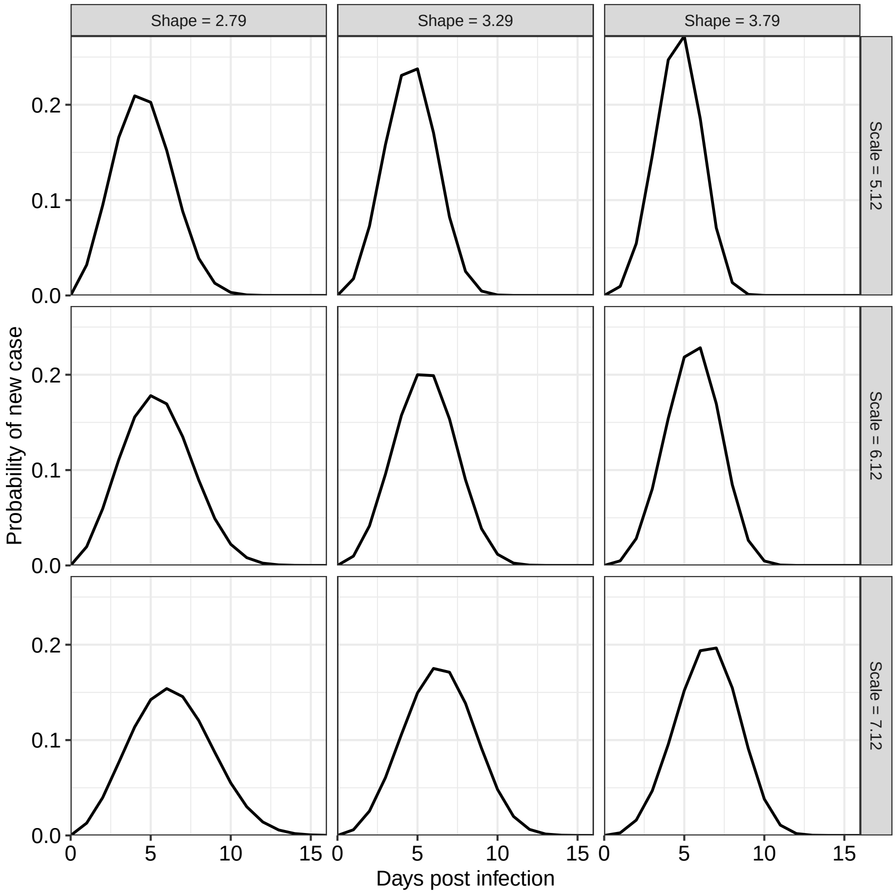

**Figure 13.** Prior choices for the generation time distribution $p_G(t)$ depending on shape parameter $\alpha$ and rate parameter $\kappa$ in the sensitivity analysis (default in main model as middle tile).

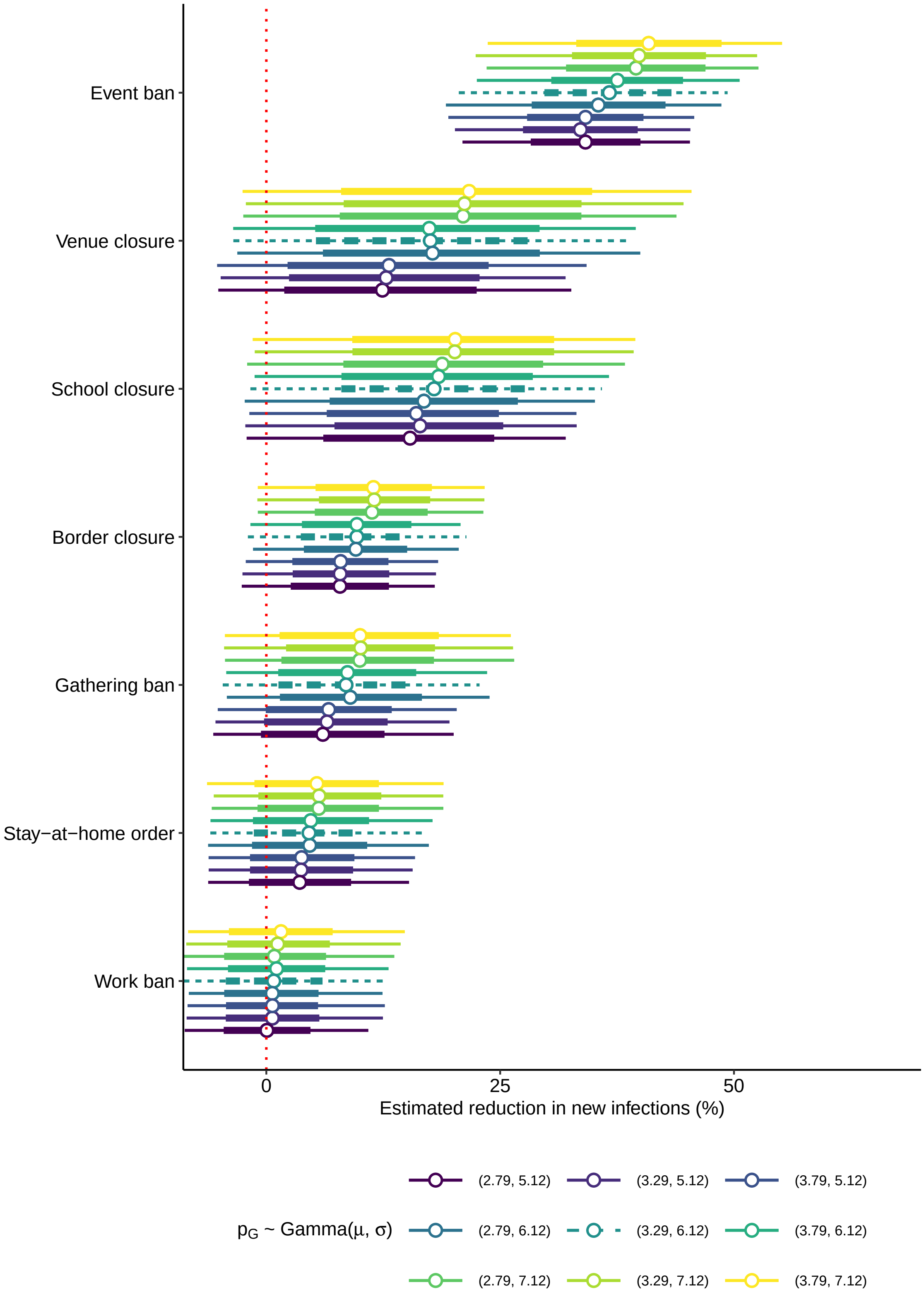

**Figure 14.** Reduction (posterior mean as dots with 80% and 95% credible interval as thick and thin lines, respectively) in the number of new infections (in %) for each non-pharmaceutical intervention (NPI) when varying the generation time distribution (default in main model turquoise as dashed line).

##### 4.7 Analyzing the influence of leaving out one country at the time

A leave-one-out analysis was conducted to analyze the influence of individual countries for the estimated NPI effects, i.e., the model is re-estimated leaving out one country at a time. Fig. 15 shows the results of this analysis for each NPI. Some estimated effects seem sensitive to the exclusion of individual countries.

The effect of school closure would be higher when estimating the model without Australia. This could implicate sensitivity or, instead, it could be that the particular country is informative for the estimated effect of a particular NPI. Australia is a good example to check for this, as here, school closures where implemented in Queensland, New South Wales, Victoria and Australian Capital Territory (Eastern Australia) but not in Western Australia, Northern Territory and South Australia (Western Australia). As a result, Australia was split into Eastern and Western Australia and the model was estimated leaving out one region at a time. This time, the effect of school closure is lower without Eastern Australia and higher without Western Australia (Fig. 16). A reason for this could be that both regions successfully reduced the number of cases, but Western Australia did so with similar measures as Eastern Australia except for closing schools, thereby providing substantial evidence against the particular effectiveness of school closures.

The effect of event bans would be higher without Switzerland and Sweden. Note that both countries implemented event bans comparably early into the epidemic. Despite that, the number of new infections was still increasing in Switzerland for a couple of weeks and continuously in Sweden, thereby indicating that event bans were potentially not as effective as in other countries.

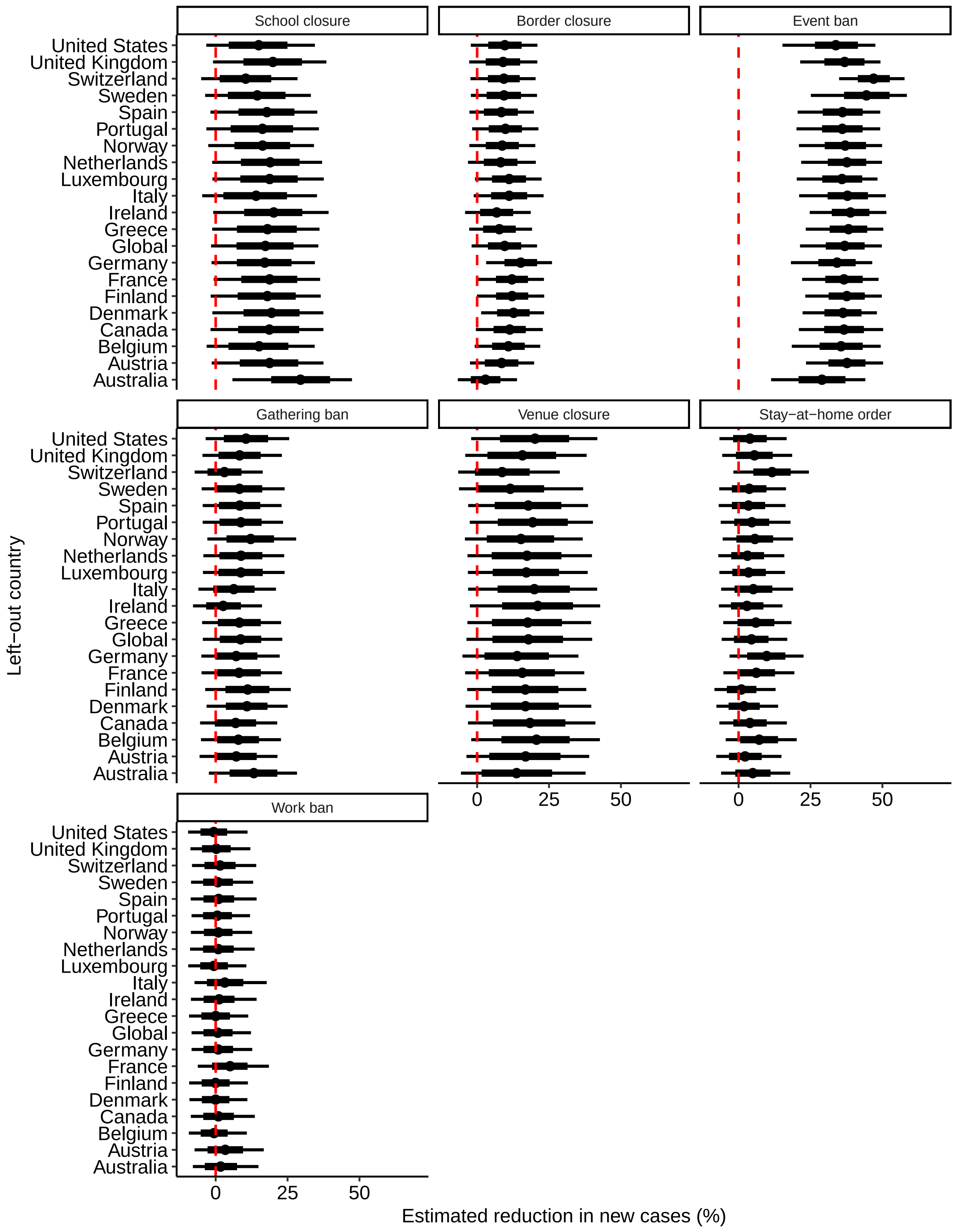

**Figure 15.** Reduction (posterior mean as dots with 80% and 95% credible interval as thick and thin lines, respectively) in the number of new infections (in %) for each non-pharmaceutical intervention (NPI) when leaving out one country at a time.

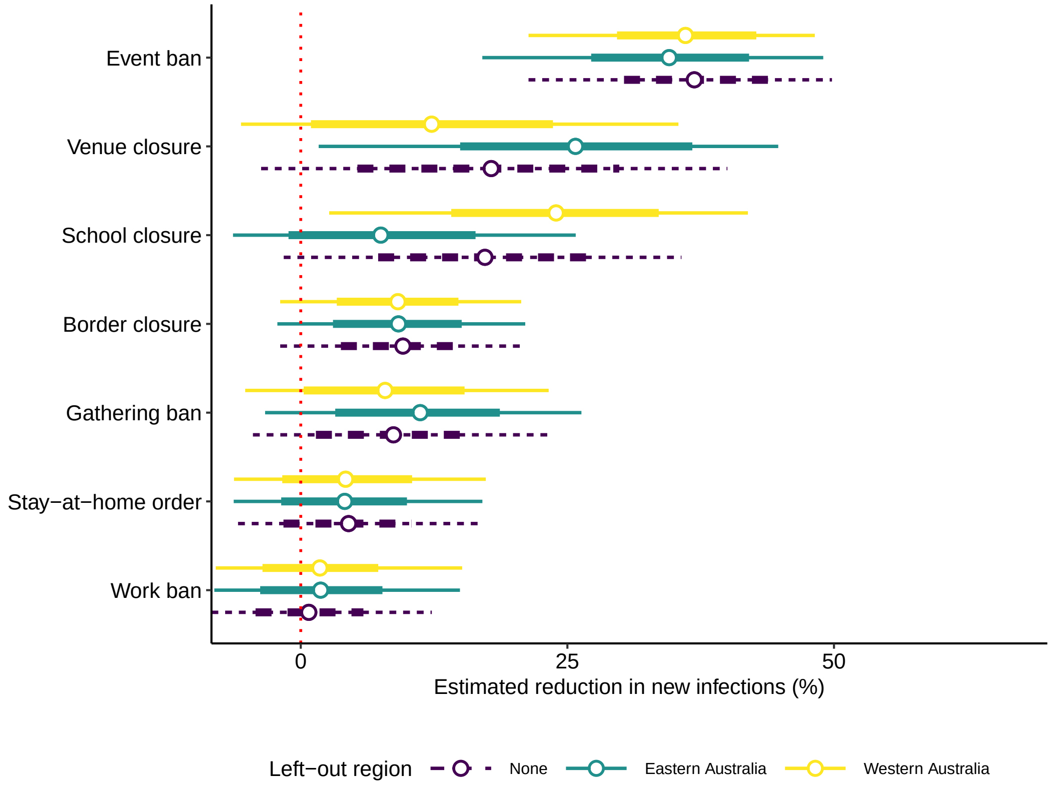

**Figure 16.** Reduction (posterior mean as dots with 80% and 95% credible interval as thick and thin lines, respectively) in the number of new infections (in %) for each non-pharmaceutical intervention (NPI) when dividing Australia into two sub-regions and leaving-out Eastern or Western Australia (default in main model as dashed line).

#### 5 Visual Inspection of the Model Fit

Fig. 17 shows the expected number of new infections ($\mu^I$) and new cases ($\mu^N$) over time for each country. The estimated numbers are compared to the observed number of new cases in order to assess the model fit. Overall, our model provides a reasonable fit in the sense that the expected number of new cases follow the development of the observed number of new cases in each country. Furthermore, changes in the expected number of new infections clearly follow the implementation of NPIs. The size of the credible intervals reflect varying uncertainty in the expected number of new infections and cases in each country (e.g., compare Germany (small) to France (large)). For a couple of countries (e.g., Ireland, Sweden, United Kingdom, United States), the credible intervals are particularly large at the very end of the epidemic. This corresponds to a rather high level of new infections at that time, indicating that the interventions were not yet enough to strongly reduce the number of new infections. The large credible intervals are a reminder that this implies a risk for a new exponential increase in the number of new infections.

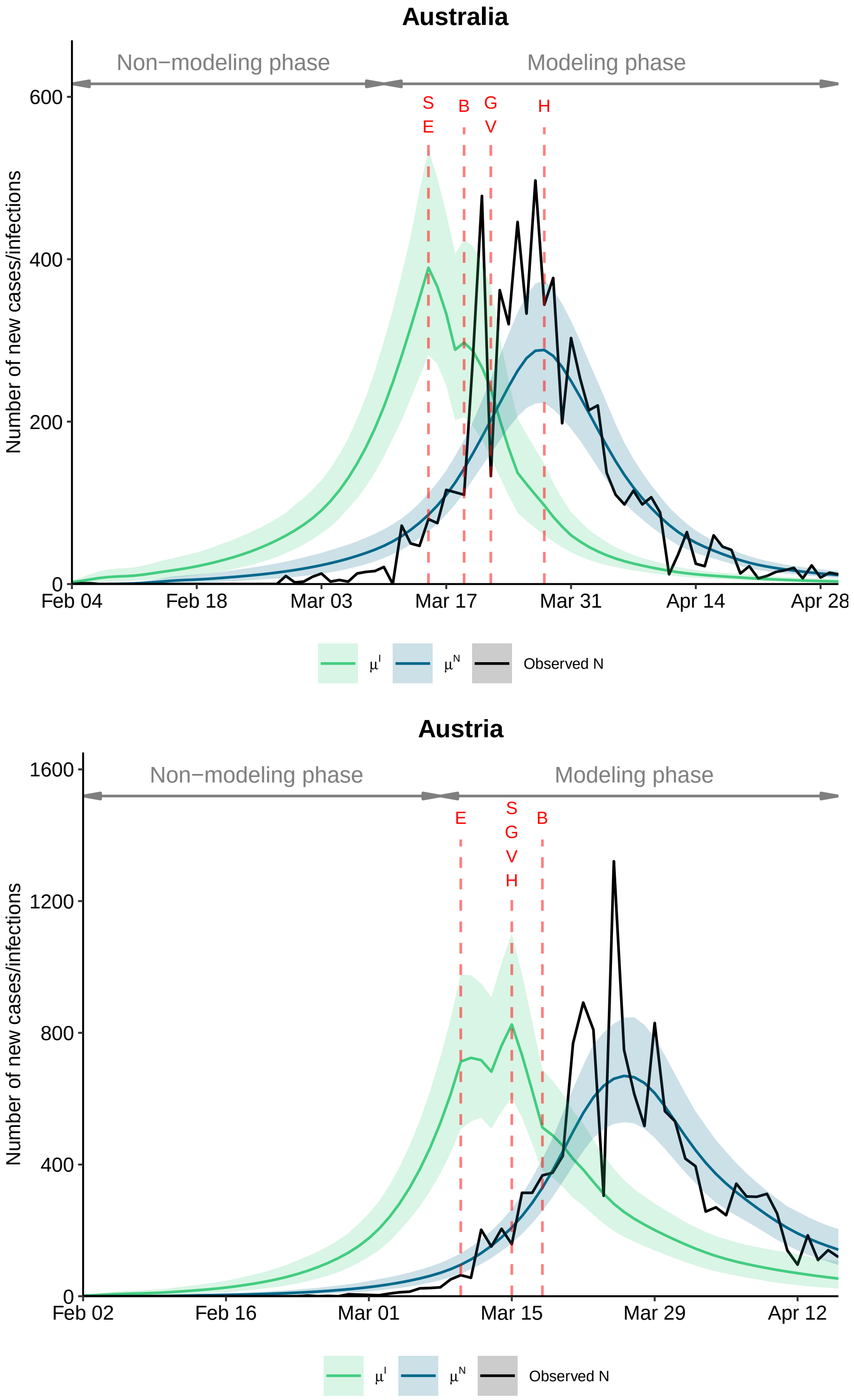

**Figure 17.** Expected number of new infections $\mu^I$ and new cases $\mu^N$ (posterior mean as colored lines with 95% credible interval as shaded area) and the observed number of new cases by country over time. Red letters and lines indicate the first day an NPI was implemented within a country (S: School closures, B: Border closure, E: Event ban, G: Gathering ban, V: Venue closure, H: Stay-at-home order, W: Work ban).

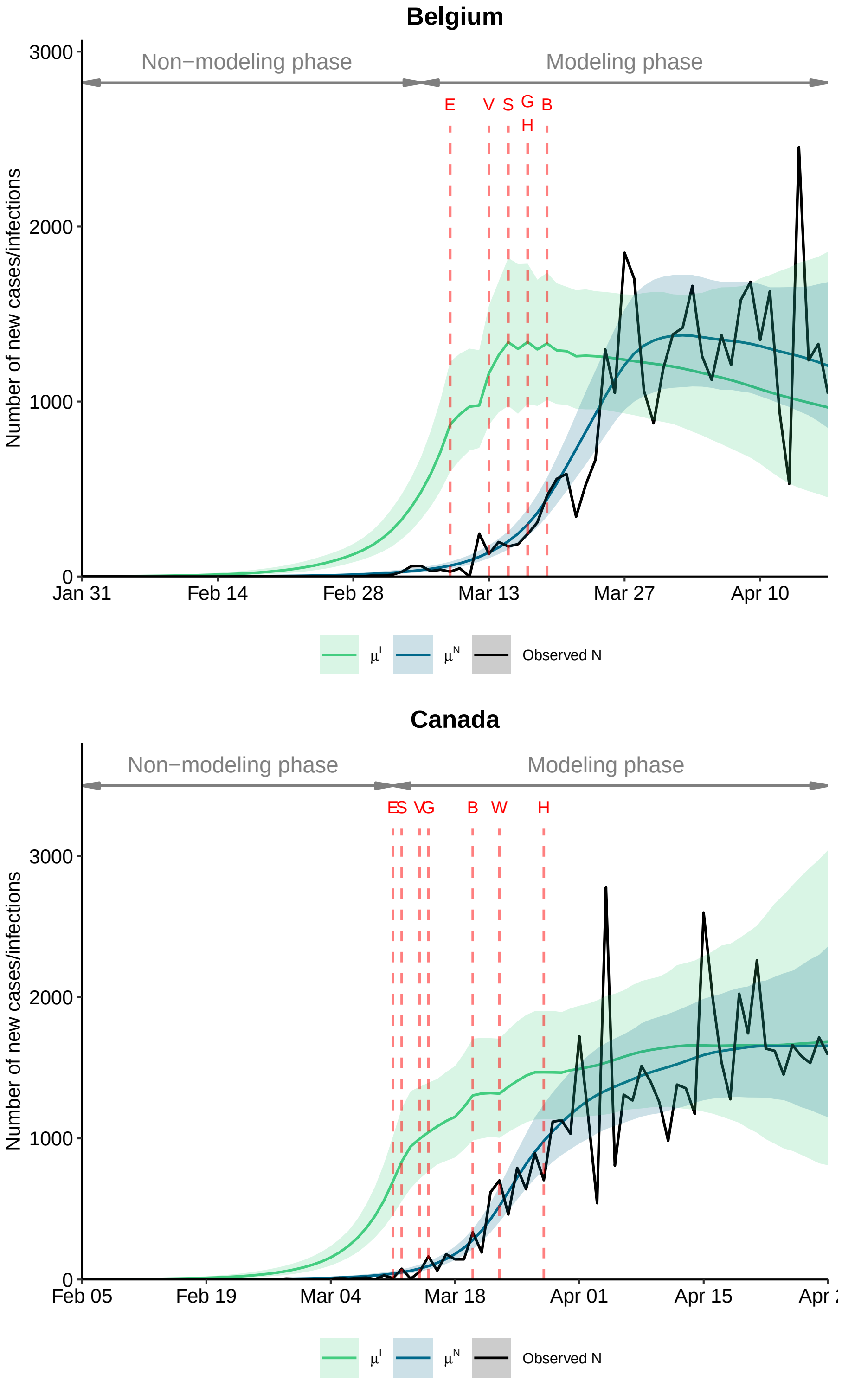

**Figure 17.** Expected number of new infections $\mu^I$ and new cases $\mu^N$ (posterior mean as colored lines with 95% credible interval as shaded area) and the observed number of new cases by country over time. Red letters and lines indicate the first day an NPI was implemented within a country (S: School closures, B: Border closure, E: Event ban, G: Gathering ban, V: Venue closure, H: Stay-at-home order, W: Work ban).

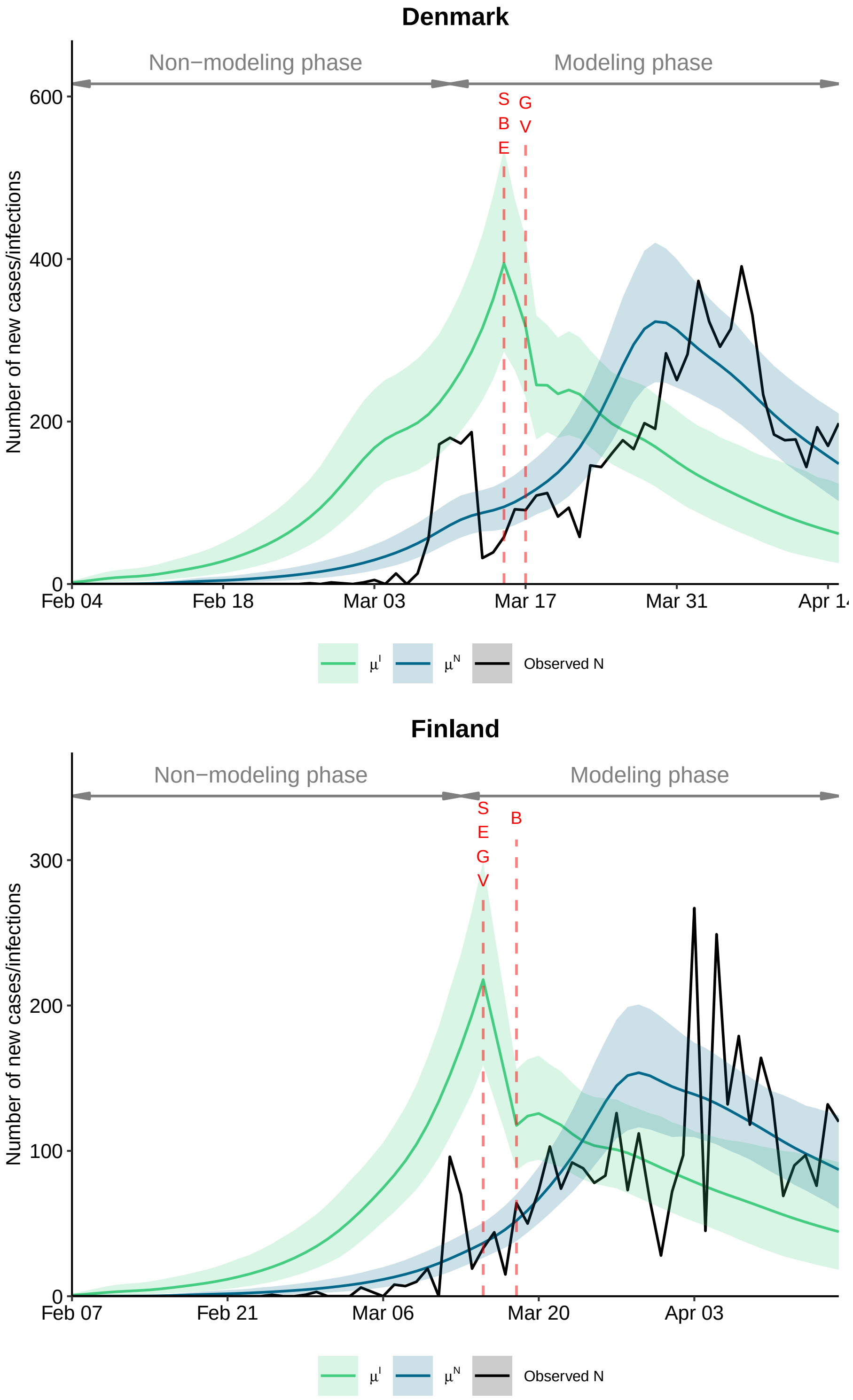

**Figure 17.** Expected number of new infections $\mu^I$ and new cases $\mu^N$ (posterior mean as colored lines with 95% credible interval as shaded area) and the observed number of new cases by country over time. Red letters and lines indicate the first day an NPI was implemented within a country (S: School closures, B: Border closure, E: Event ban, G: Gathering ban, V: Venue closure, H: Stay-at-home order, W: Work ban).

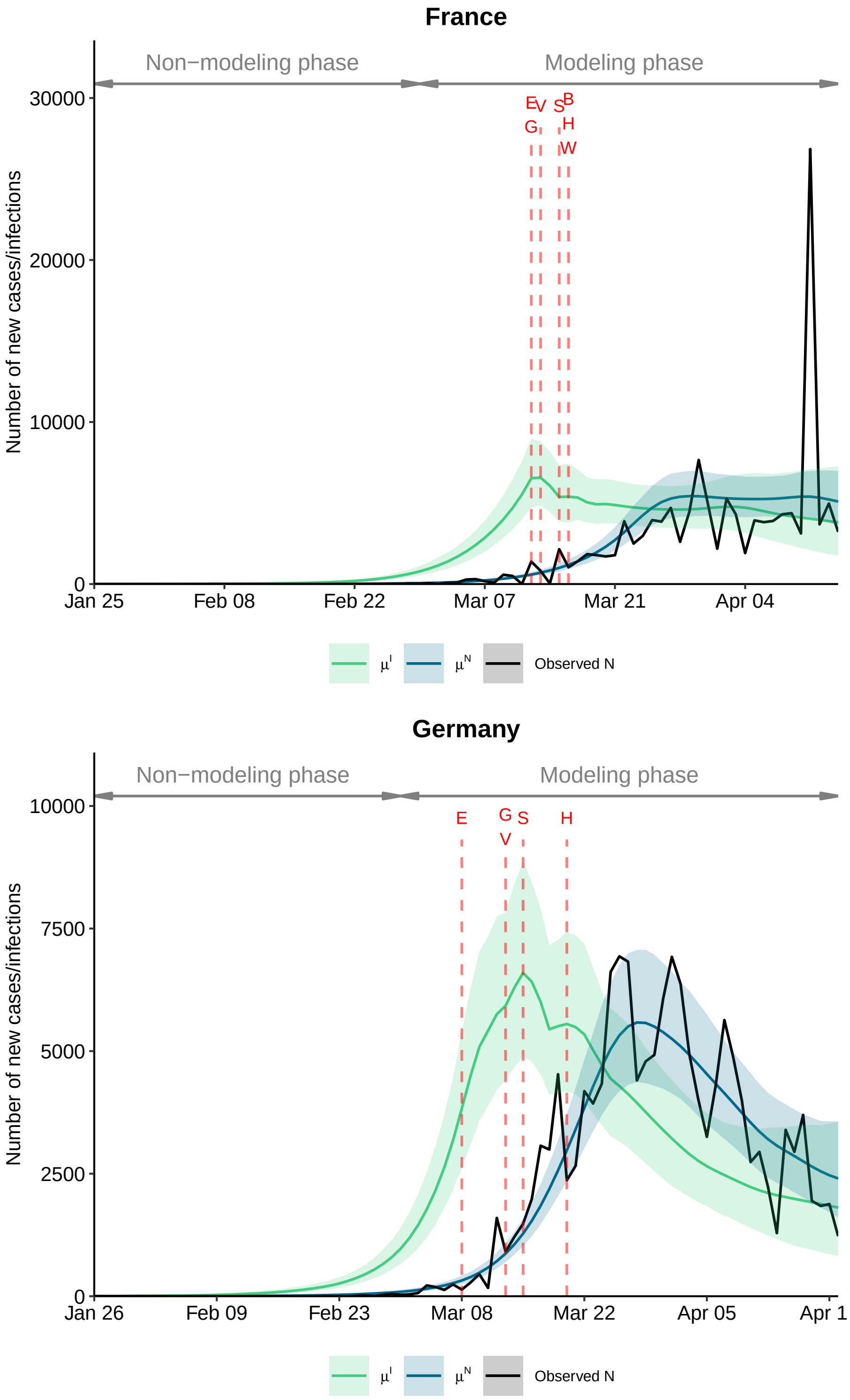

**Figure 17.** Expected number of new infections $\mu^I$ and new cases $\mu^N$ (posterior mean as colored lines with 95% credible interval as shaded area) and the observed number of new cases by country over time. Red letters and lines indicate the first day an NPI was implemented within a country (S: School closures, B: Border closure, E: Event ban, G: Gathering ban, V: Venue closure, H: Stay-at-home order, W: Work ban).

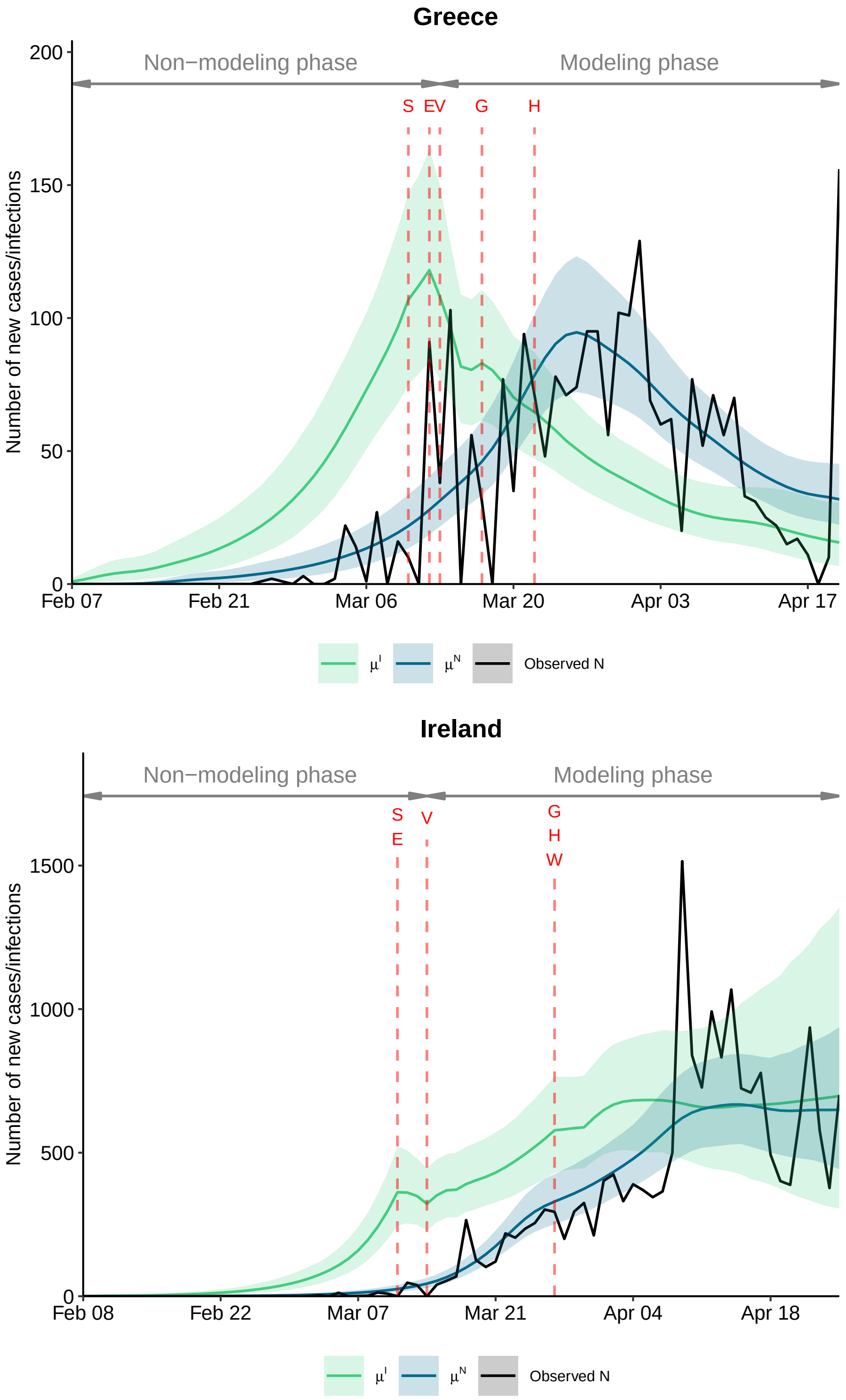

**Figure 17.** Expected number of new infections $\mu^I$ and new cases $\mu^N$ (posterior mean as colored lines with 95% credible interval as shaded area) and the observed number of new cases by country over time. Red letters and lines indicate the first day an NPI was implemented within a country (S: School closures, B: Border closure, E: Event ban, G: Gathering ban, V: Venue closure, H: Stay-at-home order, W: Work ban).

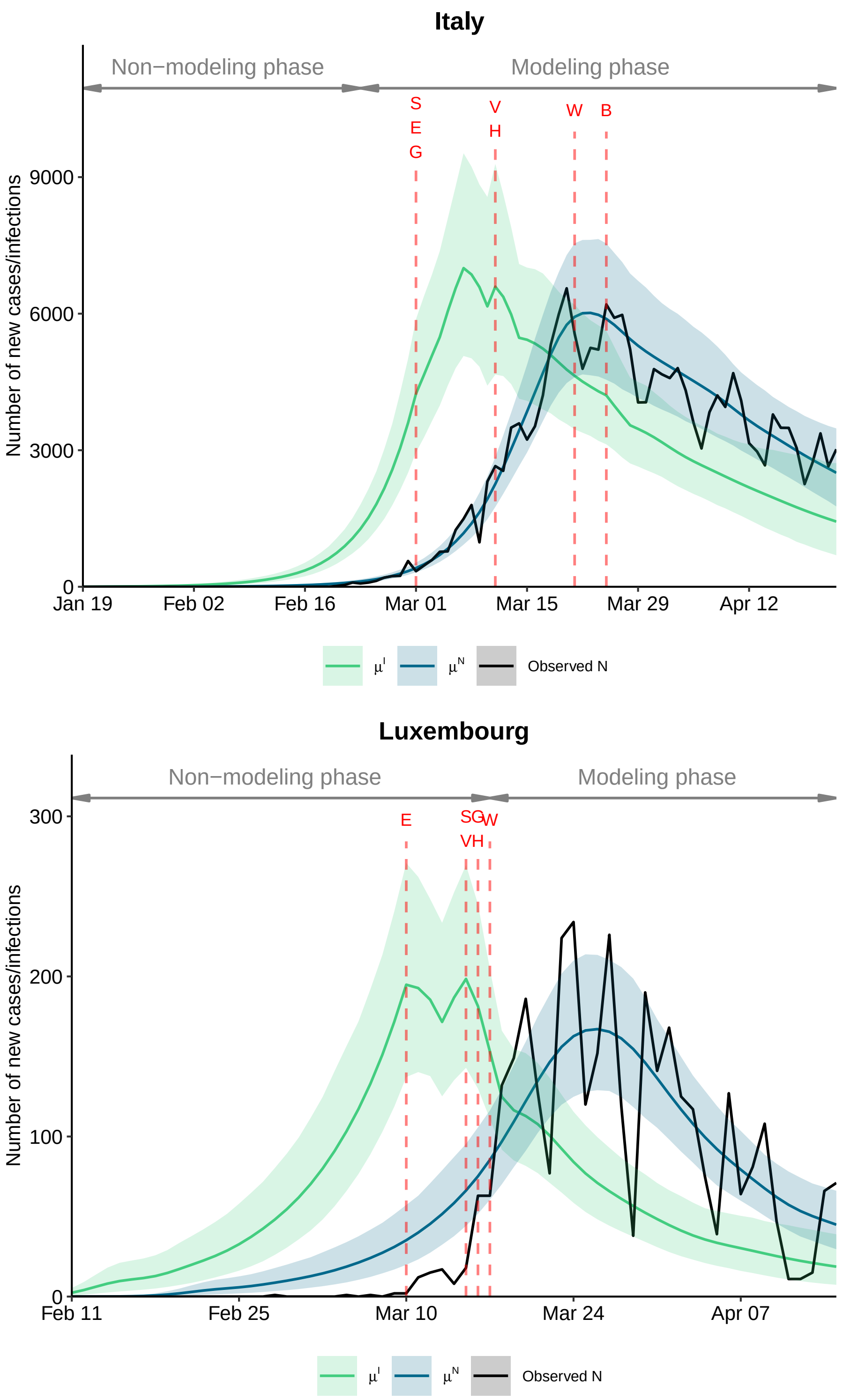

**Figure 17.** Expected number of new infections $\mu^I$ and new cases $\mu^N$ (posterior mean as colored lines with 95% credible interval as shaded area) and the observed number of new cases by country over time. Red letters and lines indicate the first day an NPI was implemented within a country (S: School closures, B: Border closure, E: Event ban, G: Gathering ban, V: Venue closure, H: Stay-at-home order, W: Work ban).

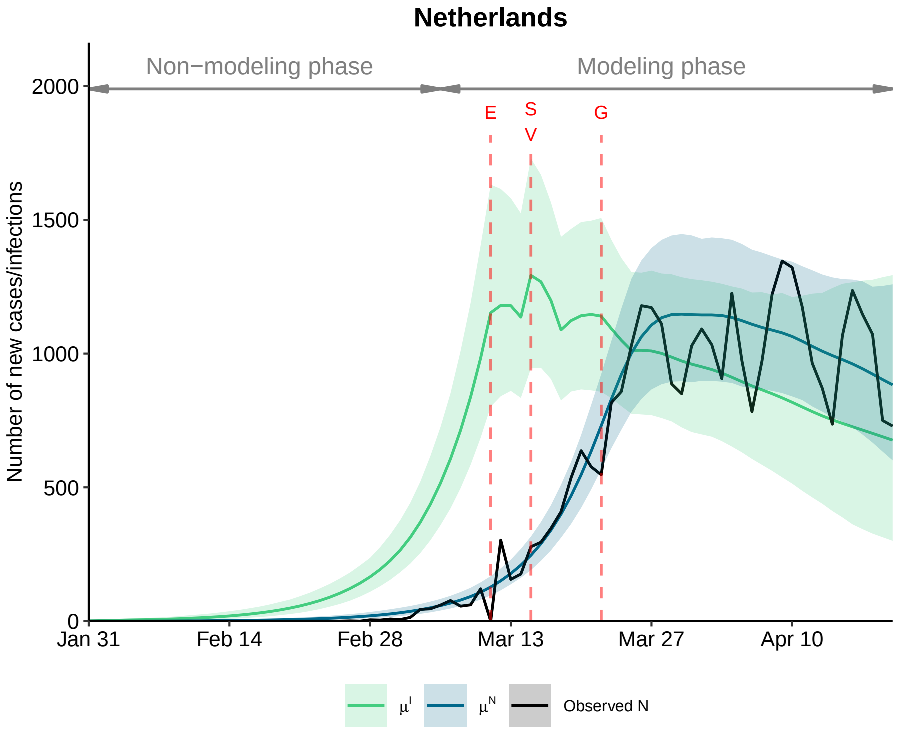

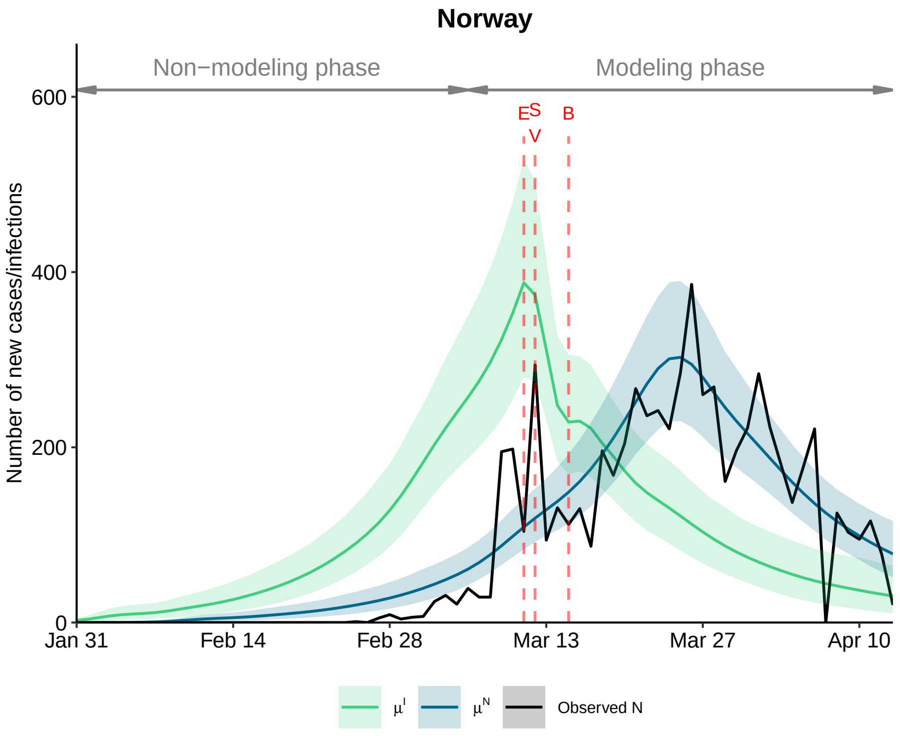

**Figure 17.** Expected number of new infections $\mu^I$ and new cases $\mu^N$ (posterior mean as colored lines with 95% credible interval as shaded area) and the observed number of new cases by country over time. Red letters and lines indicate the first day an NPI was implemented within a country (S: School closures, B: Border closure, E: Event ban, G: Gathering ban, V: Venue closure, H: Stay-at-home order, W: Work ban).

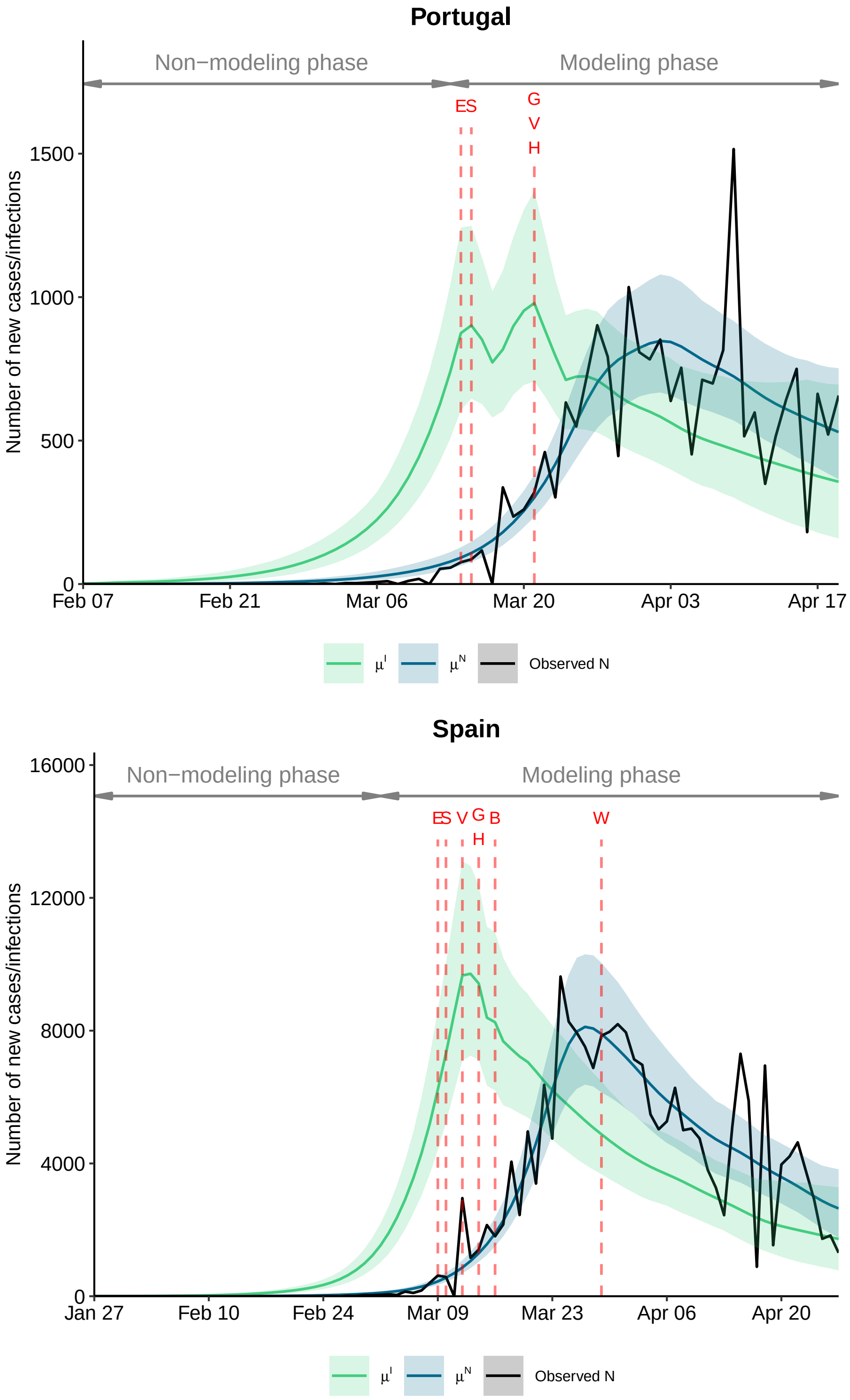

**Figure 17.** Expected number of new infections $\mu^I$ and new cases $\mu^N$ (posterior mean as colored lines with 95% credible interval as shaded area) and the observed number of new cases by country over time. Red letters and lines indicate the first day an NPI was implemented within a country (S: School closures, B: Border closure, E: Event ban, G: Gathering ban, V: Venue closure, H: Stay-at-home order, W: Work ban).

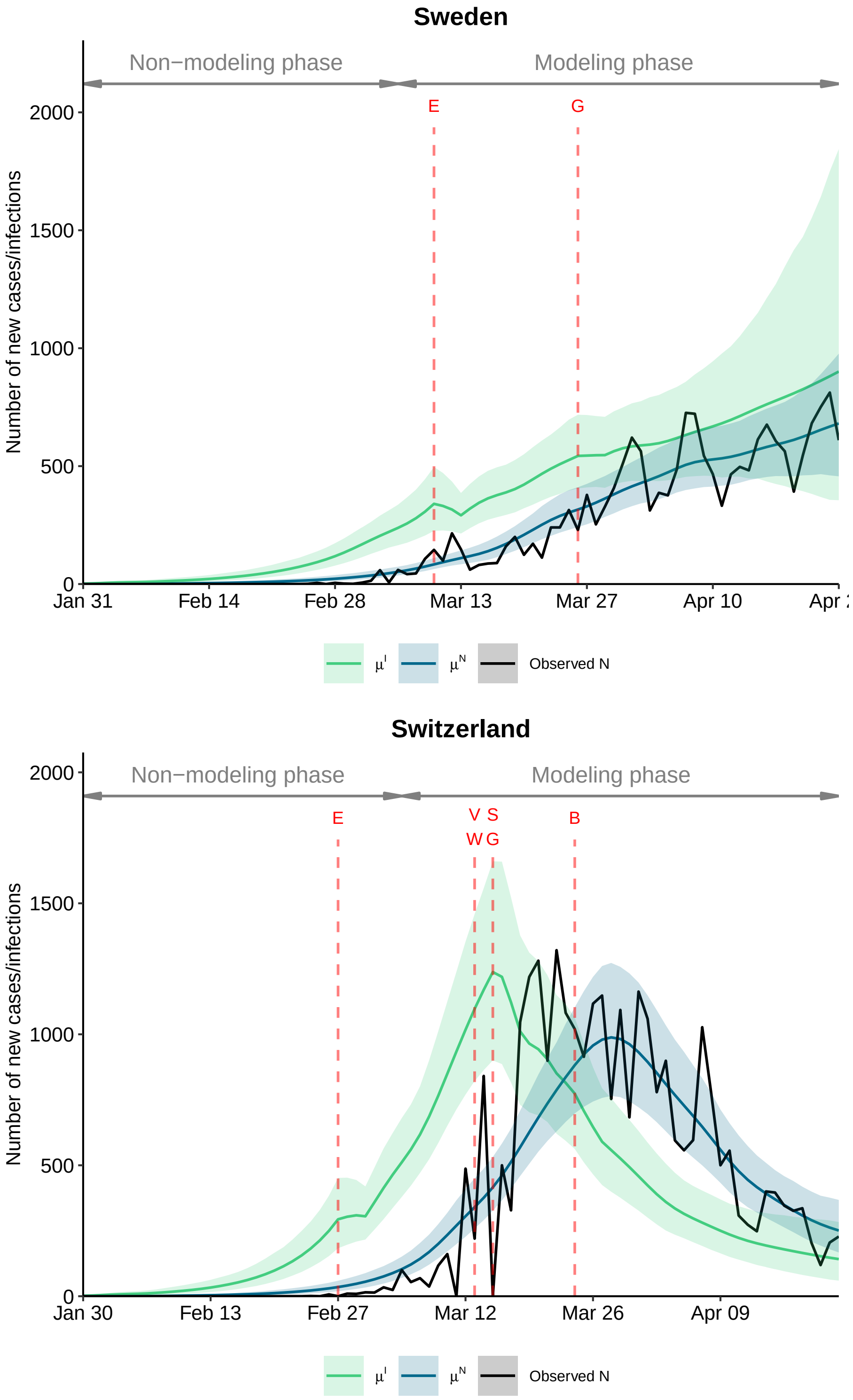

**Figure 17.** Expected number of new infections $\mu^I$ and new cases $\mu^N$ (posterior mean as colored lines with 95% credible interval as shaded area) and the observed number of new cases by country over time. Red letters and lines indicate the first day an NPI was implemented within a country (S: School closures, B: Border closure, E: Event ban, G: Gathering ban, V: Venue closure, H: Stay-at-home order, W: Work ban).

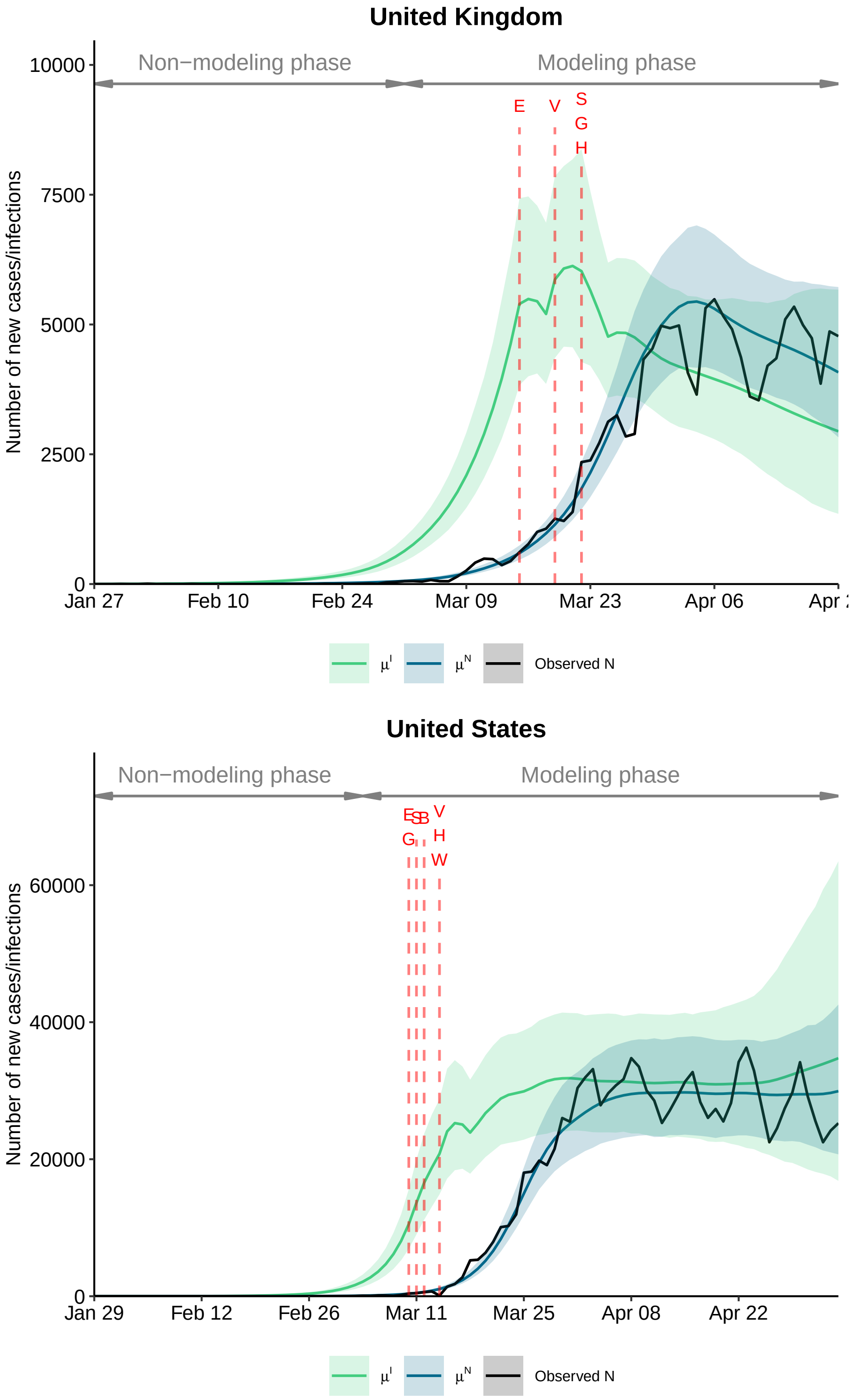

**Figure 17.** Expected number of new infections $\mu^I$ and new cases $\mu^N$ (posterior mean as colored lines with 95% credible interval as shaded area) and the observed number of new cases by country over time. Red letters and lines indicate the first day an NPI was implemented within a country (S: School closures, B: Border closure, E: Event ban, G: Gathering ban, V: Venue closure, H: Stay-at-home order, W: Work ban).

### 6 Data on non-pharmaceutical interventions

#### 6.1 Data collection

Data on non-pharmaceutical interventions (NPIs) have been collected systematically in six steps.

1. Granular information on NPIs have been gathered from government resources and news outlets by two authors (AC, PB).

2. After collecting data for the first few countries, NPIs have been classified into seven categories by seven authors (NB, EvW, AL, AS, DT, AC, PB): (1) school closures, (2) border closures, (3) public event bans, (4) gathering bans, (5) venue closures (e.g., shops, bars, restaurants, and other recreational activities), (6) stay-at-home orders prohibiting public movements without valid reason, and (7) work bans on non-essential business activities. The resulting classification has later been cross-checked against the encoding from the Imperial College COVID-19 Response Team[9] and "Coronavirus Government Response Tracker" from the University of Oxford[21].

3. The date of NPIs has been referred to as the first day a measure went into action. For instance, if a country banned events with more than 5,000 people on March 1 and events with more than 1,000 people on March 5, then March 1 has been chosen as the date of event bans.

4. NPIs for countries that subsequently followed in the data collection have been encoded accordingly.

5. The date of NPIs has been collected for each country or region.

6. A fifth author (BK) checked and verified the collected data. Part of this was also to recruit local residents and/or native speakers from each country in order to check our encoding. The reason for this is that countries have often used different legal terms to refer to the same NPI. Also, checking with local residents has helped to determine, for instance, whether the NPI was actually enforced or just recommended.

#### 6.2 Data Sources

In Tbl. 6, we provide sources of our data on NPIs at a national level. For less centrally managed countries, the aggregated (national level) date of the NPI is shown and the regional sources are displayed in Tbl. 7 (United States), Tbl. 8 (Germany), Tbl. 9 (Spain), Tbl. 10 (Canada), Tbl. 11 (Australia), and Tbl. 12 (Italy). Note that border closures are defined as a national closure of borders and is thus only considered at a national level.

In the following, we want to highlight some encoding decisions that were subject to internal discussions.

- School closure in New South Wales (Australia): Schools have remained technically open but attendance dropped below 5%. Therefore, schools have been encoded as closed.
- Border closure in the US: The US has not stopped flights to all countries but closed their land borders and has suspended travel from a huge number of Asian and European countries. Hence, US borders have been encoded as closed.
- Border closure in Germany: Germany has not closed its borders with Belgium and the Netherlands, therefore not all land borders have been closed and we have decided to encode them as open.
- School closure in Sweden: Only upper secondary schools (16+ y/o) and universities are studying from home, while other schools are still open. Thus, Swedish schools have been encoded as open.

| Country | NPI | Date effective | Source |
| --- | --- | --- | --- |
| Australia | Event ban | | Date derived by cumulative share, see Tab. 11 |
| | Gathering ban | | Date derived by cumulative share, see Tab. 11 |
| | Border closure | 2020-03-20 | https://www.pm.gov.au/media/border-restrictions |
| | Venue closure | | Date derived by cumulative share, see Tab. 11 |
| Austria | Event ban | 2020-03-11 | https://www.bundeskanzleramt.gv.at/bundeskanzleramt/nachrichten-der-bundesregierung/2020/weitere-massnahmen-gegen-ausbreitung-des-coronavirus.html |
| | Gathering ban | 2020-03-16 | https://www.reuters.com/article/us-health-coronavirus-austria/austria-imposes-major-restrictions-on-movement-over-coronavirus-idUSKBN2120D8 |
| | School closure | 2020-03-16 | https://www.reuters.com/article/us-health-coronavirus-austria/austria-closing-schools-over-coronavirus-as-border-checks-take-effect-idUSKBN20Y2YC |
| | Border closure | 2020-03-19 | https://www.reuters.com/article/us-health-coronavirus-austria/coronavirus-infections-top-2000-in-austria-more-border-controls-imposed-idUSKBN2160WK |
| | Venue closure | 2020-03-16 | https://www.bundeskanzleramt.gv.at/bundeskanzleramt/nachrichten-der-bundesregierung/2020/bundesregierung-praesentiert-aktuelle-beschluesse-zum-coronavirus.html |
| | Lockdown | 2020-03-16 | https://www.sozialministerium.at/Informationen-zum-Coronavirus/Coronavirus---Aktuelle-Ma%C3%9Fnahmen.html |
| Belgium | Event ban | 2020-03-10 | https://www.info-coronavirus.be/en/news/protect-yourself-and-protect-the-others/ |
| | Gathering ban | 2020-03-18 | https://de.reuters.com/article/health-coronavirus-belgium-lockdown-idUSB5N28S003 |

| | | | |
|---|---|---|---|
| | School closure | 2020-03-16 | https://www.info-coronavirus.be/en/2020/03/12/phase-2-maintained-transition-to-the-federal-phase-and-additional-measures/ |
| | Border closure | 2020-03-20 | https://www.politico.eu/article/belgium-closes-borders-for-non-essential-travel/ |
| | Venue closure | 2020-03-14 | https://www.info-coronavirus.be/en/2020/03/12/phase-2-maintained-transition-to-the-federal-phase-and-additional-measures/ |
| | Lockdown | 2020-03-18 | https://de.reuters.com/article/health-coronavirus-belgium-lockdown-idUSB5N28S003 |
| Canada | Event ban | | Date derived by cumulative share, see Tab. 10 |
| | Gathering ban | | Date derived by cumulative share, see Tab. 10 |
| | School closure | | Date derived by cumulative share, see Tab. 10 |
| | Border closure | 2020-03-21 | https://www.canada.ca/en/public-health/services/diseases/2019-novel-coronavirus-infection/canadas-reponse.html?topic=tilelink |
| | Venue closure | | Date derived by cumulative share, see Tab. 10 |
| Denmark | Event ban | 2020-03-16 | https://www.regeringen.dk/nyheder/pressemoede-11-marts-i-spejlsalen/ |
| | Gathering ban | 2020-03-18 | https://www.reuters.com/article/us-health-coronavirus-denmark/denmark-bans-crowds-of-over-10-people-to-curb-coronavirus-idUSKBN2143KG |
| | School closure | 2020-03-16 | https://www.regeringen.dk/nyheder/pressemoede-11-marts-i-spejlsalen/ |
| | Border closure | 2020-03-16 | https://www.oresunddirekt.dk/en/news/danish-borders-closed-due-to-coronavirus-covid-19 |
| | Venue closure | 2020-03-18 | https://www.reuters.com/article/us-health-coronavirus-denmark/denmark-bans-crowds-of-over-10-people-to-curb-coronavirus-idUSKBN2143KG |
| Finland | Event ban | 2020-03-16 | https://valtioneuvosto.fi/en/article/-/asset_publisher/10616/hallitus-totesi-suomen-olevan-poikkeusoloissa-koronavirustilanteen-vuoksi |

| | | | |
|---|---|---|---|
| | Gathering ban | 2020-03-16 | https://valtioneuvosto.fi/en/article/-/asset_publisher/10616/hallitus-totesi-suomen-olevan-poikkeusoloissa-koronavirustilanteen-vuoksi |
| | School closure | 2020-03-16 | https://valtioneuvosto.fi/en/article/-/asset_publisher/10616/hallitus-totesi-suomen-olevan-poikkeusoloissa-koronavirustilanteen-vuoksi |
| | Border closure | 2020-03-19 | https://valtioneuvosto.fi/en/article/-/asset_publisher/1410869/suomen-rajaliikennetta-aletaan-rajoittaa-elakkeella-olevia-rajavartijoita-ja-poliiseja-voidaan-kutsua-toihin |
| | Venue closure | 2020-03-16 | https://valtioneuvosto.fi/en/article/-/asset_publisher/10616/hallitus-totesi-suomen-olevan-poikkeusoloissa-koronavirustilanteen-vuoksi |
| France | Event ban | 2020-03-13 | https://www.bbc.com/news/world-europe-51892477 |
| | Gathering ban | 2020-03-13 | https://www.bbc.com/news/world-europe-51892477 |
| | School closure | 2020-03-16 | https://www.bbc.com/news/world-europe-51892477 |
| | Border closure | 2020-03-17 | https://www.gouvernement.fr/info-coronavirus |
| | Venue closure | 2020-03-14 | https://www.gouvernement.fr/partage/11444-declaration-de-m-edouard-philippe-premier-ministre-sur-le-covid-19 |
| | Lockdown | 2020-03-17 | https://www.gouvernement.fr/info-coronavirus |
| | Work ban | 2020-03-17 | https://www.gouvernement.fr/info-coronavirus |
| Germany | Event ban | | Date derived by cumulative share, see Tab. 8 |
| | Gathering ban | | Date derived by cumulative share, see Tab. 8 |
| | School closure | | Date derived by cumulative share, see Tab. 8 |
| | Venue closure | | Date derived by cumulative share, see Tab. 8 |
| Greece | Event ban | 2020-03-13 | https://www.reuters.com/article/us-health-coronavirus-greece-measures/greece-to-shut-shops-quarantine-all-arrivals-from-abroad-idUSKBN2131SF |
| | Gathering ban | 2020-03-18 | https://www.reuters.com/article/us-health-coronavirus-greece-curfew/greece-imposes-lockdown-after-coronavirus-infections-jump-idUSKBN2190Z1 |

| | | | |
|---|---|---|---|
| | School closure | 2020-03-11 | https://www.reuters.com/article/us-health-coronavirus-greece-education/greece-shuts-schools-universities-to-halt-coronavirus-spread-idUSKBN20X28V |
| | Venue closure | 2020-03-14 | https://www.cnn.gr/news/ellada/story/211153/koronoios-poia-katastimata-kleinoyn-poia-menoyn-anoikta-lista |
| | Lockdown | 2020-03-23 | https://www.reuters.com/article/us-health-coronavirus-greece-curfew/greece-imposes-lockdown-after-coronavirus-infections-jump-idUSKBN2190Z1 |
| Ireland | Event ban | 2020-03-12 | https://www.gov.ie/en/press-release/96eb4c-statement-from-the-national-public-health-emergency-team/ |
| | Gathering ban | 2020-03-28 | https://www.gov.ie/en/publication/539d23-stay-at-home-the-latest-public-health-measures-to-prevent-the-spread/ |
| | School closure | 2020-03-12 | https://www.gov.ie/en/press-release/96eb4c-statement-from-the-national-public-health-emergency-team/ |
| | Venue closure | 2020-03-15 | https://www.gov.ie/en/press-release/20fc58-all-pubs-advised-to-close-until-march-29/ |
| | Lockdown | 2020-03-28 | https://www.gov.ie/en/publication/539d23-stay-at-home-the-latest-public-health-measures-to-prevent-the-spread/ |
| | Work ban | 2020-03-28 | https://www.gov.ie/en/publication/539d23-stay-at-home-the-latest-public-health-measures-to-prevent-the-spread/ |
| Italy | Event ban | | Date derived by cumulative share, see Tab. 12 |
| | Gathering ban | | Date derived by cumulative share, see Tab. 12 |
| | School closure | | Date derived by cumulative share, see Tab. 12 |
| | Border closure | 2020-03-26 | https://www.gazzettaufficiale.it/showNewsDetail?id=2553&backTo=archivio&anno=2020&provenienza=archivio |
| | Venue closure | | Date derived by cumulative share, see Tab. 12 |

| | | | |
|---|---|---|---|
| | Lockdown | | Date derived by cumulative share, see Tab. 12 |
| | Work ban | | Date derived by cumulative share, see Tab. 12 |
| Luxembourg | Event ban | 2020-03-11 | https://gouvernement.lu/fr/actualites/toutes_actualites/communiques/2020/03-mars/11-conseil-gouvernement.html |
| | Gathering ban | 2020-03-17 | https://coronavirus.gouvernement.lu/en/communications-officielles.gouvernement%2Ben%2Bactualites%2Btoutes_actualites%2Bcommuniques%2B2020%2B03-mars%2B17-declaration-premier-chd.html |
| | School closure | 2020-03-16 | https://coronavirus.gouvernement.lu/en/communications-officielles.gouvernement%2Ben%2Bactualites%2Btoutes_actualites%2Bcommuniques%2B2020%2B03-mars%2B12-cdg-extraordinaire-coronavirus.html |
| | Venue closure | 2020-03-16 | https://coronavirus.gouvernement.lu/en/communications-officielles.gouvernement%2Ben%2Bactualites%2Btoutes_actualites%2Bcommuniques%2B2020%2B03-mars%2B15-nouvelles-mesures-coronavirus.html |
| | Lockdown | 2020-03-17 | https://coronavirus.gouvernement.lu/en/communications-officielles.gouvernement%2Ben%2Bactualites%2Btoutes_actualites%2Bcommuniques%2B2020%2B03-mars%2B17-declaration-premier-chd.html |
| | Work ban | 2020-03-18 | http://www.legilux.lu/eli/etat/leg/rgd/2020/03/18/a165/jo |
| Netherlands | Event ban | 2020-03-12 | https://www.government.nl/latest/news/2020/03/12/new-measures-to-stop-spread-of-coronavirus-in-the-netherlands |
| | Gathering ban | 2020-03-23 | https://www.rijksoverheid.nl/actueel/nieuws/2020/03/24/aanvullende-maatregelen-23-maart |
| | School closure | 2020-03-16 | https://www.government.nl/latest/news/2020/03/15/additional-measures-in-schools-the-hospitality-sector-and-sport |
| | Venue closure | 2020-03-16 | https://www.government.nl/latest/news/2020/03/15/additional-measures-in-schools-the-hospitality-sector-and-sport |

| | | | |
|---|---|---|---|
| Norway | Event ban | 2020-03-12 | https://www.fhi.no/nettpub/coronavirus/rad-og-informasjon-til-andre-sektorer-og-yrkesgrupper/anbefalinger-ved--store-arrangementer-knyttet-til-koronasmitte-i-norge/ |
| | School closure | 2020-03-13 | https://www.regjeringen.no/en/aktuelt/coronavirus-measures-to-continue/id2694682/ |
| | Border closure | 2020-03-16 | https://www.regjeringen.no/no/aktuelt/innforer-strengere-grensekontroll/id2693624/ |
| | Venue closure | 2020-03-13 | https://www.regjeringen.no/en/aktuelt/coronavirus-measures-to-continue/id2694682/ |
| Portugal | Event ban | 2020-03-15 | https://dre.pt/web/guest/home/-/dre/130277342/details/maximized |
| | Gathering ban | 2020-03-22 | https://dre.pt/web/guest/legislacao-consolidada/-/lc/130473378/202004042116/73800717/diploma/indice |
| | School closure | 2020-03-16 | https://www.portugal.gov.pt/pt/gc22/comunicacao/comunicado?i=suspensao-de-todas-as-atividades-letivas-e-nao-letivas-com-presenca-de-estudantes-em-todas-as-instituicoes-de-ensino-superior |
| | Venue closure | 2020-03-22 | https://dre.pt/web/guest/legislacao-consolidada/-/lc/130473378/202004042116/73800717/diploma/indice |
| | Lockdown | 2020-03-22 | https://dre.pt/web/guest/legislacao-consolidada/-/lc/130473378/202004042116/73800717/diploma/indice |
| Spain | Event ban | | Date derived by cumulative share, see Tab. 9 |
| | Gathering ban | | Date derived by cumulative share, see Tab. 9 |
| | School closure | | Date derived by cumulative share, see Tab. 9 |
| | Border closure | 2020-03-17 | https://english.elpais.com/society/2020-03-16/spain-closes-its-borders-to-contain-coronavirus.html |
| | Venue closure | | Date derived by cumulative share, see Tab. 9 |
| | Lockdown | | Date derived by cumulative share, see Tab. 9 |

| | | | |
|---|---|---|---|
| | Work ban | | Date derived by cumulative share, see Tab. 9 |
| Sweden | Event ban | 2020-03-11 | https://www.government.se/articles/2020/03/ordinance-on-a-prohibition-against-holding-public-gatherings-and-events/ |
| | Gathering ban | 2020-03-27 | https://www.dailymail.co.uk/news/article-8160653/Sweden-bans-gatherings-50-people-threatens-people-six-month-jail-terms.html |
| Switzerland | Event ban | | Date derived by cumulative share, see Tab. 13 |
| | Gathering ban | | Date derived by cumulative share, see Tab. 13 |
| | School closure | | Date derived by cumulative share, see Tab. 13 |
| | Border closure | 2020-03-25 | https://www.bag.admin.ch/bag/it/home/krankheiten/ausbrueche-epidemien-pandemien/aktuelle-ausbrueche-epidemien/novel-cov/massnahmen-des-bundes.html |
| | Venue closure | | Date derived by cumulative share, see Tab. 13 |
| United Kingdom | Event ban | 2020-03-16 | https://www.gov.uk/guidance/covid-19-guidance-for-mass-gatherings |
| | Gathering ban | 2020-03-23 | https://www.gov.uk/government/speeches/pm-address-to-the-nation-on-coronavirus-23-march-2020 |
| | School closure | 2020-03-23 | https://www.gov.uk/government/speeches/pm-statement-on-coronavirus-22-march-2020 |
| | Venue closure | 2020-03-20 | https://www.wsj.com/articles/u-k-escalates-measures-to-fight-coronavirus-11584741690 |
| | Lockdown | 2020-03-23 | https://www.gov.uk/government/speeches/pm-address-to-the-nation-on-coronavirus-23-march-2020 |
| United States of America | Event ban | | Date derived by cumulative share, see Tab. 7 |
| | Gathering ban | | Date derived by cumulative share, see Tab. 7 |
| | School closure | | Date derived by cumulative share, see Tab. 7 |
| | Border closure | 2020-03-13 | https://www.theguardian.com/world/2020/mar/11/coronavirus-outbreak-us-trump-latest#maincontent |

| | |
|---|---|
| Venue closure | Date derived by cumulative share, see Tab. 7 |
| Lockdown | Date derived by cumulative share, see Tab. 7 |
| Work ban | Date derived by cumulative share, see Tab. 7 |

**Table 6.** Sources for policies in Switzerland, Austria, Belgium, Denmark, Finland, France, United Kingdom, Greece, Ireland, Luxembourg, Netherlands, Norway, Portugal, Sweden, United States of America, Germany, Italy, Spain, Canada, and Australia

| NPI | Date | Cumulative share | Region | Source |
|---|---|---|---|---|
| Event ban | 2020-03-11 | 0.02 | Rhode Island | https://www.ri.gov/press/view/37892 |
| Event ban | 2020-03-11 | 0.02 | Kentucky | https://governor.ky.gov/covid19 |
| Event ban | 2020-03-11 | 0.02 | District of Columbia | https://wjla.com/news/coronavirus/dc-health-postponing-cancelling-events-1000-people-through-march |
| Event ban | 2020-03-12 | 0.48 | Maryland | https://governor.maryland.gov/2020/03/12/governor-hogan-announces-major-actions-to-protect-public-health-limit-spread-of-covid-19-pandemic/ |
| Event ban | 2020-03-12 | 0.48 | Pennsylvania | https://lancasteronline.com/news/health/gov-tom-wolf-cancel-large-events-with-more-than-250-people/article_1a6f027c-648e-11ea-aa2f-c787d0825889.html |
| Event ban | 2020-03-12 | 0.48 | New York | https://www.governor.ny.gov/news/no-2021-continuing-temporary-suspension-and-modification-laws-relating-disaster-emergency |
| Event ban | 2020-03-12 | 0.48 | Oregon | https://www.oregon.gov/osp/programs/sfm/Pages/Event_Cancellations.aspx |
| Event ban | 2020-03-12 | 0.48 | Indiana | https://www.cdc.gov/coronavirus/2019-ncov/community/large-events/mass-gatherings-ready-for-covid-19.html |
| Event ban | 2020-03-12 | 0.48 | Ohio | https://coronavirus.ohio.gov/wps/wcm/connect/gov/b815ab52-a571-4e65-9077-32468779671a/ODH+Order+to+Limit+and+Prohibit+Mass+Gatherings%2C+3.12.20.pdf?MOD=AJPERES&CONVERT_TO=url&CACHEID=ROOTWORKSPACE.Z18_M1HGGIK0N0JO00QO9DDDDM3000-b815ab52-a571-4e65-9077-32468779671a-n5828iN |
| Event ban | 2020-03-12 | 0.48 | New Jersey | https://www.nj.gov/governor/news/news/562020/approved/20200312a.shtml |
| Event ban | 2020-03-12 | 0.48 | Mississippi | https://www.coronavirus.ms.gov/2019-11/msdh-has-issued-enhanced-protective-recommendations |
| Event ban | 2020-03-12 | 0.48 | Florida | https://www.miamiherald.com/news/local/community/miami-dade/article241133076.html |

| | | | | |
|---|---|---|---|---|
| Event ban | 2020-03-12 | 0.48 | New Mexico | https://www.governor.state.nm.us/2020/03/12/health-secretary-issues-public-health-order-suspending-mass-gatherings-in-new-mexico/ |
| Event ban | 2020-03-12 | 0.48 | Connecticut | https://portal.ct.gov/-/media/Office-of-the-Governor/Executive-Orders/Lamont-Executive-Orders/Executive-Order-No-7.pdf?la=en |
| Event ban | 2020-03-12 | 0.48 | Virginia | https://www.governor.virginia.gov/newsroom/all-releases/2020/march/headline-853537-en.html |
| Event ban | 2020-03-12 | 0.48 | California | https://thehill.com/policy/healthcare/487179-california-governor-calls-for-cancellations-of-large-events |
| Event ban | 2020-03-12 | 0.48 | West Virginia | https://governor.wv.gov/News/press-releases/2020/Pages/COVID-UPDATE-Gov.-Justice-announces-State-employee-travel-ban,-basketball-tournament-cancellation-among-latest-precautions.aspx |
| Event ban | 2020-03-12 | 0.48 | Utah | https://kutv.com/news/local/live-blog-closures-event-cancellations-more-in-utah-due-to-coronavirus |
| Event ban | 2020-03-13 | 0.63 | Minnesota | https://apnews.com/24d6d0c93ded9e8a5c2712df0a045854 |
| Event ban | 2020-03-13 | 0.63 | Washington | https://www.governor.wa.gov/news-media/inslee-announces-statewide-school-closures-expansion-limits-large-gatherings |
| Event ban | 2020-03-13 | 0.63 | Louisiana | https://gov.louisiana.gov/index.cfm/communication/viewcampaign/2548?&uid=hgdtfgl%3Dn7&nowrap=1 |
| Event ban | 2020-03-13 | 0.63 | Arizona | https://www.fox10phoenix.com/news/arizona-governor-says-schools-mass-gatherings-events-of-50-or-more-canceled-amid-covid-19-spread |
| Event ban | 2020-03-13 | 0.63 | Massachusetts | https://www.mass.gov/news/governor-baker-issues-order-limiting-large-gatherings-in-the-commonwealth |
| Event ban | 2020-03-13 | 0.63 | Michigan | https://www.michigan.gov/whitmer/0,9309,7-387-90499_90705-521595--,00.html |
| Event ban | 2020-03-13 | 0.63 | Tennessee | https://www.tn.gov/governor/news/2020/3/13/governor-lee-issues-guidance-for-mass-gatherings--schools-and-state-workforce.html |

| | | | | |
|---|---|---|---|---|
| Event ban | 2020-03-14 | 0.66 | North Carolina | https://files.nc.gov/governor/documents/files/EO117-COVID-19-Prohibiting-Mass-Gathering-and-K12-School-Closure.pdf |
| Event ban | 2020-03-15 | 0.67 | Puerto Rico | https://www.estado.pr.gov/es/ordenes-ejecutivas/ |
| Event ban | 2020-03-16 | 1.00 | South Carolina | https://eu.usatoday.com/story/news/health/2020/03/16/coronavirus-live-updates-us-death-toll-rises-cases-testing/5053816002/ |
| Event ban | 2020-03-16 | 1.00 | Oklahoma | https://eu.usatoday.com/story/news/health/2020/03/16/coronavirus-live-updates-us-death-toll-rises-cases-testing/5053816002/ |
| Event ban | 2020-03-16 | 1.00 | South Dakota | https://eu.usatoday.com/story/news/health/2020/03/16/coronavirus-live-updates-us-death-toll-rises-cases-testing/5053816002/ |
| Event ban | 2020-03-16 | 1.00 | North Dakota | https://eu.usatoday.com/story/news/health/2020/03/16/coronavirus-live-updates-us-death-toll-rises-cases-testing/5053816002/ |
| Event ban | 2020-03-16 | 1.00 | Texas | https://eu.usatoday.com/story/news/health/2020/03/16/coronavirus-live-updates-us-death-toll-rises-cases-testing/5053816002/ |
| Event ban | 2020-03-16 | 1.00 | Vermont | https://eu.usatoday.com/story/news/health/2020/03/16/coronavirus-live-updates-us-death-toll-rises-cases-testing/5053816002/ |
| Event ban | 2020-03-16 | 1.00 | Alabama | https://eu.usatoday.com/story/news/health/2020/03/16/coronavirus-live-updates-us-death-toll-rises-cases-testing/5053816002/ |
| Event ban | 2020-03-16 | 1.00 | Missouri | https://governor.mo.gov/press-releases/archive/governor-parsons-statement-regarding-cdc-recommendations-mass-gatherings-and |
| Event ban | 2020-03-16 | 1.00 | Nevada | https://eu.usatoday.com/story/news/health/2020/03/16/coronavirus-live-updates-us-death-toll-rises-cases-testing/5053816002/ |
| Event ban | 2020-03-16 | 1.00 | Alaska | https://eu.usatoday.com/story/news/health/2020/03/16/coronavirus-live-updates-us-death-toll-rises-cases-testing/5053816002/ |
| Event ban | 2020-03-16 | 1.00 | Arkansas | https://eu.usatoday.com/story/news/health/2020/03/16/coronavirus-live-updates-us-death-toll-rises-cases-testing/5053816002/ |

| | | | | |
|---|---|---|---|---|
| Event ban | 2020-03-16 | 1.00 | Colorado | https://northglenn-thorntonsentinel.com/stories/colorado-events-of-50-people-no-sit-down-restaurants-bars,296332 |
| Event ban | 2020-03-16 | 1.00 | Delaware | https://eu.usatoday.com/story/news/health/2020/03/16/coronavirus-live-updates-us-death-toll-rises-cases-testing/5053816002/ |
| Event ban | 2020-03-16 | 1.00 | Georgia | https://eu.usatoday.com/story/news/health/2020/03/16/coronavirus-live-updates-us-death-toll-rises-cases-testing/5053816002/ |
| Event ban | 2020-03-16 | 1.00 | Hawaii | https://eu.usatoday.com/story/news/health/2020/03/16/coronavirus-live-updates-us-death-toll-rises-cases-testing/5053816002/ |
| Event ban | 2020-03-16 | 1.00 | New Hampshire | https://www.governor.nh.gov/news-media/press-2020/20200316-covid-10-businesses.htm |
| Event ban | 2020-03-16 | 1.00 | Idaho | https://eu.usatoday.com/story/news/health/2020/03/16/coronavirus-live-updates-us-death-toll-rises-cases-testing/5053816002/ |
| Event ban | 2020-03-16 | 1.00 | Iowa | https://eu.usatoday.com/story/news/health/2020/03/16/coronavirus-live-updates-us-death-toll-rises-cases-testing/5053816002/ |
| Event ban | 2020-03-16 | 1.00 | Kansas | https://govstatus.egov.com/coronavirus |
| Event ban | 2020-03-16 | 1.00 | Maine | https://eu.usatoday.com/story/news/health/2020/03/16/coronavirus-live-updates-us-death-toll-rises-cases-testing/5053816002/ |
| Event ban | 2020-03-16 | 1.00 | Wisconsin | https://content.govdelivery.com/accounts/WIGOV/bulletins/2817964 |
| Event ban | 2020-03-16 | 1.00 | Montana | https://eu.usatoday.com/story/news/health/2020/03/16/coronavirus-live-updates-us-death-toll-rises-cases-testing/5053816002/ |
| Event ban | 2020-03-16 | 1.00 | Nebraska | https://eu.usatoday.com/story/news/health/2020/03/16/coronavirus-live-updates-us-death-toll-rises-cases-testing/5053816002/ |
| Event ban | 2020-03-16 | 1.00 | Illinois | https://herald-review.com/news/state-and-regional/govt-and-politics/monday-update-pritzker-bans-gatherings-of-50-or-more-12-new-illinois-cases-announced/article_c5fa6e55-70f4-583b-aea9-09ce2315eb16.html |

| | | | | |
|---|---|---|---|---|
| Event ban | 2020-03-16 | 1.00 | Wyoming | https://eu.usatoday.com/story/news/health/2020/03/16/coronavirus-live-updates-us-death-toll-rises-cases-testing/5053816002/ |

| NPI | Date | Cumulative share | Region | Source |
|---|---|---|---|---|
| Gathering ban | 2020-03-11 | 0.01 | Kentucky | https://governor.ky.gov/covid19 |
| Gathering ban | 2020-03-13 | 0.04 | Arizona | https://www.fox10phoenix.com/news/arizona-governor-says-schools-mass-gatherings-events-of-50-or-more-canceled-amid-covid-19-spread |
| Gathering ban | 2020-03-15 | 0.05 | Puerto Rico | https://www.estado.pr.gov/es/ordenes-ejecutivas/ |
| Gathering ban | 2020-03-16 | 1.00 | New Hampshire | https://www.governor.nh.gov/news-media/press-2020/20200316-covid-10-businesses.htm |
| Gathering ban | 2020-03-16 | 1.00 | New Jersey | https://www.governor.nh.gov/news-media/press-2020/20200315-emergency-order-1.html |
| Gathering ban | 2020-03-16 | 1.00 | New Mexico | https://eu.usatoday.com/story/news/health/2020/03/16/coronavirus-live-updates-us-death-toll-rises-cases-testing/5053816002/ |
| Gathering ban | 2020-03-16 | 1.00 | New York | https://eu.usatoday.com/story/news/health/2020/03/16/coronavirus-live-updates-us-death-toll-rises-cases-testing/5053816002/ |
| Gathering ban | 2020-03-16 | 1.00 | North Carolina | https://eu.usatoday.com/story/news/health/2020/03/16/coronavirus-live-updates-us-death-toll-rises-cases-testing/5053816002/ |
| Gathering ban | 2020-03-16 | 1.00 | North Dakota | https://eu.usatoday.com/story/news/health/2020/03/16/coronavirus-live-updates-us-death-toll-rises-cases-testing/5053816002/ |
| Gathering ban | 2020-03-16 | 1.00 | Ohio | https://eu.usatoday.com/story/news/health/2020/03/16/coronavirus-live-updates-us-death-toll-rises-cases-testing/5053816002/ |
| Gathering ban | 2020-03-16 | 1.00 | Oklahoma | https://eu.usatoday.com/story/news/health/2020/03/16/coronavirus-live-updates-us-death-toll-rises-cases-testing/5053816002/ |

| | | | | |
|---|---|---|---|---|
| Gathering ban | 2020-03-16 | 1.00 | Oregon | https://www.kptv.com/news/covid--in-oregon-governor-cancels-gatherings-of-more-than/article_8e9aafec-67d2-11ea-af7f-2bad23d52443.html |
| Gathering ban | 2020-03-16 | 1.00 | Pennsylvania | https://www.governor.pa.gov/newsroom/gov-wolf-puts-statewide-covid-19-mitigation-efforts-in-effect-stresses-need-for-every-pennsylvanian-to-take-action-to-stop-the-spread/ |
| Gathering ban | 2020-03-16 | 1.00 | Alabama | https://governor.alabama.gov/assets/2020/03/Amended-Statewide-Social-Distancing-SHO-Order-3.27.2020-FINAL.pdf |
| Gathering ban | 2020-03-16 | 1.00 | Rhode Island | https://eu.usatoday.com/story/news/health/2020/03/16/coronavirus-live-updates-us-death-toll-rises-cases-testing/5053816002/ |
| Gathering ban | 2020-03-16 | 1.00 | South Carolina | https://eu.usatoday.com/story/news/health/2020/03/16/coronavirus-live-updates-us-death-toll-rises-cases-testing/5053816002/ |
| Gathering ban | 2020-03-16 | 1.00 | South Dakota | https://eu.usatoday.com/story/news/health/2020/03/16/coronavirus-live-updates-us-death-toll-rises-cases-testing/5053816002/ |
| Gathering ban | 2020-03-16 | 1.00 | Tennessee | https://eu.usatoday.com/story/news/health/2020/03/16/coronavirus-live-updates-us-death-toll-rises-cases-testing/5053816002/ |
| Gathering ban | 2020-03-16 | 1.00 | Texas | https://eu.usatoday.com/story/news/health/2020/03/16/coronavirus-live-updates-us-death-toll-rises-cases-testing/5053816002/ |
| Gathering ban | 2020-03-16 | 1.00 | Utah | https://eu.usatoday.com/story/news/health/2020/03/16/coronavirus-live-updates-us-death-toll-rises-cases-testing/5053816002/ |
| Gathering ban | 2020-03-16 | 1.00 | Vermont | https://eu.usatoday.com/story/news/health/2020/03/16/coronavirus-live-updates-us-death-toll-rises-cases-testing/5053816002/ |
| Gathering ban | 2020-03-16 | 1.00 | Virginia | https://eu.usatoday.com/story/news/health/2020/03/16/coronavirus-live-updates-us-death-toll-rises-cases-testing/5053816002/ |
| Gathering ban | 2020-03-16 | 1.00 | Washington | https://eu.usatoday.com/story/news/health/2020/03/16/coronavirus-live-updates-us-death-toll-rises-cases-testing/5053816002/ |

| | | | | |
|---|---|---|---|---|
| Gathering ban | 2020-03-16 | 1.00 | West Virginia | https://eu.usatoday.com/story/news/health/2020/03/16/coronavirus-live-updates-us-death-toll-rises-cases-testing/5053816002/ |
| Gathering ban | 2020-03-16 | 1.00 | Nevada | https://eu.usatoday.com/story/news/health/2020/03/16/coronavirus-live-updates-us-death-toll-rises-cases-testing/5053816002/ |
| Gathering ban | 2020-03-16 | 1.00 | Nebraska | https://eu.usatoday.com/story/news/health/2020/03/16/coronavirus-live-updates-us-death-toll-rises-cases-testing/5053816002/ |
| Gathering ban | 2020-03-16 | 1.00 | Missouri | https://eu.usatoday.com/story/news/health/2020/03/16/coronavirus-live-updates-us-death-toll-rises-cases-testing/5053816002/ |
| Gathering ban | 2020-03-16 | 1.00 | Wisconsin | https://content.govdelivery.com/accounts/WIGOV/bulletins/2817964 |
| Gathering ban | 2020-03-16 | 1.00 | Alaska | https://eu.usatoday.com/story/news/health/2020/03/16/coronavirus-live-updates-us-death-toll-rises-cases-testing/5053816002/ |
| Gathering ban | 2020-03-16 | 1.00 | Arkansas | https://eu.usatoday.com/story/news/health/2020/03/16/coronavirus-live-updates-us-death-toll-rises-cases-testing/5053816002/ |
| Gathering ban | 2020-03-16 | 1.00 | California | https://eu.usatoday.com/story/news/health/2020/03/16/coronavirus-live-updates-us-death-toll-rises-cases-testing/5053816002/ |
| Gathering ban | 2020-03-16 | 1.00 | Colorado | https://northglenn-thorntonsentinel.com/stories/colorado-events-of-50-people-no-sit-down-restaurants-bars,296332 |
| Gathering ban | 2020-03-16 | 1.00 | Connecticut | https://eu.usatoday.com/story/news/health/2020/03/16/coronavirus-live-updates-us-death-toll-rises-cases-testing/5053816002/ |
| Gathering ban | 2020-03-16 | 1.00 | Delaware | https://eu.usatoday.com/story/news/health/2020/03/16/coronavirus-live-updates-us-death-toll-rises-cases-testing/5053816002/ |
| Gathering ban | 2020-03-16 | 1.00 | District of Columbia | https://dc.gov/release/mayor%E2%80%99s-order-2020-048-prohibition-mass-gatherings-during-public-health-emergency |
| Gathering ban | 2020-03-16 | 1.00 | Florida | https://eu.usatoday.com/story/news/health/2020/03/16/coronavirus-live-updates-us-death-toll-rises-cases-testing/5053816002/ |

| | | | | |
|---|---|---|---|---|
| Gathering ban | 2020-03-16 | 1.00 | Georgia | https://eu.usatoday.com/story/news/health/2020/03/16/coronavirus-live-updates-us-death-toll-rises-cases-testing/5053816002/ |
| Gathering ban | 2020-03-16 | 1.00 | Hawaii | https://eu.usatoday.com/story/news/health/2020/03/16/coronavirus-live-updates-us-death-toll-rises-cases-testing/5053816002/ |
| Gathering ban | 2020-03-16 | 1.00 | Montana | https://eu.usatoday.com/story/news/health/2020/03/16/coronavirus-live-updates-us-death-toll-rises-cases-testing/5053816002/ |
| Gathering ban | 2020-03-16 | 1.00 | Idaho | https://eu.usatoday.com/story/news/health/2020/03/16/coronavirus-live-updates-us-death-toll-rises-cases-testing/5053816002/ |
| Gathering ban | 2020-03-16 | 1.00 | Indiana | https://eu.usatoday.com/story/news/health/2020/03/16/coronavirus-live-updates-us-death-toll-rises-cases-testing/5053816002/ |
| Gathering ban | 2020-03-16 | 1.00 | Iowa | https://eu.usatoday.com/story/news/health/2020/03/16/coronavirus-live-updates-us-death-toll-rises-cases-testing/5053816002/ |
| Gathering ban | 2020-03-16 | 1.00 | Kansas | https://govstatus.egov.com/coronavirus |
| Gathering ban | 2020-03-16 | 1.00 | Louisiana | https://eu.usatoday.com/story/news/health/2020/03/16/coronavirus-live-updates-us-death-toll-rises-cases-testing/5053816002/ |
| Gathering ban | 2020-03-16 | 1.00 | Maine | https://eu.usatoday.com/story/news/health/2020/03/16/coronavirus-live-updates-us-death-toll-rises-cases-testing/5053816002/ |
| Gathering ban | 2020-03-16 | 1.00 | Maryland | https://eu.usatoday.com/story/news/health/2020/03/16/coronavirus-live-updates-us-death-toll-rises-cases-testing/5053816002/ |
| Gathering ban | 2020-03-16 | 1.00 | Massachusetts | https://eu.usatoday.com/story/news/health/2020/03/16/coronavirus-live-updates-us-death-toll-rises-cases-testing/5053816002/ |
| Gathering ban | 2020-03-16 | 1.00 | Michigan | https://www.michigan.gov/whitmer/0,9309,7-387-90499_90705-521890--,00.html |
| Gathering ban | 2020-03-16 | 1.00 | Minnesota | https://eu.usatoday.com/story/news/health/2020/03/16/coronavirus-live-updates-us-death-toll-rises-cases-testing/5053816002/ |

| | | | | |
|---|---|---|---|---|
| Gathering ban | 2020-03-16 | 1.00 | Mississippi | https://eu.usatoday.com/story/news/health/2020/03/16/coronavirus-live-updates-us-death-toll-rises-cases-testing/5053816002/ |
| Gathering ban | 2020-03-16 | 1.00 | Illinois | https://eu.usatoday.com/story/news/health/2020/03/16/coronavirus-live-updates-us-death-toll-rises-cases-testing/5053816002/ |
| Gathering ban | 2020-03-16 | 1.00 | Wyoming | https://eu.usatoday.com/story/news/health/2020/03/16/coronavirus-live-updates-us-death-toll-rises-cases-testing/5053816002/ |

| **NPI** | **Date** | **Cumulative share** | **Region** | **Source** |
|---|---|---|---|---|
| School closure | 2020-03-12 | 0.01 | New Mexico | https://www.governor.state.nm.us/2020/03/12/new-mexico-schools-to-temporarily-close/ |
| School closure | 2020-03-13 | 0.05 | West Virginia | https://governor.wv.gov/News/press-releases/2020/Pages/COVID19-UPDATE-Gov.-Justice-announces-closure-of-West-Virginia-schools.aspx |
| School closure | 2020-03-13 | 0.05 | Washington | https://www.governor.wa.gov/news-media/inslee-announces-statewide-school-closures-expansion-limits-large-gatherings |
| School closure | 2020-03-13 | 0.05 | Wisconsin | https://content.govdelivery.com/accounts/WIGOV/bulletins/281127d |
| School closure | 2020-03-15 | 0.06 | Montana | http://opi.mt.gov/COVID-19-Information |
| School closure | 2020-03-16 | 0.50 | Nevada | http://www.doe.nv.gov |
| School closure | 2020-03-16 | 0.50 | New Hampshire | https://www.governor.nh.gov/news-media/press-2020/20200315-emergency-order-1.html |
| School closure | 2020-03-16 | 0.50 | New York | https://www.governor.ny.gov/news/governor-cuomo-signs-executive-order-closing-schools-statewide-two-weeks |
| School closure | 2020-03-16 | 0.50 | North Carolina | https://www.dpi.nc.gov/news/press-releases/2020/03/16/state-board-issues-guidance-personnel-facility-matters-covid-19-closure |

| | | | | |
|---|---|---|---|---|
| School closure | 2020-03-16 | 0.50 | North Dakota | https://www.nd.gov/dpi/school-closure-frequently-asked-questions |
| School closure | 2020-03-16 | 0.50 | Pennsylvania | https://www.governor.pa.gov/newsroom/governor-wolf-announces-closure-of-pennsylvania-schools/ |
| School closure | 2020-03-16 | 0.50 | Puerto Rico | https://twitter.com/wandavazquezg?ref_src=twsrc%5Etfw%7Ctwcamp%5Etweetembed%7Ctwterm%5E1238675549372977153%7Ctwgr%5E&ref_url=https%3A%2F%2Fwww.nbcnews.com%2Fhealth%2Fhealth-news%2Flive-blog%2F2020-03-14-coronavirus-news-n1158821%2Fncrd1158961 |
| School closure | 2020-03-16 | 0.50 | South Carolina | https://thehill.com/homenews/state-watch/487708-south-carolina-closes-schools-amid-outbreak |
| School closure | 2020-03-16 | 0.50 | South Dakota | https://www.nd.gov/dpi/executive-orders-education |
| School closure | 2020-03-16 | 0.50 | Tennessee | https://www.tn.gov/governor/news/2020/3/16/governor-lee-issues-statement-regarding-statewide-school-closure.html |
| School closure | 2020-03-16 | 0.50 | Utah | https://www.schools.utah.gov/File/b27ab22a-d14f-4e12-b247-f95becea39d3 |
| School closure | 2020-03-16 | 0.50 | Virginia | https://www.governor.virginia.gov/newsroom/all-releases/2020/march/headline-854442-en.html |
| School closure | 2020-03-16 | 0.50 | Michigan | https://www.michigan.gov/whitmer/0,9309,7-387-90499_90705-521890--,00.html |
| School closure | 2020-03-16 | 0.50 | Maryland | https://fox8.com/news/list-states-that-have-closed-all-schools-due-to-coronavirus/ |
| School closure | 2020-03-16 | 0.50 | Wyoming | https://www.ktvq.com/wyoming-governor-directs-schools-to-close |
| School closure | 2020-03-16 | 0.50 | Louisiana | https://www.fox61.com/article/news/health/coronavirus/connecticut-coronavirus-updates-march-12/520-ddfcc84c-1521-4354-907b-d53e2642bee9 |

| | | | | |
|---|---|---|---|---|
| School closure | 2020-03-16 | 0.50 | Alaska | https://www.aljazeera.com/news/2020/03/emergencies-closures-states-handling-coronavirus-200317213356419.html |
| School closure | 2020-03-16 | 0.50 | Arizona | https://www.azed.gov/finance/2020/03/16/school-closures-from-march-16-2020-through-march-27-2020/ |
| School closure | 2020-03-16 | 0.50 | Kentucky | https://governor.ky.gov/covid19 |
| School closure | 2020-03-16 | 0.50 | Delaware | https://www.abc27.com/news/list-states-that-have-closed-all-schools-due-to-coronavirus/ |
| School closure | 2020-03-16 | 0.50 | Illinois | https://www.chicagotribune.com/coronavirus/ct-cb-coronavirus-illinois-schools-closed-cps-parents-need-to-know-20200317-zrcim5cpcfcgnerkboyx7esg5y-story.html |
| School closure | 2020-03-16 | 0.50 | District of Columbia | https://dcps.dc.gov/coronavirus |
| School closure | 2020-03-16 | 0.50 | Hawaii | https://www.aljazeera.com/news/2020/03/emergencies-closures-states-handling-coronavirus-200317213356419.html |
| School closure | 2020-03-16 | 0.50 | Florida | http://www.fldoe.org/newsroom/latest-news/florida-department-of-education-announces-additional-guidance-for-the-2019-20-school-year.stml |
| School closure | 2020-03-16 | 0.50 | Maine | https://www.pressherald.com/2020/03/14/scarborough-closes-schools-through-at-least-march-20/ |
| School closure | 2020-03-17 | 0.74 | Massachusetts | https://fox8.com/news/list-states-that-have-closed-all-schools-due-to-coronavirus/ |
| School closure | 2020-03-17 | 0.74 | Arkansas | https://www.abc27.com/news/list-states-that-have-closed-all-schools-due-to-coronavirus/ |

| | | | | |
|---|---|---|---|---|
| School closure | 2020-03-17 | 0.74 | California | https://www.abc27.com/news/list-states-that-have-closed-all-schools-due-to-coronavirus/ |
| School closure | 2020-03-17 | 0.74 | Connecticut | https://patch.com/connecticut/guilford/coronavirus-ct-gov-closes-all-schools-state-has-now-26-cases |
| School closure | 2020-03-17 | 0.74 | Rhode Island | https://www.ride.ri.gov/InsideRIDE/AdditionalInformation/Covid19.aspx |
| School closure | 2020-03-17 | 0.74 | Oregon | https://www.oregon.gov/newsroom/Pages/NewsDetail.aspx?newsid=36203 |
| School closure | 2020-03-17 | 0.74 | Oklahoma | https://sde.ok.gov/newsblog/2020-03-16/emergency-state-board-meeting-expected-close-schools-until-april-6 |
| School closure | 2020-03-17 | 0.74 | Missouri | https://www.abc27.com/news/list-states-that-have-closed-all-schools-due-to-coronavirus/ |
| School closure | 2020-03-17 | 0.74 | Ohio | https://coronavirus.ohio.gov/wps/wcm/connect/gov/aeadbec1-574d-4a42-9ca4-4487b7a67a4f/Director%27s+Order+-+K-12+Schools+03.14.20.pdf?MOD=AJPERES&CONVERT_TO=url&CACHEID=ROOTWORKSPACE.Z18_M1HGGIK0N0JO00QO9DDDDM3000-aeadbec1-574d-4a42-9ca4-4487b7a67a4f-n5827Z4 |
| School closure | 2020-03-18 | 0.86 | Georgia | https://www.gpb.org/blogs/education-matters/2020/03/16/gov-kemp-orders-all-k-12-georgia-schools-close-until-end-of-march |
| School closure | 2020-03-18 | 0.86 | Idaho | https://www.idahostatesman.com/news/coronavirus/article241291676.html |
| School closure | 2020-03-18 | 0.86 | New Jersey | https://www.nj.gov/governor/news/news/562020/approved/20200316c.shtml |
| School closure | 2020-03-18 | 0.86 | Kansas | https://www.washingtonpost.com/education/2020/03/17/kansas-is-first-state-close-schools-rest-school-year-amid-coronavirus-crisis-california-could-be-next/ |

| | | | | |
|---|---|---|---|---|
| School closure | 2020-03-18 | 0.86 | Nebraska | https://www.education.ne.gov/publichealth/known-school-closures/ |
| School closure | 2020-03-18 | 0.86 | Vermont | https://governor.vermont.gov/press-release/gov-scott-orders-orderly-closure-vermont-prek-12-schools-week |
| School closure | 2020-03-18 | 0.86 | Minnesota | https://mn.gov/governor/assets/EO%2020-02%20Final_tcm1055-423084.pdf |
| School closure | 2020-03-18 | 0.86 | Alabama | https://www.aljazeera.com/news/2020/03/emergencies-closures-states-handling-coronavirus-200317213356419.html |
| School closure | 2020-03-19 | 0.97 | Indiana | https://www.abc27.com/news/list-states-that-have-closed-all-schools-due-to-coronavirus/ |
| School closure | 2020-03-19 | 0.97 | Texas | https://www.dallasnews.com/news/public-health/2020/03/19/gov-abbott-announces-temporary-statewide-school-restaurant-gym-closures/ |
| School closure | 2020-03-19 | 0.97 | Mississippi | https://www.wlox.com/2020/03/19/gov-tate-reeves-give-update-plans-mississippi-schools/ |
| School closure | 2020-03-23 | 0.99 | Colorado | https://www.denverpost.com/2020/03/11/colorado-schools-closed-coronavirus/ |

| **NPI** | **Date** | **Cumulative share** | **Region** | **Source** |
|---|---|---|---|---|
| Venue closure | 2020-03-15 | 0.09 | Pennsylvania | https://www.governor.pa.gov/newsroom/wolf-administration-orders-restaurants-and-bars-to-close-dine-in-service-in-mitigation-counties-to-stop-spread-of-covid-19/ |
| Venue closure | 2020-03-15 | 0.09 | Puerto Rico | https://www.estado.pr.gov/es/ordenes-ejecutivas/ |
| Venue closure | 2020-03-15 | 0.09 | District of Columbia | https://www.washingtonian.com/2020/03/16/mayor-closes-dc-bars-and-restaurants-for-dine-in-service/ |

| | | | | |
|---|---|---|---|---|
| Venue closure | 2020-03-15 | 0.09 | Ohio | https://governor.ohio.gov/wps/portal/gov/governor/media/news-and-media/dewine-orders-ohio-bars-restaurants-to-close |
| Venue closure | 2020-03-16 | 0.48 | Louisiana | https://eu.thenewsstar.com/story/news/2020/03/16/louisiana-coronavirus-cases-rise-114-legislature-resume-work/5057909002/ |
| Venue closure | 2020-03-16 | 0.48 | New York | https://www.politico.com/states/new-york/albany/story/2020/03/16/new-york-new-jersey-connecticut-closing-bars-restaurants-indefinitely-starting-monday-night-1267159 |
| Venue closure | 2020-03-16 | 0.48 | New Jersey | https://www.nj.gov/governor/news/news/562020/approved/20200316a.shtml |
| Venue closure | 2020-03-16 | 0.48 | New Hampshire | https://www.governor.nh.gov/news-media/press-2020/20200316-covid-10-businesses.htm |
| Venue closure | 2020-03-16 | 0.48 | Michigan | https://www.usnews.com/news/best-states/michigan/articles/2020-03-16/michigan-governor-closes-restaurants-to-dine-in-customers |
| Venue closure | 2020-03-16 | 0.48 | Maryland | https://governor.maryland.gov/2020/03/19/governor-hogan-announces-further-actions-to-slow-the-spread-of-covid-19-relaunches-maryland-unites-initiative/ |
| Venue closure | 2020-03-16 | 0.48 | Kentucky | https://governor.ky.gov/covid19 |
| Venue closure | 2020-03-16 | 0.48 | Indiana | https://www.wndu.com/content/news/Indiana-governor-closes-restaurants-bars-to-dine-in-customers-568830011.html |
| Venue closure | 2020-03-16 | 0.48 | Illinois | https://time.com/5803539/united-states-coronavirus-bars-restaurants/ |
| Venue closure | 2020-03-16 | 0.48 | Oregon | https://www.oregon.gov/newsroom/Pages/NewsDetail.aspx?newsid=36192 |
| Venue closure | 2020-03-16 | 0.48 | Rhode Island | https://www.ri.gov/press/view/37924 |

| | | | | |
|---|---|---|---|---|
| Venue closure | 2020-03-16 | 0.48 | Washington | https://www.governor.wa.gov/news-media/inslee-announces-statewide-shutdown-restaurants-bars-and-expanded-social-gathering-limits |
| Venue closure | 2020-03-16 | 0.48 | California | https://www.latimes.com/business/story/2020-03-15/coronavirus-close-los-angeles-restaurants |
| Venue closure | 2020-03-16 | 0.48 | Delaware | https://coronavirus.delaware.gov/wp-content/uploads/sites/177/2020/03/coronavirus_govdec_rest_bars-1.pdf |
| Venue closure | 2020-03-16 | 0.48 | Connecticut | https://www.courant.com/news/connecticut/hc-news-coronavirus-update-0316-20200316-ukhf6fmwh5cvtaob3yiavyzm5q-story.html |
| Venue closure | 2020-03-17 | 0.69 | Wisconsin | https://madison.com/ct/news/local/govt-and-politics/gov-tony-evers-orders-bars-restaurants-to-be-closed-across-wisconsin-for-in-house-dining/article_40abef70-b51e-5756-962f-801f0387eed6.html |
| Venue closure | 2020-03-17 | 0.69 | North Carolina | https://www.newsobserver.com/news/coronavirus/article241245211.html |
| Venue closure | 2020-03-17 | 0.69 | Nevada | https://nvhealthresponse.nv.gov/wp-content/uploads/2020/03/NV-Health-Reponse-COVID19-Risk-Management-Initiative.pdf |
| Venue closure | 2020-03-17 | 0.69 | Colorado | https://www.eater.com/2020/3/15/21180761/coronavirus-restaurants-bars-closed-new-york-la-chicago |
| Venue closure | 2020-03-17 | 0.69 | Vermont | https://governor.vermont.gov/press-release/governor-phil-scott-announces-new-guidance-covid-19-community-mitigation-measures |
| Venue closure | 2020-03-17 | 0.69 | Massachusetts | https://www.mass.gov/news/baker-polito-administration-announces-emergency-actions-to-address-covid-19 |
| Venue closure | 2020-03-17 | 0.69 | Minnesota | https://www.eater.com/2020/3/15/21180761/coronavirus-restaurants-bars-closed-new-york-la-chicago |
| Venue closure | 2020-03-17 | 0.69 | South Carolina | https://governor.sc.gov/sites/default/files/Documents/Executive-Orders/ |

| | | | | |
|---|---|---|---|---|
| Venue closure | 2020-03-17 | 0.69 | Florida | https://www.eater.com/2020/3/15/21180761/coronavirus-restaurants-bars-closed-new-york-la-chicago |
| Venue closure | 2020-03-17 | 0.69 | Iowa | https://governor.iowa.gov/press-release/gov-reynolds-issues-a-state-of-public-health-disaster-emergency |
| Venue closure | 2020-03-18 | 0.71 | Maine | https://www.maine.gov/governor/mills/news/governor-mills-takes-further-steps-respond-covid-19-protect-health-and-safety-maine-people |
| Venue closure | 2020-03-18 | 0.71 | Utah | https://governor.utah.gov/2020/03/18/state-orders-restaurants-bars-to-suspend-dine-in-services-to-slow-spread-of-covid-19/ |
| Venue closure | 2020-03-18 | 0.71 | West Virginia | https://governor.wv.gov/News/press-releases/2020/Pages/COVID-19-UPDATE-Executive-Order-limiting-restaurants-and-bars,-closing-casinos-statewide.aspx |
| Venue closure | 2020-03-19 | 0.83 | Nebraska | https://www.3newsnow.com/news/coronavirus/directed-health-measures-released-by-governor-ricketts-office |
| Venue closure | 2020-03-19 | 0.83 | New Mexico | https://www.governor.state.nm.us/2020/03/18/new-mexico-to-order-additional-closures-to-limit-spread-of-covid-19/ |
| Venue closure | 2020-03-19 | 0.83 | Arizona | https://azgovernor.gov/governor/news/2020/03/governor-ducey-announces-latest-covid-19-actions |
| Venue closure | 2020-03-19 | 0.83 | Texas | https://www.dallasnews.com/news/public-health/2020/03/19/gov-abbott-announces-temporary-statewide-school-restaurant-gym-closures/ |
| Venue closure | 2020-03-19 | 0.83 | Hawaii | https://www.hawaiinewsnow.com/2020/03/19/city-orders-restaurants-bars-night-clubs-close-dine-in-services-days/ |
| Venue closure | 2020-03-20 | 0.85 | Wyoming | https://health.wyo.gov/governor-and-state-health-officer-issue-public-spaces-closure-order/ |
| Venue closure | 2020-03-20 | 0.85 | North Dakota | https://www.governor.nd.gov/news/burgum-orders-bars-restaurants-closed-site-patrons-provides-additional-guidance-k-12-schools |

| | | | | |
|---|---|---|---|---|
| Venue closure | 2020-03-20 | 0.85 | Montana | https://news.mt.gov/governor-bullock-announces-closure-of-dine-in-food-service-and-alcoholic-beverage-businesses-and-other-activities-that-pose-enhanced-risks-to-curtail-spread-of-covid-19 |
| Venue closure | 2020-03-20 | 0.85 | Arkansas | https://www.nwaonline.com/news/2020/mar/20/governor-orders-gyms-restaurants-bars-c/ |
| Venue closure | 2020-03-20 | 0.85 | South Dakota | https://www.governor.nd.gov/news/burgum-orders-bars-restaurants-closed-site-patrons-provides-additional-guidance-k-12-schools |
| Venue closure | 2020-03-23 | 0.89 | Tennessee | https://www.tn.gov/governor/news/2020/3/22/gov--bill-lee-signs-executive-order-mandating-alternative-business-models-for-restaurants-and-gyms--lifts-alcohol-regulations.html |
| Venue closure | 2020-03-23 | 0.89 | Missouri | https://governor.mo.gov/press-releases/archive/governor-parson-signs-executive-order-20-05-allowing-sale-unprepared-foods |
| Venue closure | 2020-03-24 | 0.96 | Mississippi | https://www.jacksonfreepress.com/documents/2020/mar/24/mississippis-covid-19-response/ |
| Venue closure | 2020-03-24 | 0.96 | Virginia | https://www.governor.virginia.gov/newsroom/all-releases/2020/march/headline-855292-en.html |
| Venue closure | 2020-03-24 | 0.96 | Georgia | https://www.usnews.com/news/best-states/georgia/articles/2020-03-23/counties-in-georgia-enact-restrictions-as-virus-spreads |
| Venue closure | 2020-03-24 | 0.96 | Alaska | https://gov.alaska.gov/wp-content/uploads/sites/2/03232020-SOA-COVID-19-Health-Mandate-009.pdf |
| Venue closure | 2020-03-25 | 0.98 | Idaho | https://coronavirus.idaho.gov/essential-services/ |
| Venue closure | 2020-03-25 | 0.98 | Oklahoma | https://kfor.com/news/local/gov-stitt-orders-all-non-essential-businesses-to-close-in-counties-affected-by-covid-19/ |
| Venue closure | 2020-03-27 | 0.99 | Alabama | https://governor.alabama.gov/assets/2020/03/Amended-Statewide-Social-Distancing-SHO-Order-3.27.2020-FINAL.pdf |

| | | | | |
|---|---|---|---|---|
| Venue closure | 2020-03-30 | 1.00 | Kansas | https://governor.kansas.gov/governor-kelly-issues-temporary-statewide-stay-home-order-in-ongoing-effort-to-combat-covid-19/ |

| **NPI** | **Date** | **Cumulative share** | **Region** | **Source** |
|---|---|---|---|---|
| Lockdown | 2020-03-15 | 0.01 | Puerto Rico | https://www.estado.pr.gov/es/ordenes-ejecutivas/ |
| Lockdown | 2020-03-19 | 0.13 | California | https://www.gov.ca.gov/2020/03/19/governor-gavin-newsom-issues-stay-at-home-order/ |
| Lockdown | 2020-03-21 | 0.19 | New Jersey | https://www.nj.gov/governor/news/news/562020/approved/20200320j.shtml |
| Lockdown | 2020-03-21 | 0.19 | Illinois | https://www2.illinois.gov/Pages/news-item.aspx?ReleaseID=21288 |
| Lockdown | 2020-03-22 | 0.25 | New York | https://patch.com/new-york/new-york-city/new-yorks-stay-home-order-goes-effect |
| Lockdown | 2020-03-23 | 0.35 | Oregon | https://www.oregon.gov/newsroom/Pages/NewsDetail.aspx?newsid=36240 |
| Lockdown | 2020-03-23 | 0.35 | Ohio | https://coronavirus.ohio.gov/static/DirectorsOrderStayAtHome.pdf |
| Lockdown | 2020-03-23 | 0.35 | Connecticut | https://portal.ct.gov/Office-of-the-Governor/News/Press-Releases/2020/03-2020/Governor-Lamont-Releases-Guidance-to-Businesses-on-Order-Asking-Connecticut-to-Stay-Safe-Stay-Home |
| Lockdown | 2020-03-23 | 0.35 | New Mexico | https://www.governor.state.nm.us/2020/03/23/state-enacts-further-restrictions-to-stop-spread-including-stay-at-home-instruction/ |
| Lockdown | 2020-03-23 | 0.35 | Washington | https://www.governor.wa.gov/news-media/inslee-announces-stay-home-stay-healthy%C2%A0order |
| Lockdown | 2020-03-23 | 0.35 | Louisiana | https://gov.louisiana.gov/assets/Proclamations/2020/JBE-33-2020.pdf |
| Lockdown | 2020-03-24 | 0.42 | Michigan | https://www.michigan.gov/coronavirus/0,9753,7-406-98178_98455-521682--,00.html |
| Lockdown | 2020-03-24 | 0.42 | West Virginia | https://governor.wv.gov/News/press-releases/2020/Pages/COVID-19-UPDATE-Gov.-Justice-issues-Stay-at-Home-order-for-all-West-Virginians.aspx |

| | | | | |
|---|---|---|---|---|
| Lockdown | 2020-03-24 | 0.42 | Massachusetts | https://www.mass.gov/news/dph-public-health-advisory-stay-at-home-advisory |
| Lockdown | 2020-03-24 | 0.42 | Vermont | https://governor.vermont.gov/sites/scott/files/documents/ADDENDUM%206%20TO%20EXECUTIVE%20ORDER%2001-20.pdf |
| Lockdown | 2020-03-24 | 0.42 | Delaware | https://governor.delaware.gov/wp-content/uploads/sites/24/2020/03/Fifth-Modification-to-State-of-Emergency-03222020.pdf |
| Lockdown | 2020-03-25 | 0.46 | Indiana | https://www.in.gov/gov/3232.htm |
| Lockdown | 2020-03-25 | 0.46 | Hawaii | https://governor.hawaii.gov/newsroom/latest-news/office-of-the-governor-news-release-governor-ige-issues-statewide-order-to-stay-at-home-work-from-home-to-fight-covid-19/ |
| Lockdown | 2020-03-25 | 0.46 | Wisconsin | https://content.govdelivery.com/accounts/WIGOV/bulletins/282deef |
| Lockdown | 2020-03-25 | 0.46 | Idaho | https://www.idahostatesman.com/news/coronavirus/article241479406.html |
| Lockdown | 2020-03-26 | 0.48 | New Hampshire | https://www.governor.nh.gov/news-media/emergency-orders/documents/emergency-order-17-1.pdf |
| Lockdown | 2020-03-26 | 0.48 | Colorado | https://bloximages.newyork1.vip.townnews.com/coloradopolitics.com/content/tncms/assets/v3/editorial/1/5c/15c0d646-6efd-11ea-9f44-936924cd21a7/5e7bfea00dc56.pdf.pdf |
| Lockdown | 2020-03-27 | 0.51 | Utah | https://coronavirus.utah.gov/full-text-governors-stay-home-stay-safe-directive/ |
| Lockdown | 2020-03-27 | 0.51 | Minnesota | https://mn.gov/governor/assets/3a.%20EO%2020-20%20FINAL%20SIGNED%20Filed_tcm1055-425020.pdf |
| Lockdown | 2020-03-28 | 0.52 | Rhode Island | http://www.governor.ri.gov/documents/orders/Executive-Order-20-13.pdf |
| Lockdown | 2020-03-28 | 0.52 | Montana | https://news.mt.gov/governor-bullock-issues-stay-at-home-directive-to-slow-the-spread-of-covid-19 |
| Lockdown | 2020-03-28 | 0.52 | Alaska | https://gov.alaska.gov/home/covid19-healthmandates/ |

| | | | | |
|---|---|---|---|---|
| Lockdown | 2020-03-30 | 0.61 | Virginia | https://www.governor.virginia.gov/newsroom/all-releases/2020/march/headline-855702-en.html |
| Lockdown | 2020-03-30 | 0.61 | North Carolina | https://files.nc.gov/governor/documents/files/EO121-Stay-at-Home-Order-3.pdf |
| Lockdown | 2020-03-30 | 0.61 | Kansas | https://governor.kansas.gov/wp-content/uploads/2020/03/EO20-16.pdf |
| Lockdown | 2020-03-30 | 0.61 | Maryland | https://www.youtube.com/watch?v=8TPx6pxBCyM |
| Lockdown | 2020-03-30 | 0.61 | District of Columbia | https://coronavirus.dc.gov/release/mayor-bowser-issues-stay-home-order |
| Lockdown | 2020-03-31 | 0.63 | Arizona | https://azgovernor.gov/governor/news/2020/03/stay-home-stay-healthy-stay-connected |
| Lockdown | 2020-04-01 | 0.76 | Florida | https://eu.floridatoday.com/story/news/2020/04/01/coronavirus-florida-stay-home-order-what-means-explanation-what-essential-non-essential-desantis/5104936002/ |
| Lockdown | 2020-04-01 | 0.76 | Pennsylvania | https://www.nytimes.com/interactive/2020/us/coronavirus-stay-at-home-order.html |
| Lockdown | 2020-04-01 | 0.76 | Nevada | https://www.fox5vegas.com/coronavirus/nevada-gov-sisolak-issues-stay-at-home-directive-through-april-30/article_6b11e83c-7430-11ea-abe6-0f026faccbed.html |
| Lockdown | 2020-04-01 | 0.76 | Tennessee | https://publications.tnsosfiles.com/pub/execorders/exec-orders-lee22.pdf |
| Lockdown | 2020-04-02 | 0.89 | Georgia | https://gov.georgia.gov/document/2020-executive-order/04022001/download |
| Lockdown | 2020-04-02 | 0.89 | Texas | https://www.nbcnews.com/health/health-news/here-are-stay-home-orders-across-country-n1168736 |
| Lockdown | 2020-04-02 | 0.89 | Maine | https://www.maine.gov/governor/mills/news/governor-mills-issues-stay-healthy-home-mandate-2020-03-31 |
| Lockdown | 2020-04-03 | 0.89 | Mississippi | https://thehill.com/homenews/state-watch/490674-mississippi-governor-issues-stay-at-home-order |

| | | | | |
|---|---|---|---|---|
| Lockdown | 2020-04-04 | 0.91 | Alabama | https://governor.alabama.gov/newsroom/2020/04/governor-ivey-issues-stay-at-home-order/ |
| Lockdown | 2020-04-06 | 0.93 | Missouri | https://governor.mo.gov/priorities/stay-home-order |
| Lockdown | 2020-04-07 | 0.94 | South Carolina | http://abcnews4.com/news/local/gov-mcmaster-orders-stay-at-home-order-for-south-carolina |

| **NPI** | **Date** | **Cumulative share** | **Region** | **Source** |
|---|---|---|---|---|
| Work ban | 2020-03-15 | 0.01 | Puerto Rico | https://www.estado.pr.gov/es/ordenes-ejecutivas/ |
| Work ban | 2020-03-19 | 0.17 | California | https://www.gov.ca.gov/2020/03/19/governor-gavin-newsom-issues-stay-at-home-order/ |
| Work ban | 2020-03-19 | 0.17 | Pennsylvania | https://www.governor.pa.gov/newsroom/all-non-life-sustaining-businesses-in-pennsylvania-to-close-physical-locations-as-of-8-pm-today-to-slow-spread-of-covid-19/ |
| Work ban | 2020-03-21 | 0.23 | Illinois | https://www.nytimes.com/2020/03/21/world/coronavirus-news.html |
| Work ban | 2020-03-21 | 0.23 | New Jersey | https://www.nj.gov/governor/news/news/562020/approved/20200320j.shtml |
| Work ban | 2020-03-22 | 0.29 | New York | https://www.governor.ny.gov/news/no-2028-continuing-temporary-suspension-and-modification-laws-relating-disaster-emergency |
| Work ban | 2020-03-23 | 0.42 | New Mexico | https://www.governor.state.nm.us/2020/03/23/state-enacts-further-restrictions-to-stop-spread-including-stay-at-home-instruction/ |
| Work ban | 2020-03-23 | 0.42 | Washington | https://www.governor.wa.gov/news-media/inslee-announces-stay-home-stay-healthy%C2%A0order |
| Work ban | 2020-03-23 | 0.42 | Oregon | https://govstatus.egov.com/or-covid-19 |
| Work ban | 2020-03-23 | 0.42 | Connecticut | https://portal.ct.gov/Office-of-the-Governor/News/Press-Releases/2020/03-2020/Governor-Lamont-Signs-Executive-Order-Asking-Connecticut-Businesses-and-Residents-Stay-Safe |

| | | | | |
|---|---|---|---|---|
| Work ban | 2020-03-23 | 0.42 | Massachusetts | https://www.mass.gov/news/governor-charlie-baker-orders-all-non-essential-businesses-to-cease-in-person-operation |
| Work ban | 2020-03-23 | 0.42 | Ohio | https://coronavirus.ohio.gov/static/DirectorsOrderStayAtHome.pdf |
| Work ban | 2020-03-23 | 0.42 | Maryland | https://governor.maryland.gov/2020/03/23/governor-hogan-announces-closure-of-all-non-essential-businesses-175-million-relief-package-for-workers-and-small-businesses-affected-by-covid-19/ |
| Work ban | 2020-03-24 | 0.46 | Michigan | https://www.michigan.gov/coronavirus/0,9753,7-406-98178_98455-521682--,00.html |
| Work ban | 2020-03-24 | 0.46 | West Virginia | https://governor.wv.gov/News/press-releases/2020/Pages/COVID-19-UPDATE-Gov.-Justice-issues-Stay-at-Home-order-for-all-West-Virginians.aspx |
| Work ban | 2020-03-24 | 0.46 | Delaware | https://governor.delaware.gov/wp-content/uploads/sites/24/2020/03/Fourth-Modification-to-State-of-Emergency-03222020.pdf |
| Work ban | 2020-03-24 | 0.46 | Vermont | https://governor.vermont.gov/sites/scott/files/documents/ADDENDUM%206%20TO%20EXECUTIVE%20ORDER%2001-20.pdf |
| Work ban | 2020-03-25 | 0.52 | Wisconsin | https://content.govdelivery.com/accounts/WIGOV/bulletins/282deef |
| Work ban | 2020-03-25 | 0.52 | Oklahoma | https://www.sos.ok.gov/documents/executive/1919.pdf |
| Work ban | 2020-03-25 | 0.52 | Indiana | https://www.in.gov/gov/3232.htm |
| Work ban | 2020-03-25 | 0.52 | Idaho | https://www.idahostatesman.com/news/coronavirus/article241479406.html |
| Work ban | 2020-03-25 | 0.52 | Hawaii | https://governor.hawaii.gov/newsroom/latest-news/office-of-the-governor-news-release-governor-ige-issues-statewide-order-to-stay-at-home-work-from-home-to-fight-covid-19/ |
| Work ban | 2020-03-26 | 0.55 | Colorado | https://bloximages.newyork1.vip.townnews.com/coloradopolitics.com/content/tncms/assets/v3/editorial/1/5c/15c0d646-6efd-11ea-9f44-936924cd21a7/5e7bfea00dc56.pdf.pdf |
| Work ban | 2020-03-26 | 0.55 | New Hampshire | https://www.governor.nh.gov/news-media/emergency-orders/documents/emergency-order-17-1.pdf |

| | | | | |
|---|---|---|---|---|
| Work ban | 2020-03-26 | 0.55 | Kentucky | https://governor.ky.gov/attachments/20200325_Executive-Order_2020-257_Healthy-at-Home.pdf |
| Work ban | 2020-03-27 | 0.58 | Alabama | https://governor.alabama.gov/newsroom/2020/04/governor-ivey-issues-stay-at-home-order/ |
| Work ban | 2020-03-27 | 0.58 | Minnesota | https://mn.gov/governor/assets/3a.%20EO%2020-20%20FINAL%20SIGNED%20Filed_tcm1055-425020.pdf |
| Work ban | 2020-03-28 | 0.59 | Alaska | https://gov.alaska.gov/home/covid19-healthmandates/ |
| Work ban | 2020-03-28 | 0.59 | Montana | https://news.mt.gov/governor-bullock-issues-stay-at-home-directive-to-slow-the-spread-of-covid-19 |
| Work ban | 2020-03-30 | 0.64 | Kansas | https://governor.kansas.gov/governor-kelly-issues-temporary-statewide-stay-home-order-in-ongoing-effort-to-combat-covid-19/ |
| Work ban | 2020-03-30 | 0.64 | Rhode Island | http://www.governor.ri.gov/documents/orders/Executive-Order-20-13.pdf |
| Work ban | 2020-03-30 | 0.64 | District of Columbia | https://coronavirus.dc.gov/release/mayor-bowser-issues-stay-home-order |
| Work ban | 2020-03-30 | 0.64 | North Carolina | https://files.nc.gov/governor/documents/files/EO121-Stay-at-Home-Order-3.pdf |
| Work ban | 2020-04-01 | 0.66 | Tennessee | https://publications.tnsosfiles.com/pub/execorders/exec-orders-lee22.pdf |
| Work ban | 2020-04-02 | 0.74 | Texas | https://www.nbcnews.com/health/health-news/here-are-stay-home-orders-across-country-n1168736 |
| Work ban | 2020-04-07 | 0.76 | South Carolina | http://abcnews4.com/news/local/gov-mcmaster-orders-stay-at-home-order-for-south-carolina |

**Table 7.** Sources for policies implemented across different US States

| NPI | Date | Cumulative share | Region | Source |
|---|---|---|---|---|
| Event ban | 2020-03-09 | 0.15 | Bavaria | https://www.br.de/nachrichten/bayern/coronavirus-bayern-will-grossveranstaltungen-verbieten,RslNyZO |
| Event ban | 2020-03-10 | 0.37 | North Rhine-Westphalia | https://www1.wdr.de/nachrichten/themen/coronavirus/veranstaltungen-corona-virus-absage-nrw-100.html |
| Event ban | 2020-03-11 | 0.70 | Baden-Wuerttemberg | https://bnn.de/lokales/karlsruhe/baden-wuerttemberg-will-grosse-veranstaltungen-wegen-des-coronavirus-untersagen-lassen |
| Event ban | 2020-03-11 | 0.70 | Berlin | https://www.t-online.de/nachrichten/panorama/id_87498882/coronavirus-in-diesen-bundeslaendern-sind-grossveranstaltungen-verboten.html |
| Event ban | 2020-03-11 | 0.70 | Hamburg | https://www.t-online.de/nachrichten/panorama/id_87498882/coronavirus-in-diesen-bundeslaendern-sind-grossveranstaltungen-verboten.html |
| Event ban | 2020-03-11 | 0.70 | Lower Saxony | https://www.t-online.de/nachrichten/panorama/id_87498882/coronavirus-in-diesen-bundeslaendern-sind-grossveranstaltungen-verboten.html |
| Event ban | 2020-03-11 | 0.70 | Schleswig-Holstein | https://www.ndr.de/nachrichten/schleswig-holstein/Details-zur-Absage-von-Grossveranstaltungen-,pk214.html |
| Event ban | 2020-03-12 | 0.73 | Bremen | https://www.t-online.de/nachrichten/panorama/id_87498882/coronavirus-in-diesen-bundeslaendern-sind-grossveranstaltungen-verboten.html |
| Event ban | 2020-03-12 | 0.73 | Thuringia | https://www.mdr.de/thueringen/coronavirus-veranstaltungen-massnahmen-teilnehmer-100.html |
| Event ban | 2020-03-13 | 1.00 | Brandenburg | https://twitter.com/StM_Klose/status/1238028608469336070 |
| Event ban | 2020-03-13 | 1.00 | Hesse | https://twitter.com/StM_Klose/status/1238028608469336070 |
| Event ban | 2020-03-13 | 1.00 | Mecklenburg-Western Pomerania | https://twitter.com/StM_Klose/status/1238028608469336070 |

| | | | | |
|---|---|---|---|---|
| Event ban | 2020-03-13 | 1.00 | Rhineland-Palatinate | https://twitter.com/StM_Klose/status/1238028608469336070 |
| Event ban | 2020-03-13 | 1.00 | Saarland | https://twitter.com/StM_Klose/status/1238028608469336070 |
| Event ban | 2020-03-13 | 1.00 | Saxony | https://twitter.com/StM_Klose/status/1238028608469336070 |
| Event ban | 2020-03-13 | 1.00 | Saxony-Anhalt | https://twitter.com/StM_Klose/status/1238028608469336070 |

| **NPI** | **Date** | **Cumulative share** | **Region** | **Source** |
|---|---|---|---|---|
| Gathering ban | 2020-03-14 | 0.04 | Berlin | https://www.reuters.com/article/us-health-coronavirus-germany/berlin-joins-cologne-in-closing-bars-clubs-as-germany-toughens-coronavirus-response-idUSKBN2110LS |
| Gathering ban | 2020-03-17 | 0.20 | Bavaria | https://www.reuters.com/article/us-health-coronavirus-germany-economy/merkel-says-lets-get-through-this-as-shops-bars-and-churches-shut-idUSKBN213178 |
| Gathering ban | 2020-03-23 | 1.00 | Baden-Wuerttemberg | https://www.bundesregierung.de/breg-de/themen/coronavirus/faqs-neue-leitlinien-1733416 |
| Gathering ban | 2020-03-23 | 1.00 | Brandenburg | https://www.bundesregierung.de/breg-de/themen/coronavirus/faqs-neue-leitlinien-1733416 |
| Gathering ban | 2020-03-23 | 1.00 | Bremen | https://www.bundesregierung.de/breg-de/themen/coronavirus/faqs-neue-leitlinien-1733416 |
| Gathering ban | 2020-03-23 | 1.00 | Hamburg | https://www.bundesregierung.de/breg-de/themen/coronavirus/faqs-neue-leitlinien-1733416 |
| Gathering ban | 2020-03-23 | 1.00 | Hesse | https://www.bundesregierung.de/breg-de/themen/coronavirus/faqs-neue-leitlinien-1733416 |
| Gathering ban | 2020-03-23 | 1.00 | Lower Saxony | https://www.bundesregierung.de/breg-de/themen/coronavirus/faqs-neue-leitlinien-1733416 |

| | | | | |
|---|---|---|---|---|
| Gathering ban | 2020-03-23 | 1.00 | Mecklenburg-Western Pomerania | https://www.bundesregierung.de/breg-de/themen/coronavirus/faqs-neue-leitlinien-1733416 |
| Gathering ban | 2020-03-23 | 1.00 | North Rhine-Westphalia | https://www.bundesregierung.de/breg-de/themen/coronavirus/faqs-neue-leitlinien-1733416 |
| Gathering ban | 2020-03-23 | 1.00 | Rhineland-Palatinate | https://www.bundesregierung.de/breg-de/themen/coronavirus/faqs-neue-leitlinien-1733416 |
| Gathering ban | 2020-03-23 | 1.00 | Saarland | https://www.bundesregierung.de/breg-de/themen/coronavirus/faqs-neue-leitlinien-1733416 |
| Gathering ban | 2020-03-23 | 1.00 | Saxony | https://www.bundesregierung.de/breg-de/themen/coronavirus/faqs-neue-leitlinien-1733416 |
| Gathering ban | 2020-03-23 | 1.00 | Saxony-Anhalt | https://www.bundesregierung.de/breg-de/themen/coronavirus/faqs-neue-leitlinien-1733416 |
| Gathering ban | 2020-03-23 | 1.00 | Schleswig-Holstein | https://www.bundesregierung.de/breg-de/themen/coronavirus/faqs-neue-leitlinien-1733416 |
| Gathering ban | 2020-03-23 | 1.00 | Thuringia | https://www.bundesregierung.de/breg-de/themen/coronavirus/faqs-neue-leitlinien-1733416 |

| **NPI** | **Date** | **Cumulative share** | **Region** | **Source** |
|---|---|---|---|---|
| School closure | 2020-03-16 | 0.81 | Bavaria | https://www.spiegel.de/international/germany/the-shutdown-begins-across-germany-a-3c541d1d-1d42-4672-9fdc-af6c3247df76 |
| School closure | 2020-03-16 | 0.81 | Berlin | https://www.spiegel.de/international/germany/the-shutdown-begins-across-germany-a-3c541d1d-1d42-4672-9fdc-af6c3247df76 |
| School closure | 2020-03-16 | 0.81 | Bremen | https://www.spiegel.de/international/germany/the-shutdown-begins-across-germany-a-3c541d1d-1d42-4672-9fdc-af6c3247df76 |
| School closure | 2020-03-16 | 0.81 | Hamburg | https://www.spiegel.de/international/germany/the-shutdown-begins-across-germany-a-3c541d1d-1d42-4672-9fdc-af6c3247df76 |

| | | | | |
|---|---|---|---|---|
| School closure | 2020-03-16 | 0.81 | Hesse | https://www.spiegel.de/international/germany/the-shutdown-begins-across-germany-a-3c541d1d-1d42-4672-9fdc-af6c3247df76 |
| School closure | 2020-03-16 | 0.81 | Lower Saxony | https://www.spiegel.de/international/germany/the-shutdown-begins-across-germany-a-3c541d1d-1d42-4672-9fdc-af6c3247df76 |
| School closure | 2020-03-16 | 0.81 | Mecklenburg-Western Pomerania | https://www.spiegel.de/international/germany/the-shutdown-begins-across-germany-a-3c541d1d-1d42-4672-9fdc-af6c3247df76 |
| School closure | 2020-03-16 | 0.81 | North Rhine-Westphalia | https://www.spiegel.de/international/germany/the-shutdown-begins-across-germany-a-3c541d1d-1d42-4672-9fdc-af6c3247df76 |
| School closure | 2020-03-16 | 0.81 | Rhineland-Palatinate | https://www.spiegel.de/international/germany/the-shutdown-begins-across-germany-a-3c541d1d-1d42-4672-9fdc-af6c3247df76 |
| School closure | 2020-03-16 | 0.81 | Saarland | https://www.spiegel.de/international/germany/the-shutdown-begins-across-germany-a-3c541d1d-1d42-4672-9fdc-af6c3247df76 |
| School closure | 2020-03-16 | 0.81 | Saxony | https://www.spiegel.de/international/germany/germany-states-move-to-close-educational-and-daycare-facilities-a-e9c13296-002b-484b-88bc-e14ea295ff10 |
| School closure | 2020-03-16 | 0.81 | Saxony-Anhalt | https://www.spiegel.de/international/germany/the-shutdown-begins-across-germany-a-3c541d1d-1d42-4672-9fdc-af6c3247df76 |
| School closure | 2020-03-16 | 0.81 | Schleswig-Holstein | https://www.spiegel.de/international/germany/the-shutdown-begins-across-germany-a-3c541d1d-1d42-4672-9fdc-af6c3247df76 |
| School closure | 2020-03-17 | 0.97 | Baden-Wuerttemberg | https://www.spiegel.de/international/germany/the-shutdown-begins-across-germany-a-3c541d1d-1d42-4672-9fdc-af6c3247df76 |
| School closure | 2020-03-17 | 0.97 | Thuringia | https://www.spiegel.de/international/germany/the-shutdown-begins-across-germany-a-3c541d1d-1d42-4672-9fdc-af6c3247df76 |
| School closure | 2020-03-18 | 1.00 | Brandenburg | https://www.spiegel.de/international/germany/the-shutdown-begins-across-germany-a-3c541d1d-1d42-4672-9fdc-af6c3247df76 |

| NPI | Date | Cumulative share | Region | Source |
|---|---|---|---|---|
| Venue closure | 2020-03-14 | 0.05 | Berlin | https://www.reuters.com/article/us-health-coronavirus-germany/berlin-joins-cologne-in-closing-bars-clubs-as-germany-toughens-coronavirus-response-idUSKBN2110LS |
| Venue closure | 2020-03-14 | 0.05 | Saarland | https://www.reuters.com/article/us-health-coronavirus-germany/berlin-joins-cologne-in-closing-bars-clubs-as-germany-toughens-coronavirus-response-idUSKBN2110LS |
| Venue closure | 2020-03-16 | 1.00 | Baden-Wuerttemberg | https://www.bundesregierung.de/breg-de/themen/coronavirus/leitlinien-bund-laender-1731000 |
| Venue closure | 2020-03-16 | 1.00 | Bavaria | https://www.bundesregierung.de/breg-de/themen/coronavirus/leitlinien-bund-laender-1731000 |
| Venue closure | 2020-03-16 | 1.00 | Brandenburg | https://www.bundesregierung.de/breg-de/themen/coronavirus/leitlinien-bund-laender-1731000 |
| Venue closure | 2020-03-16 | 1.00 | Bremen | https://www.bundesregierung.de/breg-de/themen/coronavirus/leitlinien-bund-laender-1731000 |
| Venue closure | 2020-03-16 | 1.00 | Hamburg | https://www.bundesregierung.de/breg-de/themen/coronavirus/leitlinien-bund-laender-1731000 |
| Venue closure | 2020-03-16 | 1.00 | Hesse | https://www.bundesregierung.de/breg-de/themen/coronavirus/leitlinien-bund-laender-1731000 |
| Venue closure | 2020-03-16 | 1.00 | Lower Saxony | https://www.bundesregierung.de/breg-de/themen/coronavirus/leitlinien-bund-laender-1731000 |
| Venue closure | 2020-03-16 | 1.00 | Mecklenburg-Western Pomerania | https://www.bundesregierung.de/breg-de/themen/coronavirus/leitlinien-bund-laender-1731000 |
| Venue closure | 2020-03-16 | 1.00 | North Rhine-Westphalia | https://www.bundesregierung.de/breg-de/themen/coronavirus/leitlinien-bund-laender-1731000 |

| | | | | |
|---|---|---|---|---|
| Venue closure | 2020-03-16 | 1.00 | Rhineland-Palatinate | https://www.bundesregierung.de/breg-de/themen/coronavirus/leitlinien-bund-laender-1731000 |
| Venue closure | 2020-03-16 | 1.00 | Saxony | https://www.bundesregierung.de/breg-de/themen/coronavirus/leitlinien-bund-laender-1731000 |
| Venue closure | 2020-03-16 | 1.00 | Saxony-Anhalt | https://www.bundesregierung.de/breg-de/themen/coronavirus/leitlinien-bund-laender-1731000 |
| Venue closure | 2020-03-16 | 1.00 | Schleswig-Holstein | https://www.bundesregierung.de/breg-de/themen/coronavirus/leitlinien-bund-laender-1731000 |
| Venue closure | 2020-03-16 | 1.00 | Thuringia | https://www.bundesregierung.de/breg-de/themen/coronavirus/leitlinien-bund-laender-1731000 |

| **NPI** | **Date** | **Cumulative share** | **Region** | **Source** |
|---|---|---|---|---|
| Lockdown | 2020-03-21 | 0.15 | Bavaria | https://www.corona-katastrophenschutz.bayern.de/ |

| **NPI** | **Date** | **Cumulative share** | **Region** | **Source** |
|---|---|---|---|---|
| Work ban | *NPI implemented in no region* | | | |

**Table 8.** Sources for policies implemented across different German regions

| NPI | Date | Cumulative share | Region | Source |
|---|---|---|---|---|
| Event ban | 2020-03-10 | 0.15 | La Rioja | https://www.eldiario.es/sociedad/Sanidad-consejo-ministros-medidas_0_1004400105.html |
| Event ban | 2020-03-10 | 0.15 | Community of Madrid | https://www.eldiario.es/sociedad/Sanidad-consejo-ministros-medidas_0_1004400105.html |
| Event ban | 2020-03-12 | 0.31 | Catalonia | https://www.catalannews.com/society-science/item/catalonia-bans-events-of-over-1000-people-in-efforts-to-control-coronavirus |
| Event ban | 2020-03-13 | 1.00 | Andalusia | https://elpais.com/sociedad/2020-03-12/el-gobierno-extiende-a-toda-espana-la-recomendacion-de-cancelar-clases-y-celebrar-eventos-masivos-en-espacios-cerrados.html |
| Event ban | 2020-03-13 | 1.00 | Navarre | https://elpais.com/sociedad/2020-03-12/el-gobierno-extiende-a-toda-espana-la-recomendacion-de-cancelar-clases-y-celebrar-eventos-masivos-en-espacios-cerrados.html |
| Event ban | 2020-03-13 | 1.00 | Galicia | https://elpais.com/sociedad/2020-03-12/el-gobierno-extiende-a-toda-espana-la-recomendacion-de-cancelar-clases-y-celebrar-eventos-masivos-en-espacios-cerrados.html |
| Event ban | 2020-03-13 | 1.00 | Extremadura | https://elpais.com/sociedad/2020-03-12/el-gobierno-extiende-a-toda-espana-la-recomendacion-de-cancelar-clases-y-celebrar-eventos-masivos-en-espacios-cerrados.html |
| Event ban | 2020-03-13 | 1.00 | Region of Murcia | https://elpais.com/sociedad/2020-03-12/el-gobierno-extiende-a-toda-espana-la-recomendacion-de-cancelar-clases-y-celebrar-eventos-masivos-en-espacios-cerrados.html |

| | | | | |
|---|---|---|---|---|
| Event ban | 2020-03-13 | 1.00 | Castille-La Mancha | https://elpais.com/sociedad/2020-03-12/el-gobierno-extiende-a-toda-espana-la-recomendacion-de-cancelar-clases-y-celebrar-eventos-masivos-en-espacios-cerrados.html |
| Event ban | 2020-03-13 | 1.00 | Cantabria | https://elpais.com/sociedad/2020-03-12/el-gobierno-extiende-a-toda-espana-la-recomendacion-de-cancelar-clases-y-celebrar-eventos-masivos-en-espacios-cerrados.html |
| Event ban | 2020-03-13 | 1.00 | Canary Islands | https://elpais.com/sociedad/2020-03-12/el-gobierno-extiende-a-toda-espana-la-recomendacion-de-cancelar-clases-y-celebrar-eventos-masivos-en-espacios-cerrados.html |
| Event ban | 2020-03-13 | 1.00 | Basque Country | https://elpais.com/sociedad/2020-03-12/el-gobierno-extiende-a-toda-espana-la-recomendacion-de-cancelar-clases-y-celebrar-eventos-masivos-en-espacios-cerrados.html |
| Event ban | 2020-03-13 | 1.00 | Balearic Islands | https://elpais.com/sociedad/2020-03-12/el-gobierno-extiende-a-toda-espana-la-recomendacion-de-cancelar-clases-y-celebrar-eventos-masivos-en-espacios-cerrados.html |
| Event ban | 2020-03-13 | 1.00 | Asturias | https://elpais.com/sociedad/2020-03-12/el-gobierno-extiende-a-toda-espana-la-recomendacion-de-cancelar-clases-y-celebrar-eventos-masivos-en-espacios-cerrados.html |
| Event ban | 2020-03-13 | 1.00 | Aragon | https://elpais.com/sociedad/2020-03-12/el-gobierno-extiende-a-toda-espana-la-recomendacion-de-cancelar-clases-y-celebrar-eventos-masivos-en-espacios-cerrados.html |
| Event ban | 2020-03-13 | 1.00 | Castille and Leon | https://elpais.com/sociedad/2020-03-12/el-gobierno-extiende-a-toda-espana-la-recomendacion-de-cancelar-clases-y-celebrar-eventos-masivos-en-espacios-cerrados.html |

| | | | | |
|---|---|---|---|---|
| Event ban | 2020-03-13 | 1.00 | Valencian Community | https://elpais.com/sociedad/2020-03-12/el-gobierno-extiende-a-toda-espana-la-recomendacion-de-cancelar-clases-y-celebrar-eventos-masivos-en-espacios-cerrados.html |

| NPI | Date | Cumulative share | Region | Source |
|---|---|---|---|---|
| Gathering ban | 2020-03-15 | 1.00 | Andalusia | https://www.cnbc.com/2020/03/14/spain-declares-state-of-emergency-due-to-coronavirus.html |
| Gathering ban | 2020-03-15 | 1.00 | Navarre | https://www.cnbc.com/2020/03/14/spain-declares-state-of-emergency-due-to-coronavirus.html |
| Gathering ban | 2020-03-15 | 1.00 | La Rioja | https://www.cnbc.com/2020/03/14/spain-declares-state-of-emergency-due-to-coronavirus.html |
| Gathering ban | 2020-03-15 | 1.00 | Galicia | https://www.cnbc.com/2020/03/14/spain-declares-state-of-emergency-due-to-coronavirus.html |
| Gathering ban | 2020-03-15 | 1.00 | Extremadura | https://www.cnbc.com/2020/03/14/spain-declares-state-of-emergency-due-to-coronavirus.html |
| Gathering ban | 2020-03-15 | 1.00 | Community of Madrid | https://www.cnbc.com/2020/03/14/spain-declares-state-of-emergency-due-to-coronavirus.html |
| Gathering ban | 2020-03-15 | 1.00 | Catalonia | https://www.cnbc.com/2020/03/14/spain-declares-state-of-emergency-due-to-coronavirus.html |
| Gathering ban | 2020-03-15 | 1.00 | Region of Murcia | https://www.cnbc.com/2020/03/14/spain-declares-state-of-emergency-due-to-coronavirus.html |
| Gathering ban | 2020-03-15 | 1.00 | Castille-La Mancha | https://www.cnbc.com/2020/03/14/spain-declares-state-of-emergency-due-to-coronavirus.html |
| Gathering ban | 2020-03-15 | 1.00 | Cantabria | https://www.cnbc.com/2020/03/14/spain-declares-state-of-emergency-due-to-coronavirus.html |

| | | | | |
|---|---|---|---|---|
| Gathering ban | 2020-03-15 | 1.00 | Canary Islands | https://www.cnbc.com/2020/03/14/spain-declares-state-of-emergency-due-to-coronavirus.html |
| Gathering ban | 2020-03-15 | 1.00 | Basque Country | https://www.cnbc.com/2020/03/14/spain-declares-state-of-emergency-due-to-coronavirus.html |
| Gathering ban | 2020-03-15 | 1.00 | Balearic Islands | https://www.cnbc.com/2020/03/14/spain-declares-state-of-emergency-due-to-coronavirus.html |
| Gathering ban | 2020-03-15 | 1.00 | Asturias | https://www.cnbc.com/2020/03/14/spain-declares-state-of-emergency-due-to-coronavirus.html |
| Gathering ban | 2020-03-15 | 1.00 | Aragon | https://www.cnbc.com/2020/03/14/spain-declares-state-of-emergency-due-to-coronavirus.html |
| Gathering ban | 2020-03-15 | 1.00 | Castille and Leon | https://www.cnbc.com/2020/03/14/spain-declares-state-of-emergency-due-to-coronavirus.html |
| Gathering ban | 2020-03-15 | 1.00 | Valencian Community | https://www.cnbc.com/2020/03/14/spain-declares-state-of-emergency-due-to-coronavirus.html |

| **NPI** | **Date** | **Cumulative share** | **Region** | **Source** |
|---|---|---|---|---|
| School closure | 2020-03-11 | 0.15 | La Rioja | https://www.thestar.com.my/news/world/2020/03/10/spain039s-la-rioja-region-orders-schools-shutdown-as-coronavirus-spreads |
| School closure | 2020-03-11 | 0.15 | Community of Madrid | https://english.elpais.com/society/2020-03-09/madrid-basque-city-close-schools-as-coronavirus-continues-spread-in-spain.html |
| School closure | 2020-03-13 | 0.46 | Galicia | https://english.elpais.com/society/2020-03-12/basque-country-galicia-and-murcia-close-schools-in-bid-to-slow-coronavirus.html |
| School closure | 2020-03-13 | 0.46 | Basque Country | https://english.elpais.com/society/2020-03-12/basque-country-galicia-and-murcia-close-schools-in-bid-to-slow-coronavirus.html |
| School closure | 2020-03-13 | 0.46 | Canary Islands | https://english.elpais.com/society/2020-03-12/basque-country-galicia-and-murcia-close-schools-in-bid-to-slow-coronavirus.html |

| | | | | |
|---|---|---|---|---|
| School closure | 2020-03-13 | 0.46 | Catalonia | https://english.elpais.com/society/2020-03-12/basque-country-galicia-and-murcia-close-schools-in-bid-to-slow-coronavirus.html |
| School closure | 2020-03-16 | 1.00 | Andalusia | https://english.elpais.com/society/2020-03-12/basque-country-galicia-and-murcia-close-schools-in-bid-to-slow-coronavirus.html |
| School closure | 2020-03-16 | 1.00 | Navarre | https://english.elpais.com/society/2020-03-12/basque-country-galicia-and-murcia-close-schools-in-bid-to-slow-coronavirus.html |
| School closure | 2020-03-16 | 1.00 | Extremadura | https://english.elpais.com/society/2020-03-12/basque-country-galicia-and-murcia-close-schools-in-bid-to-slow-coronavirus.html |
| School closure | 2020-03-16 | 1.00 | Castille-La Mancha | https://english.elpais.com/society/2020-03-12/basque-country-galicia-and-murcia-close-schools-in-bid-to-slow-coronavirus.html |
| School closure | 2020-03-16 | 1.00 | Castille and Leon | https://english.elpais.com/society/2020-03-12/basque-country-galicia-and-murcia-close-schools-in-bid-to-slow-coronavirus.html |
| School closure | 2020-03-16 | 1.00 | Cantabria | https://english.elpais.com/society/2020-03-12/basque-country-galicia-and-murcia-close-schools-in-bid-to-slow-coronavirus.html |
| School closure | 2020-03-16 | 1.00 | Balearic Islands | https://english.elpais.com/society/2020-03-12/basque-country-galicia-and-murcia-close-schools-in-bid-to-slow-coronavirus.html |
| School closure | 2020-03-16 | 1.00 | Asturias | https://english.elpais.com/society/2020-03-12/basque-country-galicia-and-murcia-close-schools-in-bid-to-slow-coronavirus.html |
| School closure | 2020-03-16 | 1.00 | Aragon | https://english.elpais.com/society/2020-03-12/basque-country-galicia-and-murcia-close-schools-in-bid-to-slow-coronavirus.html |
| School closure | 2020-03-16 | 1.00 | Region of Murcia | https://english.elpais.com/society/2020-03-12/basque-country-galicia-and-murcia-close-schools-in-bid-to-slow-coronavirus.html |
| School closure | 2020-03-16 | 1.00 | Valencian Community | https://english.elpais.com/society/2020-03-12/basque-country-galicia-and-murcia-close-schools-in-bid-to-slow-coronavirus.html |
| **NPI** | **Date** | **Cumulative share** | **Region** | **Source** |

| | | | | |
|---|---|---|---|---|
| Venue closure | 2020-03-13 | 0.29 | Galicia | https://www.nytimes.com/aponline/2020/03/14/world/europe/ap-eu-virus-outbreak-spain.html |
| Venue closure | 2020-03-13 | 0.29 | Cantabria | https://www.eldiario.es/cantabria/ultima-hora/Cantabria-hosteleria-superficies-edificios-actividades_0_1005450439.html |
| Venue closure | 2020-03-13 | 0.29 | Castille and Leon | https://twitter.com/FranciscoIgea/status/1238571610468220929 |
| Venue closure | 2020-03-13 | 0.29 | Catalonia | https://elpais.com/espana/catalunya/2020-03-13/cataluna-cierra-pistas-de-esqui-discotecas-y-areas-comerciales-que-no-sean-de-alimentacion.html |
| Venue closure | 2020-03-14 | 0.56 | Valencian Community | https://english.elpais.com/society/2020-03-13/madrid-orders-restaurants-and-bars-to-close-from-saturday-onward-to-slow-coronavirus-spread.html |
| Venue closure | 2020-03-14 | 0.56 | Asturias | https://cadenaser.com/emisora/2020/03/13/ser_gijon/1584138177_171951.html |
| Venue closure | 2020-03-14 | 0.56 | Community of Madrid | https://english.elpais.com/society/2020-03-13/madrid-orders-restaurants-and-bars-to-close-from-saturday-onward-to-slow-coronavirus-spread.html |
| Venue closure | 2020-03-15 | 1.00 | Navarre | https://www.cnbc.com/2020/03/14/spain-declares-state-of-emergency-due-to-coronavirus.html |
| Venue closure | 2020-03-15 | 1.00 | La Rioja | https://www.cnbc.com/2020/03/14/spain-declares-state-of-emergency-due-to-coronavirus.html |
| Venue closure | 2020-03-15 | 1.00 | Extremadura | https://www.cnbc.com/2020/03/14/spain-declares-state-of-emergency-due-to-coronavirus.html |
| Venue closure | 2020-03-15 | 1.00 | Andalusia | https://www.cnbc.com/2020/03/14/spain-declares-state-of-emergency-due-to-coronavirus.html |
| Venue closure | 2020-03-15 | 1.00 | Canary Islands | https://www.cnbc.com/2020/03/14/spain-declares-state-of-emergency-due-to-coronavirus.html |

| | | | | |
|---|---|---|---|---|
| Venue closure | 2020-03-15 | 1.00 | Basque Country | https://www.cnbc.com/2020/03/14/spain-declares-state-of-emergency-due-to-coronavirus.html |
| Venue closure | 2020-03-15 | 1.00 | Balearic Islands | https://www.cnbc.com/2020/03/14/spain-declares-state-of-emergency-due-to-coronavirus.html |
| Venue closure | 2020-03-15 | 1.00 | Aragon | https://www.cnbc.com/2020/03/14/spain-declares-state-of-emergency-due-to-coronavirus.html |
| Venue closure | 2020-03-15 | 1.00 | Region of Murcia | https://www.cnbc.com/2020/03/14/spain-declares-state-of-emergency-due-to-coronavirus.html |
| Venue closure | 2020-03-15 | 1.00 | Castille-La Mancha | https://www.cnbc.com/2020/03/14/spain-declares-state-of-emergency-due-to-coronavirus.html |

| **NPI** | **Date** | **Cumulative share** | **Region** | **Source** |
|---|---|---|---|---|
| Lockdown | 2020-03-15 | 1.00 | Andalusia | https://www.cnbc.com/2020/03/14/spain-declares-state-of-emergency-due-to-coronavirus.html |
| Lockdown | 2020-03-15 | 1.00 | Navarre | https://www.cnbc.com/2020/03/14/spain-declares-state-of-emergency-due-to-coronavirus.html |
| Lockdown | 2020-03-15 | 1.00 | La Rioja | https://www.cnbc.com/2020/03/14/spain-declares-state-of-emergency-due-to-coronavirus.html |
| Lockdown | 2020-03-15 | 1.00 | Galicia | https://www.cnbc.com/2020/03/14/spain-declares-state-of-emergency-due-to-coronavirus.html |
| Lockdown | 2020-03-15 | 1.00 | Extremadura | https://www.cnbc.com/2020/03/14/spain-declares-state-of-emergency-due-to-coronavirus.html |
| Lockdown | 2020-03-15 | 1.00 | Community of Madrid | https://www.cnbc.com/2020/03/14/spain-declares-state-of-emergency-due-to-coronavirus.html |

| NPI | Date | Cumulative share | Region | Source |
|---|---|---|---|---|
| Lockdown | 2020-03-15 | 1.00 | Catalonia | https://www.cnbc.com/2020/03/14/spain-declares-state-of-emergency-due-to-coronavirus.html |
| Lockdown | 2020-03-15 | 1.00 | Region of Murcia | https://www.cnbc.com/2020/03/14/spain-declares-state-of-emergency-due-to-coronavirus.html |
| Lockdown | 2020-03-15 | 1.00 | Castille-La Mancha | https://www.cnbc.com/2020/03/14/spain-declares-state-of-emergency-due-to-coronavirus.html |
| Lockdown | 2020-03-15 | 1.00 | Cantabria | https://www.cnbc.com/2020/03/14/spain-declares-state-of-emergency-due-to-coronavirus.html |
| Lockdown | 2020-03-15 | 1.00 | Canary Islands | https://www.cnbc.com/2020/03/14/spain-declares-state-of-emergency-due-to-coronavirus.html |
| Lockdown | 2020-03-15 | 1.00 | Basque Country | https://www.cnbc.com/2020/03/14/spain-declares-state-of-emergency-due-to-coronavirus.html |
| Lockdown | 2020-03-15 | 1.00 | Balearic Islands | https://www.cnbc.com/2020/03/14/spain-declares-state-of-emergency-due-to-coronavirus.html |
| Lockdown | 2020-03-15 | 1.00 | Asturias | https://www.cnbc.com/2020/03/14/spain-declares-state-of-emergency-due-to-coronavirus.html |
| Lockdown | 2020-03-15 | 1.00 | Aragon | https://www.cnbc.com/2020/03/14/spain-declares-state-of-emergency-due-to-coronavirus.html |
| Lockdown | 2020-03-15 | 1.00 | Castille and Leon | https://www.cnbc.com/2020/03/14/spain-declares-state-of-emergency-due-to-coronavirus.html |
| Lockdown | 2020-03-15 | 1.00 | Valencian Community | https://www.cnbc.com/2020/03/14/spain-declares-state-of-emergency-due-to-coronavirus.html |

| | | | | |
|---|---|---|---|---|
| Work ban | 2020-03-30 | 1.00 | Andalusia | https://elpais.com/espana/2020-03-28/el-gobierno-amplia-el-confinamiento-los-trabajadores-de-actividades-no-esenciales-deberan-quedarse-en-casa.html |
| Work ban | 2020-03-30 | 1.00 | Navarre | https://elpais.com/espana/2020-03-28/el-gobierno-amplia-el-confinamiento-los-trabajadores-de-actividades-no-esenciales-deberan-quedarse-en-casa.html |
| Work ban | 2020-03-30 | 1.00 | La Rioja | https://elpais.com/espana/2020-03-28/el-gobierno-amplia-el-confinamiento-los-trabajadores-de-actividades-no-esenciales-deberan-quedarse-en-casa.html |
| Work ban | 2020-03-30 | 1.00 | Galicia | https://elpais.com/espana/2020-03-28/el-gobierno-amplia-el-confinamiento-los-trabajadores-de-actividades-no-esenciales-deberan-quedarse-en-casa.html |
| Work ban | 2020-03-30 | 1.00 | Extremadura | https://elpais.com/espana/2020-03-28/el-gobierno-amplia-el-confinamiento-los-trabajadores-de-actividades-no-esenciales-deberan-quedarse-en-casa.html |
| Work ban | 2020-03-30 | 1.00 | Community of Madrid | https://elpais.com/espana/2020-03-28/el-gobierno-amplia-el-confinamiento-los-trabajadores-de-actividades-no-esenciales-deberan-quedarse-en-casa.html |
| Work ban | 2020-03-30 | 1.00 | Catalonia | https://elpais.com/espana/2020-03-28/el-gobierno-amplia-el-confinamiento-los-trabajadores-de-actividades-no-esenciales-deberan-quedarse-en-casa.html |
| Work ban | 2020-03-30 | 1.00 | Region of Murcia | https://elpais.com/espana/2020-03-28/el-gobierno-amplia-el-confinamiento-los-trabajadores-de-actividades-no-esenciales-deberan-quedarse-en-casa.html |

| | | | | |
|---|---|---|---|---|
| Work ban | 2020-03-30 | 1.00 | Castille-La Mancha | https://elpais.com/espana/2020-03-28/el-gobierno-amplia-el-confinamiento-los-trabajadores-de-actividades-no-esenciales-deberan-quedarse-en-casa.html |
| Work ban | 2020-03-30 | 1.00 | Cantabria | https://elpais.com/espana/2020-03-28/el-gobierno-amplia-el-confinamiento-los-trabajadores-de-actividades-no-esenciales-deberan-quedarse-en-casa.html |
| Work ban | 2020-03-30 | 1.00 | Canary Islands | https://elpais.com/espana/2020-03-28/el-gobierno-amplia-el-confinamiento-los-trabajadores-de-actividades-no-esenciales-deberan-quedarse-en-casa.html |
| Work ban | 2020-03-30 | 1.00 | Basque Country | https://elpais.com/espana/2020-03-28/el-gobierno-amplia-el-confinamiento-los-trabajadores-de-actividades-no-esenciales-deberan-quedarse-en-casa.html |
| Work ban | 2020-03-30 | 1.00 | Balearic Islands | https://elpais.com/espana/2020-03-28/el-gobierno-amplia-el-confinamiento-los-trabajadores-de-actividades-no-esenciales-deberan-quedarse-en-casa.html |
| Work ban | 2020-03-30 | 1.00 | Asturias | https://elpais.com/espana/2020-03-28/el-gobierno-amplia-el-confinamiento-los-trabajadores-de-actividades-no-esenciales-deberan-quedarse-en-casa.html |
| Work ban | 2020-03-30 | 1.00 | Aragon | https://elpais.com/espana/2020-03-28/el-gobierno-amplia-el-confinamiento-los-trabajadores-de-actividades-no-esenciales-deberan-quedarse-en-casa.html |
| Work ban | 2020-03-30 | 1.00 | Castille and Leon | https://elpais.com/espana/2020-03-28/el-gobierno-amplia-el-confinamiento-los-trabajadores-de-actividades-no-esenciales-deberan-quedarse-en-casa.html |

| | | | | |
|---|---|---|---|---|
| Work ban | 2020-03-30 | 1.00 | Valencian Community | https://elpais.com/espana/2020-03-28/el-gobierno-amplia-el-confinamiento-los-trabajadores-de-actividades-no-esenciales-deberan-quedarse-en-casa.html |

**Table 9.** Sources for policies implemented across different Spanish regions

| NPI | Date | Cumulative share | Region | Source |
|---|---|---|---|---|
| Event ban | 2020-03-12 | 0.27 | Alberta | https://www.alberta.ca/release.cfm?xID=6980324A5B1B0-BC2C-40A8-A6AD9E30E3189425 |
| Event ban | 2020-03-12 | 0.27 | British Columbia | https://news.gov.bc.ca/releases/2020HLTH0077-000484 |
| Event ban | 2020-03-12 | 0.27 | New Brunswick | https://www2.gnb.ca/content/gnb/en/news/news_release.2020.03.0114.html |
| Event ban | 2020-03-13 | 0.71 | Manitoba | https://news.gov.mb.ca/news/index.html?item=46933&posted=2020-03-13 |
| Event ban | 2020-03-13 | 0.71 | Newfoundland and Labrador | https://www.gov.nl.ca/releases/2020/tcii/0313n04/ |
| Event ban | 2020-03-13 | 0.71 | Nunavut | https://www.gov.nu.ca/health/news/government-nunavut-response-covid-19 |
| Event ban | 2020-03-13 | 0.71 | Ontario | https://news.ontario.ca/mtc/en/2020/03/statement-from-minister-elliott-and-minister-macleod-on-the-2019-novel-coronavirus-covid-19-1.html |
| Event ban | 2020-03-14 | 0.94 | Quebec | http://www.fil-information.gouv.qc.ca/Pages/Article.aspx?lang=en&motsCles=Covid&listeThe=&listeReg=&listeDiff=&type=&dateDebut=2019-09-28&dateFin=2020-03-28&afficherResultats=oui&Page=5&idArticle=2803149905 |
| Event ban | 2020-03-15 | 0.96 | Nova Scotia | https://novascotia.ca/news/release/?id=20200315002 |
| Event ban | 2020-03-16 | 1.00 | Prince Edward Island | https://www.princeedwardisland.ca/en/news/premier-announces-initial-financial-support-declares-public-health-emergency |
| Event ban | 2020-03-16 | 1.00 | Saskatchewan | https://www.saskatchewan.ca/government/news-and-media/2020/march/13/further-measures-for-covid-19 |
| Event ban | 2020-03-16 | 1.00 | Yukon | https://yukon.ca/en/news/chief-medical-officer-health-recommends-broad-new-measures-yukon |
| Event ban | 2020-03-22 | 1.00 | Northwest Territories | https://www.hss.gov.nt.ca/en/newsroom/all-gatherings-are-advised-cancel-effective-immediately |

| NPI | Date | Cumulative share | Region | Source |
|---|---|---|---|---|
| Gathering ban | 2020-03-16 | 0.00 | Prince Edward Island | https://www.princeedwardisland.ca/en/news/premier-announces-initial-financial-support-declares-public-health-emergency |
| Gathering ban | 2020-03-19 | 0.02 | New Brunswick | https://www2.gnb.ca/content/gnb/en/news/news_release.2020.03.0139.html |
| Gathering ban | 2020-03-20 | 0.06 | Saskatchewan | https://www.saskatchewan.ca/government/news-and-media/2020/march/20/covid-19-update-march-20 |
| Gathering ban | 2020-03-21 | 0.28 | Quebec | http://www.fil-information.gouv.qc.ca/Pages/Article.aspx?lang=en&motsCles=Covid&listeThe=&listeReg=&listeDiff=&type=&dateDebut=2019-09-28&dateFin=2020-03-28&afficherResultats=oui&Page=2&idArticle=2803211636 |
| Gathering ban | 2020-03-22 | 0.31 | Nova Scotia | https://dailyhive.com/vancouver/nova-scotia-coronavirus-state-of-emergency |
| Gathering ban | 2020-03-22 | 0.31 | Yukon | https://yukon.ca/en/news/yukons-chief-medical-officer-health-provides-update-covid-19-0 |
| Gathering ban | 2020-03-24 | 0.31 | Nunavut | https://www.gov.nu.ca/health/news/chief-public-health-officer-orders-prohibition-travel-nunavut-limited-exceptions |
| Gathering ban | 2020-03-27 | 0.43 | Alberta | https://www.cbc.ca/news/canada/edmonton/alberta-covid-19-coronavirus-deena-hinshaw-1.5512445 |
| Gathering ban | 2020-03-28 | 0.81 | Ontario | https://globalnews.ca/news/6746181/ontario-ban-gatherings/ |
| Gathering ban | 2020-03-31 | 0.83 | Newfoundland and Labrador | https://www.gov.nl.ca/covid-19/faqs/ |
| Gathering ban | 2020-04-01 | 0.86 | Manitoba | https://news.gov.mb.ca/news/index.html?item=47337&posted=2020-03-30 |

| | | | | |
|---|---|---|---|---|
| Gathering ban | 2020-04-11 | 0.87 | Northwest Territories | https://www.gov.nt.ca/en/newsroom/two-new-orders-nwt-chief-public-health-officer-strengthen-response-covid-19-pandemic |

| **NPI** | **Date** | **Cumulative share** | **Region** | **Source** |
|---|---|---|---|---|
| School closure | 2020-03-13 | 0.02 | New Brunswick | https://www2.gnb.ca/content/gnb/en/news/news_release.2020.03.0117.html |
| School closure | 2020-03-14 | 0.25 | Quebec | http://www.fil-information.gouv.qc.ca/Pages/Article.aspx?lang=en&motsCles=Covid&listeThe=&listeReg=&listeDiff=&type=&dateDebut=2019-09-28&dateFin=2020-03-28&afficherResultats=oui&Page=6&idArticle=2803137507 |
| School closure | 2020-03-15 | 0.36 | Alberta | https://www.alberta.ca/release.cfm?xID=69818C355F188-C2A3-F5C6-875A2A33929D5C05 |
| School closure | 2020-03-16 | 0.80 | Newfoundland and Labrador | https://www.gov.nl.ca/releases/2020/eecd/0316n04/ |
| School closure | 2020-03-16 | 0.80 | Northwest Territories | https://cabinradio.ca/32133/news/education/nwt-tells-schools-to-close-until-after-easter-daycares-unaffected/ |
| School closure | 2020-03-16 | 0.80 | Ontario | https://news.ontario.ca/maesd/en/2020/03/statement-from-minister-elliott-and-minister-romano-on-the-2019-novel-coronavirus-covid-19.html |
| School closure | 2020-03-16 | 0.80 | Saskatchewan | https://www.saskatchewan.ca/government/news-and-media/2020/march/16/class-suspensions |
| School closure | 2020-03-17 | 0.96 | British Columbia | https://news.gov.bc.ca/releases/2020EMBC0014-000552 |
| School closure | 2020-03-17 | 0.96 | Nova Scotia | https://novascotia.ca/news/release/?id=20200315002 |
| School closure | 2020-03-17 | 0.96 | Nunavut | https://www.gov.nu.ca/health/news/temporary-nunavut-wide-school-and-daycare-closures-precaution-covid-19 |

| | | | | |
|---|---|---|---|---|
| School closure | 2020-03-17 | 0.96 | Prince Edward Island | https://www.princeedwardisland.ca/en/news/province-announces-covid-19-related-closures |
| School closure | 2020-03-18 | 0.96 | Yukon | https://yukon.ca/en/news/chief-medical-officer-health-declares-public-health-emergency |
| School closure | 2020-03-23 | 1.00 | Manitoba | https://news.gov.mb.ca/news/index.html?item=46936&posted=2020-03-14 |

| **NPI** | **Date** | **Cumulative share** | **Region** | **Source** |
|---|---|---|---|---|
| Venue closure | 2020-03-15 | 0.23 | Quebec | http://www.fil-information.gouv.qc.ca/Pages/Article.aspx?lang=en&motsCles=Covid&listeThe=&listeReg=&listeDiff=&type=&dateDebut=2019-09-28&dateFin=2020-03-28&afficherResultats=oui&Page=5&idArticle=2803159403 |
| Venue closure | 2020-03-16 | 0.61 | Ontario | https://news.ontario.ca/mohltc/en/2020/03/enhanced-measures-to-protect-ontarians-from-covid-19.html |
| Venue closure | 2020-03-17 | 0.89 | Alberta | https://www.alberta.ca/release.cfm?xID=69828242A5FFC-D75A-C83E-690D8028C0C4E09F |
| Venue closure | 2020-03-17 | 0.89 | British Columbia | https://news.gov.bc.ca/releases/2020EMBC0014-000552 |
| Venue closure | 2020-03-17 | 0.89 | New Brunswick | https://www2.gnb.ca/content/gnb/en/news/news_release.2020.03.0127.html |
| Venue closure | 2020-03-17 | 0.89 | Prince Edward Island | https://www.princeedwardisland.ca/en/news/chief-public-health-officer-urges-islanders-work-together-reduce-spread-covid-19 |
| Venue closure | 2020-03-19 | 0.92 | Nova Scotia | https://novascotia.ca/news/release/?id=20200317005 |
| Venue closure | 2020-03-20 | 0.95 | Nunavut | https://www.gov.nu.ca/health/news/minister-health-declares-public-health-emergency |

| NPI | Date | Cumulative share | Region | Source |
| --- | --- | --- | --- | --- |
| Venue closure | 2020-03-20 | 0.95 | Saskatchewan | https://www.saskatchewan.ca/government/news-and-media/2020/march/20/covid-19-update-march-20 |
| Venue closure | 2020-03-22 | 0.95 | Northwest Territories | https://www.hss.gov.nt.ca/en/newsroom/all-gatherings-are-advised-cancel-effective-immediately |
| Venue closure | 2020-03-23 | 0.96 | Newfoundland and Labrador | https://www.gov.nl.ca/covid-19/ |
| Venue closure | 2020-03-25 | 0.96 | Yukon | https://yukon.ca/en/news/yukons-chief-medical-officer-health-provides-update-covid-19-0 |
| Venue closure | 2020-04-01 | 1.00 | Manitoba | https://news.gov.mb.ca/news/index.html?item=47337&posted=2020-03-30 |

| NPI | Date | Cumulative share | Region | Source |
| --- | --- | --- | --- | --- |
| Lockdown | 2020-03-29 | 0.02 | New Brunswick | https://www2.gnb.ca/content/gnb/en/news/news_release.2020.03.0164.html |
| Lockdown | 2020-03-30 | 0.41 | Ontario | https://news.ontario.ca/mohltc/en/2020/03/statement-from-the-chief-medical-officer-of-health.html |

| NPI | Date | Cumulative share | Region | Source |
| --- | --- | --- | --- | --- |
| Work ban | 2020-03-24 | 0.39 | Ontario | https://news.ontario.ca/opo/en/2020/03/ontario-closing-at-risk-workplaces-to-protect-health-and-safety.html |
| Work ban | 2020-03-27 | 0.39 | Prince Edward Island | https://www.princeedwardisland.ca/en/news/prince-edward-island-extends-closures-for-schools-daycares-non-essential-services |
| Work ban | 2020-04-01 | 0.43 | Manitoba | https://news.gov.mb.ca/news/index.html?item=47337&posted=2020-03-30 |

**Table 10.** Sources for policies implemented across different Canadian regions

| NPI | Date | Cumulative share | Region | Source |
|---|---|---|---|---|
| Event ban | 2020-03-16 | 1.00 | Western Australia | https://www.watoday.com.au/politics/federal/effective-ban-on-non-essential-mass-gatherings-of-500-people-20200313-p549u5.html |
| Event ban | 2020-03-16 | 1.00 | Northern Territory | https://www.watoday.com.au/politics/federal/effective-ban-on-non-essential-mass-gatherings-of-500-people-20200313-p549u5.html |
| Event ban | 2020-03-16 | 1.00 | South Australia | https://www.watoday.com.au/politics/federal/effective-ban-on-non-essential-mass-gatherings-of-500-people-20200313-p549u5.html |
| Event ban | 2020-03-16 | 1.00 | Queensland | https://www.watoday.com.au/politics/federal/effective-ban-on-non-essential-mass-gatherings-of-500-people-20200313-p549u5.html |
| Event ban | 2020-03-16 | 1.00 | New South Wales | https://www.watoday.com.au/politics/federal/effective-ban-on-non-essential-mass-gatherings-of-500-people-20200313-p549u5.html |
| Event ban | 2020-03-16 | 1.00 | Victoria | https://www.watoday.com.au/politics/federal/effective-ban-on-non-essential-mass-gatherings-of-500-people-20200313-p549u5.html |
| Event ban | 2020-03-16 | 1.00 | Tasmania | https://www.watoday.com.au/politics/federal/effective-ban-on-non-essential-mass-gatherings-of-500-people-20200313-p549u5.html |
| Event ban | 2020-03-16 | 1.00 | Australian Capital Territory | https://www.watoday.com.au/politics/federal/effective-ban-on-non-essential-mass-gatherings-of-500-people-20200313-p549u5.html |
| **NPI** | **Date** | **Cumulative share** | **Region** | **Source** |
| Gathering ban | 2020-03-23 | 0.01 | Northern Territory | https://coronavirus.nt.gov.au/updates |
| Gathering ban | 2020-03-28 | 0.08 | South Australia | https://www.sa.gov.au/__data/assets/pdf_file/0003/605055/Emergency-Management-GatheringsCOVID-19-Direction-2020_FINAL.pdf |

| | | | | |
|---|---|---|---|---|
| Gathering ban | 2020-03-29 | 0.90 | Queensland | https://www.health.gov.au/news/health-alerts/novel-coronavirus-2019-ncov-health-alert/how-to-protect-yourself-and-others-from-coronavirus-covid-19/limits-on-public-gatherings-for-coronavirus-covid-19 |
| Gathering ban | 2020-03-29 | 0.90 | New South Wales | https://www.health.gov.au/news/health-alerts/novel-coronavirus-2019-ncov-health-alert/how-to-protect-yourself-and-others-from-coronavirus-covid-19/limits-on-public-gatherings-for-coronavirus-covid-19 |
| Gathering ban | 2020-03-29 | 0.90 | Victoria | https://www.health.gov.au/news/health-alerts/novel-coronavirus-2019-ncov-health-alert/how-to-protect-yourself-and-others-from-coronavirus-covid-19/limits-on-public-gatherings-for-coronavirus-covid-19 |
| Gathering ban | 2020-03-29 | 0.90 | Tasmania | https://www.health.gov.au/news/health-alerts/novel-coronavirus-2019-ncov-health-alert/how-to-protect-yourself-and-others-from-coronavirus-covid-19/limits-on-public-gatherings-for-coronavirus-covid-19 |
| Gathering ban | 2020-03-29 | 0.90 | Australian Capital Territory | https://www.health.gov.au/news/health-alerts/novel-coronavirus-2019-ncov-health-alert/how-to-protect-yourself-and-others-from-coronavirus-covid-19/limits-on-public-gatherings-for-coronavirus-covid-19 |
| Gathering ban | 2020-04-01 | 1.00 | Western Australia | https://ww2.health.wa.gov.au/~/media/Files/Corporate/general%20documents/Infectious%20diseases/PDF/Coronavirus/COVID19-Agency-Advisory-14-2020.pdf |

| **NPI** | **Date** | **Cumulative share** | **Region** | **Source** |
|---|---|---|---|---|
| School closure | 2020-03-16 | 0.32 | New South Wales | https://www.abc.net.au/news/2020-03-23/coronavirus-parents-told-to-keep-children-home-from-school/12079524 |
| School closure | 2020-03-23 | 0.60 | Victoria | https://www.theguardian.com/world/2020/mar/22/victoria-nsw-lockdowns-scott-morrison-coronavirus-national-cabinet |
| School closure | 2020-03-23 | 0.60 | Australian Capital Territory | https://www.theguardian.com/world/2020/mar/22/victoria-nsw-lockdowns-scott-morrison-coronavirus-national-cabinet |

| | | | | |
|---|---|---|---|---|
| School closure | 2020-04-20 | 0.80 | Queensland | https://www.theguardian.com/australia-news/2020/apr/13/are-schools-open-closed-term-2-australia-coronavirus-easter-holidays |

| NPI | Date | Cumulative share | Region | Source |
|---|---|---|---|---|
| Venue closure | 2020-03-23 | 1.00 | Western Australia | https://www.theguardian.com/world/live/2020/mar/22/coronavirus-updates-live-australia-nsw-victoria-qld-tasmania-cases-government-stimulus-latest-update-news |
| Venue closure | 2020-03-23 | 1.00 | Northern Territory | https://www.theguardian.com/world/live/2020/mar/22/coronavirus-updates-live-australia-nsw-victoria-qld-tasmania-cases-government-stimulus-latest-update-news |
| Venue closure | 2020-03-23 | 1.00 | South Australia | https://www.theguardian.com/world/live/2020/mar/22/coronavirus-updates-live-australia-nsw-victoria-qld-tasmania-cases-government-stimulus-latest-update-news |
| Venue closure | 2020-03-23 | 1.00 | Queensland | https://www.theguardian.com/world/live/2020/mar/22/coronavirus-updates-live-australia-nsw-victoria-qld-tasmania-cases-government-stimulus-latest-update-news |
| Venue closure | 2020-03-23 | 1.00 | New South Wales | https://www.theguardian.com/world/live/2020/mar/22/coronavirus-updates-live-australia-nsw-victoria-qld-tasmania-cases-government-stimulus-latest-update-news |
| Venue closure | 2020-03-23 | 1.00 | Victoria | https://www.theguardian.com/world/live/2020/mar/22/coronavirus-updates-live-australia-nsw-victoria-qld-tasmania-cases-government-stimulus-latest-update-news |
| Venue closure | 2020-03-23 | 1.00 | Tasmania | https://www.theguardian.com/world/live/2020/mar/22/coronavirus-updates-live-australia-nsw-victoria-qld-tasmania-cases-government-stimulus-latest-update-news |

| | | | | |
|---|---|---|---|---|
| Venue closure | 2020-03-23 | 1.00 | Australian Capital Territory | https://www.theguardian.com/world/live/2020/mar/22/coronavirus-updates-live-australia-nsw-victoria-qld-tasmania-cases-government-stimulus-latest-update-news |

| NPI | Date | Cumulative share | Region | Source |
|---|---|---|---|---|
| Lockdown | 2020-03-29 | 0.02 | Australian Capital Territory | https://www.covid19.act.gov.au/news-articles/latest-federal-government-announcement |
| Lockdown | 2020-03-31 | 0.62 | New South Wales | https://www.legislation.nsw.gov.au/_emergency/Public%20Health%20(COVID-19%20Restrictions%20on%20Gathering%20and%20Movement)%20Order%202020.pdf |
| Lockdown | 2020-03-31 | 0.62 | Victoria | https://www.vic.gov.au/coronavirusresponse |
| Lockdown | 2020-03-31 | 0.62 | Tasmania | http://www.premier.tas.gov.au/releases/keeping_tasmanians_safe_and_secure_stay_home,_save_lives |
| Lockdown | 2020-04-02 | 0.82 | Queensland | https://www.health.qld.gov.au/system-governance/legislation/cho-public-health-directions-under-expanded-public-health-act-powers/home-confinement-movement-gathering-direction |

**Table 11.** Sources for policies implemented across different Australian regions

| NPI | Date | Cumulative share | Region | Source |
|---|---|---|---|---|
| Event ban | 2020-03-02 | 0.32 | Lombardia | https://www.gazzettaufficiale.it/eli/id/2020/03/01/20A01381/sg |
| Event ban | 2020-03-02 | 0.32 | Veneto | https://www.gazzettaufficiale.it/eli/id/2020/03/01/20A01381/sg |
| Event ban | 2020-03-02 | 0.32 | Emilia-Romagna | https://www.gazzettaufficiale.it/eli/id/2020/03/01/20A01381/sg |
| Event ban | 2020-03-08 | 1.00 | Trentino-South Tyrol | https://www.gazzettaufficiale.it/showNewsDetail?id=2513&backTo=archivio&anno=2020&provenienza=archivio |
| Event ban | 2020-03-08 | 1.00 | Tuscany | https://www.gazzettaufficiale.it/showNewsDetail?id=2513&backTo=archivio&anno=2020&provenienza=archivio |
| Event ban | 2020-03-08 | 1.00 | Sicily | https://www.gazzettaufficiale.it/showNewsDetail?id=2513&backTo=archivio&anno=2020&provenienza=archivio |
| Event ban | 2020-03-08 | 1.00 | Sardinia | https://www.gazzettaufficiale.it/showNewsDetail?id=2513&backTo=archivio&anno=2020&provenienza=archivio |
| Event ban | 2020-03-08 | 1.00 | Puglia (Apulia) | https://www.gazzettaufficiale.it/showNewsDetail?id=2513&backTo=archivio&anno=2020&provenienza=archivio |
| Event ban | 2020-03-08 | 1.00 | Piemonte | https://www.gazzettaufficiale.it/showNewsDetail?id=2513&backTo=archivio&anno=2020&provenienza=archivio |
| Event ban | 2020-03-08 | 1.00 | Molise | https://www.gazzettaufficiale.it/showNewsDetail?id=2513&backTo=archivio&anno=2020&provenienza=archivio |
| Event ban | 2020-03-08 | 1.00 | Liguria | https://www.gazzettaufficiale.it/showNewsDetail?id=2513&backTo=archivio&anno=2020&provenienza=archivio |
| Event ban | 2020-03-08 | 1.00 | Umbria | https://www.gazzettaufficiale.it/showNewsDetail?id=2513&backTo=archivio&anno=2020&provenienza=archivio |
| Event ban | 2020-03-08 | 1.00 | Lazio | https://www.gazzettaufficiale.it/showNewsDetail?id=2513&backTo=archivio&anno=2020&provenienza=archivio |

| | | | | |
|---|---|---|---|---|
| Event ban | 2020-03-08 | 1.00 | Friuli-Venezia Giulia | https://www.gazzettaufficiale.it/showNewsDetail?id=2513&backTo=archivio&anno=2020&provenienza=archivio |
| Event ban | 2020-03-08 | 1.00 | Campania | https://www.gazzettaufficiale.it/showNewsDetail?id=2513&backTo=archivio&anno=2020&provenienza=archivio |
| Event ban | 2020-03-08 | 1.00 | Calabria | https://www.gazzettaufficiale.it/showNewsDetail?id=2513&backTo=archivio&anno=2020&provenienza=archivio |
| Event ban | 2020-03-08 | 1.00 | Basilicata | https://www.gazzettaufficiale.it/showNewsDetail?id=2513&backTo=archivio&anno=2020&provenienza=archivio |
| Event ban | 2020-03-08 | 1.00 | Abruzzo | https://www.gazzettaufficiale.it/showNewsDetail?id=2513&backTo=archivio&anno=2020&provenienza=archivio |
| Event ban | 2020-03-08 | 1.00 | Marche | https://www.gazzettaufficiale.it/showNewsDetail?id=2513&backTo=archivio&anno=2020&provenienza=archivio |
| Event ban | 2020-03-08 | 1.00 | Aosta Valley | https://www.gazzettaufficiale.it/showNewsDetail?id=2513&backTo=archivio&anno=2020&provenienza=archivio |

| **NPI** | **Date** | **Cumulative share** | **Region** | **Source** |
|---|---|---|---|---|
| Gathering ban | 2020-03-02 | 0.32 | Lombardia | https://www.gazzettaufficiale.it/eli/id/2020/03/01/20A01381/sg |
| Gathering ban | 2020-03-02 | 0.32 | Veneto | https://www.gazzettaufficiale.it/eli/id/2020/03/01/20A01381/sg |
| Gathering ban | 2020-03-02 | 0.32 | Emilia-Romagna | https://www.gazzettaufficiale.it/eli/id/2020/03/01/20A01381/sg |
| Gathering ban | 2020-03-08 | 1.00 | Trentino-South Tyrol | https://www.gazzettaufficiale.it/showNewsDetail?id=2513&backTo=archivio&anno=2020&provenienza=archivio |
| Gathering ban | 2020-03-08 | 1.00 | Tuscany | https://www.gazzettaufficiale.it/showNewsDetail?id=2513&backTo=archivio&anno=2020&provenienza=archivio |

| | | | | |
|---|---|---|---|---|
| Gathering ban | 2020-03-08 | 1.00 | Sicily | https://www.gazzettaufficiale.it/showNewsDetail?id=2513&backTo=archivio&anno=2020&provenienza=archivio |
| Gathering ban | 2020-03-08 | 1.00 | Sardinia | https://www.gazzettaufficiale.it/showNewsDetail?id=2513&backTo=archivio&anno=2020&provenienza=archivio |
| Gathering ban | 2020-03-08 | 1.00 | Puglia (Apulia) | https://www.gazzettaufficiale.it/showNewsDetail?id=2513&backTo=archivio&anno=2020&provenienza=archivio |
| Gathering ban | 2020-03-08 | 1.00 | Piemonte | https://www.gazzettaufficiale.it/showNewsDetail?id=2513&backTo=archivio&anno=2020&provenienza=archivio |
| Gathering ban | 2020-03-08 | 1.00 | Molise | https://www.gazzettaufficiale.it/showNewsDetail?id=2513&backTo=archivio&anno=2020&provenienza=archivio |
| Gathering ban | 2020-03-08 | 1.00 | Liguria | https://www.gazzettaufficiale.it/showNewsDetail?id=2513&backTo=archivio&anno=2020&provenienza=archivio |
| Gathering ban | 2020-03-08 | 1.00 | Umbria | https://www.gazzettaufficiale.it/showNewsDetail?id=2513&backTo=archivio&anno=2020&provenienza=archivio |
| Gathering ban | 2020-03-08 | 1.00 | Lazio | https://www.gazzettaufficiale.it/showNewsDetail?id=2513&backTo=archivio&anno=2020&provenienza=archivio |
| Gathering ban | 2020-03-08 | 1.00 | Friuli-Venezia Giulia | https://www.gazzettaufficiale.it/showNewsDetail?id=2513&backTo=archivio&anno=2020&provenienza=archivio |
| Gathering ban | 2020-03-08 | 1.00 | Campania | https://www.gazzettaufficiale.it/showNewsDetail?id=2513&backTo=archivio&anno=2020&provenienza=archivio |
| Gathering ban | 2020-03-08 | 1.00 | Calabria | https://www.gazzettaufficiale.it/showNewsDetail?id=2513&backTo=archivio&anno=2020&provenienza=archivio |
| Gathering ban | 2020-03-08 | 1.00 | Basilicata | https://www.gazzettaufficiale.it/showNewsDetail?id=2513&backTo=archivio&anno=2020&provenienza=archivio |

| | | | | |
|---|---|---|---|---|
| Gathering ban | 2020-03-08 | 1.00 | Abruzzo | https://www.gazzettaufficiale.it/showNewsDetail?id=2513&backTo=archivio&anno=2020&provenienza=archivio |
| Gathering ban | 2020-03-08 | 1.00 | Marche | https://www.gazzettaufficiale.it/showNewsDetail?id=2513&backTo=archivio&anno=2020&provenienza=archivio |
| Gathering ban | 2020-03-08 | 1.00 | Aosta Valley | https://www.gazzettaufficiale.it/showNewsDetail?id=2513&backTo=archivio&anno=2020&provenienza=archivio |

| **NPI** | **Date** | **Cumulative share** | **Region** | **Source** |
|---|---|---|---|---|
| School closure | 2020-03-02 | 0.32 | Lombardia | https://www.gazzettaufficiale.it/eli/id/2020/03/01/20A01381/sg |
| School closure | 2020-03-02 | 0.32 | Veneto | https://www.gazzettaufficiale.it/eli/id/2020/03/01/20A01381/sg |
| School closure | 2020-03-02 | 0.32 | Emilia-Romagna | https://www.gazzettaufficiale.it/eli/id/2020/03/01/20A01381/sg |
| School closure | 2020-03-05 | 1.00 | Trentino-South Tyrol | https://www.gazzettaufficiale.it/eli/id/2020/03/04/20A01475/sg |
| School closure | 2020-03-05 | 1.00 | Tuscany | https://www.gazzettaufficiale.it/eli/id/2020/03/04/20A01475/sg |
| School closure | 2020-03-05 | 1.00 | Sicily | https://www.gazzettaufficiale.it/eli/id/2020/03/04/20A01475/sg |
| School closure | 2020-03-05 | 1.00 | Sardinia | https://www.gazzettaufficiale.it/eli/id/2020/03/04/20A01475/sg |
| School closure | 2020-03-05 | 1.00 | Puglia (Apulia) | https://www.gazzettaufficiale.it/eli/id/2020/03/04/20A01475/sg |
| School closure | 2020-03-05 | 1.00 | Piemonte | https://www.gazzettaufficiale.it/eli/id/2020/03/04/20A01475/sg |

| | | | | |
|---|---|---|---|---|
| School closure | 2020-03-05 | 1.00 | Molise | https://www.gazzettaufficiale.it/eli/id/2020/03/04/20A01475/sg |
| School closure | 2020-03-05 | 1.00 | Liguria | https://www.gazzettaufficiale.it/eli/id/2020/03/04/20A01475/sg |
| School closure | 2020-03-05 | 1.00 | Umbria | https://www.gazzettaufficiale.it/eli/id/2020/03/04/20A01475/sg |
| School closure | 2020-03-05 | 1.00 | Lazio | https://www.gazzettaufficiale.it/eli/id/2020/03/04/20A01475/sg |
| School closure | 2020-03-05 | 1.00 | Friuli-Venezia Giulia | https://www.gazzettaufficiale.it/eli/id/2020/03/04/20A01475/sg |
| School closure | 2020-03-05 | 1.00 | Campania | https://www.gazzettaufficiale.it/eli/id/2020/03/04/20A01475/sg |
| School closure | 2020-03-05 | 1.00 | Calabria | https://www.gazzettaufficiale.it/eli/id/2020/03/04/20A01475/sg |
| School closure | 2020-03-05 | 1.00 | Basilicata | https://www.gazzettaufficiale.it/eli/id/2020/03/04/20A01475/sg |
| School closure | 2020-03-05 | 1.00 | Abruzzo | https://www.gazzettaufficiale.it/eli/id/2020/03/04/20A01475/sg |
| School closure | 2020-03-05 | 1.00 | Marche | https://www.gazzettaufficiale.it/eli/id/2020/03/04/20A01475/sg |
| School closure | 2020-03-05 | 1.00 | Aosta Valley | https://www.gazzettaufficiale.it/eli/id/2020/03/04/20A01475/sg |

| **NPI** | **Date** | **Cumulative share** | **Region** | **Source** |
|---|---|---|---|---|
| Venue closure | 2020-03-12 | 1.00 | Lombardia | https://www.gazzettaufficiale.it/showNewsDetail?id=2532&backTo=archivio&anno=2020&provenienza=archivio |

| | | | | |
|---|---|---|---|---|
| Venue closure | 2020-03-12 | 1.00 | Trentino-South Tyrol | https://www.gazzettaufficiale.it/showNewsDetail?id=2532&backTo=archivio&anno=2020&provenienza=archivio |
| Venue closure | 2020-03-12 | 1.00 | Tuscany | https://www.gazzettaufficiale.it/showNewsDetail?id=2532&backTo=archivio&anno=2020&provenienza=archivio |
| Venue closure | 2020-03-12 | 1.00 | Sicily | https://www.gazzettaufficiale.it/showNewsDetail?id=2532&backTo=archivio&anno=2020&provenienza=archivio |
| Venue closure | 2020-03-12 | 1.00 | Sardinia | https://www.gazzettaufficiale.it/showNewsDetail?id=2532&backTo=archivio&anno=2020&provenienza=archivio |
| Venue closure | 2020-03-12 | 1.00 | Puglia (Apulia) | https://www.gazzettaufficiale.it/showNewsDetail?id=2532&backTo=archivio&anno=2020&provenienza=archivio |
| Venue closure | 2020-03-12 | 1.00 | Piemonte | https://www.gazzettaufficiale.it/showNewsDetail?id=2532&backTo=archivio&anno=2020&provenienza=archivio |
| Venue closure | 2020-03-12 | 1.00 | Molise | https://www.gazzettaufficiale.it/showNewsDetail?id=2532&backTo=archivio&anno=2020&provenienza=archivio |
| Venue closure | 2020-03-12 | 1.00 | Marche | https://www.gazzettaufficiale.it/showNewsDetail?id=2532&backTo=archivio&anno=2020&provenienza=archivio |
| Venue closure | 2020-03-12 | 1.00 | Liguria | https://www.gazzettaufficiale.it/showNewsDetail?id=2532&backTo=archivio&anno=2020&provenienza=archivio |
| Venue closure | 2020-03-12 | 1.00 | Lazio | https://www.gazzettaufficiale.it/showNewsDetail?id=2532&backTo=archivio&anno=2020&provenienza=archivio |
| Venue closure | 2020-03-12 | 1.00 | Friuli-Venezia Giulia | https://www.gazzettaufficiale.it/showNewsDetail?id=2532&backTo=archivio&anno=2020&provenienza=archivio |
| Venue closure | 2020-03-12 | 1.00 | Campania | https://www.gazzettaufficiale.it/showNewsDetail?id=2532&backTo=archivio&anno=2020&provenienza=archivio |
| Venue closure | 2020-03-12 | 1.00 | Calabria | https://www.gazzettaufficiale.it/showNewsDetail?id=2532&backTo=archivio&anno=2020&provenienza=archivio |

| | | | | |
|---|---|---|---|---|
| Venue closure | 2020-03-12 | 1.00 | Basilicata | https://www.gazzettaufficiale.it/showNewsDetail?id=2532&backTo=archivio&anno=2020&provenienza=archivio |
| Venue closure | 2020-03-12 | 1.00 | Abruzzo | https://www.gazzettaufficiale.it/showNewsDetail?id=2532&backTo=archivio&anno=2020&provenienza=archivio |
| Venue closure | 2020-03-12 | 1.00 | Emilia-Romagna | https://www.gazzettaufficiale.it/showNewsDetail?id=2532&backTo=archivio&anno=2020&provenienza=archivio |
| Venue closure | 2020-03-12 | 1.00 | Veneto | https://www.gazzettaufficiale.it/showNewsDetail?id=2532&backTo=archivio&anno=2020&provenienza=archivio |
| Venue closure | 2020-03-12 | 1.00 | Umbria | https://www.gazzettaufficiale.it/showNewsDetail?id=2532&backTo=archivio&anno=2020&provenienza=archivio |
| Venue closure | 2020-03-12 | 1.00 | Aosta Valley | https://www.gazzettaufficiale.it/showNewsDetail?id=2532&backTo=archivio&anno=2020&provenienza=archivio |

| **NPI** | **Date** | **Cumulative share** | **Region** | **Source** |
|---|---|---|---|---|
| Lockdown | 2020-03-12 | 1.00 | Lombardia | https://www.gazzettaufficiale.it/showNewsDetail?id=2532&backTo=archivio&anno=2020&provenienza=archivio |
| Lockdown | 2020-03-12 | 1.00 | Trentino-South Tyrol | https://www.gazzettaufficiale.it/showNewsDetail?id=2532&backTo=archivio&anno=2020&provenienza=archivio |
| Lockdown | 2020-03-12 | 1.00 | Tuscany | https://www.gazzettaufficiale.it/showNewsDetail?id=2532&backTo=archivio&anno=2020&provenienza=archivio |
| Lockdown | 2020-03-12 | 1.00 | Sicily | https://www.gazzettaufficiale.it/showNewsDetail?id=2532&backTo=archivio&anno=2020&provenienza=archivio |
| Lockdown | 2020-03-12 | 1.00 | Sardinia | https://www.gazzettaufficiale.it/showNewsDetail?id=2532&backTo=archivio&anno=2020&provenienza=archivio |
| Lockdown | 2020-03-12 | 1.00 | Puglia (Apulia) | https://www.gazzettaufficiale.it/showNewsDetail?id=2532&backTo=archivio&anno=2020&provenienza=archivio |

| | | | | |
|---|---|---|---|---|
| Lockdown | 2020-03-12 | 1.00 | Piemonte | https://www.gazzettaufficiale.it/showNewsDetail?id=2532&backTo=archivio&anno=2020&provenienza=archivio |
| Lockdown | 2020-03-12 | 1.00 | Molise | https://www.gazzettaufficiale.it/showNewsDetail?id=2532&backTo=archivio&anno=2020&provenienza=archivio |
| Lockdown | 2020-03-12 | 1.00 | Marche | https://www.gazzettaufficiale.it/showNewsDetail?id=2532&backTo=archivio&anno=2020&provenienza=archivio |
| Lockdown | 2020-03-12 | 1.00 | Liguria | https://www.gazzettaufficiale.it/showNewsDetail?id=2532&backTo=archivio&anno=2020&provenienza=archivio |
| Lockdown | 2020-03-12 | 1.00 | Lazio | https://www.gazzettaufficiale.it/showNewsDetail?id=2532&backTo=archivio&anno=2020&provenienza=archivio |
| Lockdown | 2020-03-12 | 1.00 | Friuli-Venezia Giulia | https://www.gazzettaufficiale.it/showNewsDetail?id=2532&backTo=archivio&anno=2020&provenienza=archivio |
| Lockdown | 2020-03-12 | 1.00 | Campania | https://www.gazzettaufficiale.it/showNewsDetail?id=2532&backTo=archivio&anno=2020&provenienza=archivio |
| Lockdown | 2020-03-12 | 1.00 | Calabria | https://www.gazzettaufficiale.it/showNewsDetail?id=2532&backTo=archivio&anno=2020&provenienza=archivio |
| Lockdown | 2020-03-12 | 1.00 | Basilicata | https://www.gazzettaufficiale.it/showNewsDetail?id=2532&backTo=archivio&anno=2020&provenienza=archivio |
| Lockdown | 2020-03-12 | 1.00 | Abruzzo | https://www.gazzettaufficiale.it/showNewsDetail?id=2532&backTo=archivio&anno=2020&provenienza=archivio |
| Lockdown | 2020-03-12 | 1.00 | Emilia-Romagna | https://www.gazzettaufficiale.it/showNewsDetail?id=2532&backTo=archivio&anno=2020&provenienza=archivio |
| Lockdown | 2020-03-12 | 1.00 | Veneto | https://www.gazzettaufficiale.it/showNewsDetail?id=2532&backTo=archivio&anno=2020&provenienza=archivio |
| Lockdown | 2020-03-12 | 1.00 | Umbria | https://www.gazzettaufficiale.it/showNewsDetail?id=2532&backTo=archivio&anno=2020&provenienza=archivio |

| | | | | |
|---|---|---|---|---|
| Lockdown | 2020-03-12 | 1.00 | Aosta Valley | https://www.gazzettaufficiale.it/showNewsDetail?id=2532&backTo=archivio&anno=2020&provenienza=archivio |

| NPI | Date | Cumulative share | Region | Source |
|---|---|---|---|---|
| Work ban | 2020-03-22 | 1.00 | Lombardia | https://www.gazzettaufficiale.it/showNewsDetail?id=2545&backTo=archivio&anno=2020&provenienza=archivio |
| Work ban | 2020-03-22 | 1.00 | Trentino-South Tyrol | https://www.gazzettaufficiale.it/showNewsDetail?id=2545&backTo=archivio&anno=2020&provenienza=archivio |
| Work ban | 2020-03-22 | 1.00 | Tuscany | https://www.gazzettaufficiale.it/showNewsDetail?id=2545&backTo=archivio&anno=2020&provenienza=archivio |
| Work ban | 2020-03-22 | 1.00 | Sicily | https://www.gazzettaufficiale.it/showNewsDetail?id=2545&backTo=archivio&anno=2020&provenienza=archivio |
| Work ban | 2020-03-22 | 1.00 | Sardinia | https://www.gazzettaufficiale.it/showNewsDetail?id=2545&backTo=archivio&anno=2020&provenienza=archivio |
| Work ban | 2020-03-22 | 1.00 | Puglia (Apulia) | https://www.gazzettaufficiale.it/showNewsDetail?id=2545&backTo=archivio&anno=2020&provenienza=archivio |
| Work ban | 2020-03-22 | 1.00 | Piemonte | https://www.gazzettaufficiale.it/showNewsDetail?id=2545&backTo=archivio&anno=2020&provenienza=archivio |
| Work ban | 2020-03-22 | 1.00 | Molise | https://www.gazzettaufficiale.it/showNewsDetail?id=2545&backTo=archivio&anno=2020&provenienza=archivio |
| Work ban | 2020-03-22 | 1.00 | Marche | https://www.gazzettaufficiale.it/showNewsDetail?id=2545&backTo=archivio&anno=2020&provenienza=archivio |
| Work ban | 2020-03-22 | 1.00 | Liguria | https://www.gazzettaufficiale.it/showNewsDetail?id=2545&backTo=archivio&anno=2020&provenienza=archivio |
| Work ban | 2020-03-22 | 1.00 | Lazio | https://www.gazzettaufficiale.it/showNewsDetail?id=2545&backTo=archivio&anno=2020&provenienza=archivio |

| | | | | |
|---|---|---|---|---|
| Work ban | 2020-03-22 | 1.00 | Friuli-Venezia Giulia | https://www.gazzettaufficiale.it/showNewsDetail?id=2545&backTo=archivio&anno=2020&provenienza=archivio |
| Work ban | 2020-03-22 | 1.00 | Campania | https://www.gazzettaufficiale.it/showNewsDetail?id=2545&backTo=archivio&anno=2020&provenienza=archivio |
| Work ban | 2020-03-22 | 1.00 | Calabria | https://www.gazzettaufficiale.it/showNewsDetail?id=2545&backTo=archivio&anno=2020&provenienza=archivio |
| Work ban | 2020-03-22 | 1.00 | Basilicata | https://www.gazzettaufficiale.it/showNewsDetail?id=2545&backTo=archivio&anno=2020&provenienza=archivio |
| Work ban | 2020-03-22 | 1.00 | Abruzzo | https://www.gazzettaufficiale.it/showNewsDetail?id=2545&backTo=archivio&anno=2020&provenienza=archivio |
| Work ban | 2020-03-22 | 1.00 | Emilia-Romagna | https://www.gazzettaufficiale.it/showNewsDetail?id=2545&backTo=archivio&anno=2020&provenienza=archivio |
| Work ban | 2020-03-22 | 1.00 | Veneto | https://www.gazzettaufficiale.it/showNewsDetail?id=2545&backTo=archivio&anno=2020&provenienza=archivio |
| Work ban | 2020-03-22 | 1.00 | Umbria | https://www.gazzettaufficiale.it/showNewsDetail?id=2545&backTo=archivio&anno=2020&provenienza=archivio |
| Work ban | 2020-03-22 | 1.00 | Aosta Valley | https://www.gazzettaufficiale.it/showNewsDetail?id=2545&backTo=archivio&anno=2020&provenienza=archivio |

**Table 12.** Sources for policies implemented across different Italian regions

| NPI | Date | Cumulative share | Region | Source |
|---|---|---|---|---|
| Event ban | 2020-02-28 | 1.00 | Zurich | https://www.admin.ch/gov/it/pagina-iniziale/documentazione/comunicati-stampa.msg-id-78289.html |

| | | | | |
|---|---|---|---|---|
| Event ban | 2020-02-28 | 1.00 | Neuchâtel | https://www.admin.ch/gov/it/pagina-iniziale/documentazione/comunicati-stampa.msg-id-78289.html |
| Event ban | 2020-02-28 | 1.00 | Valais | https://www.admin.ch/gov/it/pagina-iniziale/documentazione/comunicati-stampa.msg-id-78289.html |
| Event ban | 2020-02-28 | 1.00 | Vaud | https://www.admin.ch/gov/it/pagina-iniziale/documentazione/comunicati-stampa.msg-id-78289.html |
| Event ban | 2020-02-28 | 1.00 | Ticino | https://www.admin.ch/gov/it/pagina-iniziale/documentazione/comunicati-stampa.msg-id-78289.html |
| Event ban | 2020-02-28 | 1.00 | Thurgau | https://www.admin.ch/gov/it/pagina-iniziale/documentazione/comunicati-stampa.msg-id-78289.html |
| Event ban | 2020-02-28 | 1.00 | Aargau | https://www.admin.ch/gov/it/pagina-iniziale/documentazione/comunicati-stampa.msg-id-78289.html |
| Event ban | 2020-02-28 | 1.00 | Graubünden | https://www.admin.ch/gov/it/pagina-iniziale/documentazione/comunicati-stampa.msg-id-78289.html |
| Event ban | 2020-02-28 | 1.00 | St. Gallen | https://www.admin.ch/gov/it/pagina-iniziale/documentazione/comunicati-stampa.msg-id-78289.html |
| Event ban | 2020-02-28 | 1.00 | Appenzell Innerrhoden | https://www.admin.ch/gov/it/pagina-iniziale/documentazione/comunicati-stampa.msg-id-78289.html |
| Event ban | 2020-02-28 | 1.00 | Appenzell Ausserrhoden | https://www.admin.ch/gov/it/pagina-iniziale/documentazione/comunicati-stampa.msg-id-78289.html |
| Event ban | 2020-02-28 | 1.00 | Schaffhausen | https://www.admin.ch/gov/it/pagina-iniziale/documentazione/comunicati-stampa.msg-id-78289.html |
| Event ban | 2020-02-28 | 1.00 | Basel-Landschaft | https://www.admin.ch/gov/it/pagina-iniziale/documentazione/comunicati-stampa.msg-id-78289.html |
| Event ban | 2020-02-28 | 1.00 | Basel-Stadt | https://www.admin.ch/gov/it/pagina-iniziale/documentazione/comunicati-stampa.msg-id-78289.html |

| NPI | Date | Cumulative share | Region | Source |
|---|---|---|---|---|
| Event ban | 2020-02-28 | 1.00 | Solothurn | https://www.admin.ch/gov/it/pagina-iniziale/documentazione/comunicati-stampa.msg-id-78289.html |
| Event ban | 2020-02-28 | 1.00 | Fribourg | https://www.admin.ch/gov/it/pagina-iniziale/documentazione/comunicati-stampa.msg-id-78289.html |
| Event ban | 2020-02-28 | 1.00 | Zug | https://www.admin.ch/gov/it/pagina-iniziale/documentazione/comunicati-stampa.msg-id-78289.html |
| Event ban | 2020-02-28 | 1.00 | Glarus | https://www.admin.ch/gov/it/pagina-iniziale/documentazione/comunicati-stampa.msg-id-78289.html |
| Event ban | 2020-02-28 | 1.00 | Nidwalden | https://www.admin.ch/gov/it/pagina-iniziale/documentazione/comunicati-stampa.msg-id-78289.html |
| Event ban | 2020-02-28 | 1.00 | Obwalden | https://www.admin.ch/gov/it/pagina-iniziale/documentazione/comunicati-stampa.msg-id-78289.html |
| Event ban | 2020-02-28 | 1.00 | Schwyz | https://www.admin.ch/gov/it/pagina-iniziale/documentazione/comunicati-stampa.msg-id-78289.html |
| Event ban | 2020-02-28 | 1.00 | Uri | https://www.admin.ch/gov/it/pagina-iniziale/documentazione/comunicati-stampa.msg-id-78289.html |
| Event ban | 2020-02-28 | 1.00 | Lucerne | https://www.admin.ch/gov/it/pagina-iniziale/documentazione/comunicati-stampa.msg-id-78289.html |
| Event ban | 2020-02-28 | 1.00 | Bern | https://www.admin.ch/gov/it/pagina-iniziale/documentazione/comunicati-stampa.msg-id-78289.html |
| Event ban | 2020-02-28 | 1.00 | Geneva | https://www.admin.ch/gov/it/pagina-iniziale/documentazione/comunicati-stampa.msg-id-78289.html |
| Event ban | 2020-02-28 | 1.00 | Jura | https://www.admin.ch/gov/it/pagina-iniziale/documentazione/comunicati-stampa.msg-id-78289.html |

| | | | | |
|---|---|---|---|---|
| Gathering ban | 2020-03-16 | 0.02 | Neuchâtel | https://www.ne.ch/medias/Pages/20200315-mesures-urgence-lutte-covid19-canton-de-neuchatel.aspx |
| Gathering ban | 2020-03-18 | 0.03 | Jura | https://www.jura.ch/CHA/SIC/Centre-medias/Communiques-2020/COVID-19-etat-de-necessite-decrete-et-interdiction-des-rassemblements-de-plus-de-5-personnes.html |
| Gathering ban | 2020-03-20 | 1.00 | Valais | https://www.admin.ch/gov/it/pagina-iniziale/documentazione/comunicati-stampa/comunicati-stampa-consiglio-federale.msg-id-78513.html |
| Gathering ban | 2020-03-20 | 1.00 | Vaud | https://www.admin.ch/gov/it/pagina-iniziale/documentazione/comunicati-stampa/comunicati-stampa-consiglio-federale.msg-id-78513.html |
| Gathering ban | 2020-03-20 | 1.00 | Ticino | https://www.admin.ch/gov/it/pagina-iniziale/documentazione/comunicati-stampa/comunicati-stampa-consiglio-federale.msg-id-78513.html |
| Gathering ban | 2020-03-20 | 1.00 | Thurgau | https://www.admin.ch/gov/it/pagina-iniziale/documentazione/comunicati-stampa/comunicati-stampa-consiglio-federale.msg-id-78513.html |
| Gathering ban | 2020-03-20 | 1.00 | Aargau | https://www.admin.ch/gov/it/pagina-iniziale/documentazione/comunicati-stampa/comunicati-stampa-consiglio-federale.msg-id-78513.html |
| Gathering ban | 2020-03-20 | 1.00 | Graubünden | https://www.admin.ch/gov/it/pagina-iniziale/documentazione/comunicati-stampa/comunicati-stampa-consiglio-federale.msg-id-78513.html |
| Gathering ban | 2020-03-20 | 1.00 | St. Gallen | https://www.admin.ch/gov/it/pagina-iniziale/documentazione/comunicati-stampa/comunicati-stampa-consiglio-federale.msg-id-78513.html |
| Gathering ban | 2020-03-20 | 1.00 | Appenzell Innerrhoden | https://www.admin.ch/gov/it/pagina-iniziale/documentazione/comunicati-stampa/comunicati-stampa-consiglio-federale.msg-id-78513.html |
| Gathering ban | 2020-03-20 | 1.00 | Appenzell Ausserrhoden | https://www.admin.ch/gov/it/pagina-iniziale/documentazione/comunicati-stampa/comunicati-stampa-consiglio-federale.msg-id-78513.html |

| | | | | |
|---|---|---|---|---|
| Gathering ban | 2020-03-20 | 1.00 | Schaffhausen | https://www.admin.ch/gov/it/pagina-iniziale/documentazione/comunicati-stampa/comunicati-stampa-consiglio-federale.msg-id-78513.html |
| Gathering ban | 2020-03-20 | 1.00 | Zurich | https://www.admin.ch/gov/it/pagina-iniziale/documentazione/comunicati-stampa/comunicati-stampa-consiglio-federale.msg-id-78513.html |
| Gathering ban | 2020-03-20 | 1.00 | Basel-Stadt | https://www.admin.ch/gov/it/pagina-iniziale/documentazione/comunicati-stampa/comunicati-stampa-consiglio-federale.msg-id-78513.html |
| Gathering ban | 2020-03-20 | 1.00 | Solothurn | https://www.admin.ch/gov/it/pagina-iniziale/documentazione/comunicati-stampa/comunicati-stampa-consiglio-federale.msg-id-78513.html |
| Gathering ban | 2020-03-20 | 1.00 | Fribourg | https://www.admin.ch/gov/it/pagina-iniziale/documentazione/comunicati-stampa/comunicati-stampa-consiglio-federale.msg-id-78513.html |
| Gathering ban | 2020-03-20 | 1.00 | Zug | https://www.admin.ch/gov/it/pagina-iniziale/documentazione/comunicati-stampa/comunicati-stampa-consiglio-federale.msg-id-78513.html |
| Gathering ban | 2020-03-20 | 1.00 | Glarus | https://www.admin.ch/gov/it/pagina-iniziale/documentazione/comunicati-stampa/comunicati-stampa-consiglio-federale.msg-id-78513.html |
| Gathering ban | 2020-03-20 | 1.00 | Nidwalden | https://www.admin.ch/gov/it/pagina-iniziale/documentazione/comunicati-stampa/comunicati-stampa-consiglio-federale.msg-id-78513.html |
| Gathering ban | 2020-03-20 | 1.00 | Obwalden | https://www.admin.ch/gov/it/pagina-iniziale/documentazione/comunicati-stampa/comunicati-stampa-consiglio-federale.msg-id-78513.html |
| Gathering ban | 2020-03-20 | 1.00 | Schwyz | https://www.admin.ch/gov/it/pagina-iniziale/documentazione/comunicati-stampa/comunicati-stampa-consiglio-federale.msg-id-78513.html |
| Gathering ban | 2020-03-20 | 1.00 | Uri | https://www.admin.ch/gov/it/pagina-iniziale/documentazione/comunicati-stampa/comunicati-stampa-consiglio-federale.msg-id-78513.html |
| Gathering ban | 2020-03-20 | 1.00 | Lucerne | https://www.admin.ch/gov/it/pagina-iniziale/documentazione/comunicati-stampa/comunicati-stampa-consiglio-federale.msg-id-78513.html |
| Gathering ban | 2020-03-20 | 1.00 | Bern | https://www.admin.ch/gov/it/pagina-iniziale/documentazione/comunicati-stampa/comunicati-stampa-consiglio-federale.msg-id-78513.html |

| | | | | |
|---|---|---|---|---|
| Gathering ban | 2020-03-20 | 1.00 | Geneva | https://www.admin.ch/gov/it/pagina-iniziale/documentazione/comunicati-stampa/comunicati-stampa-consiglio-federale.msg-id-78513.html |
| Gathering ban | 2020-03-20 | 1.00 | Basel-Landschaft | https://www.admin.ch/gov/it/pagina-iniziale/documentazione/comunicati-stampa/comunicati-stampa-consiglio-federale.msg-id-78513.html |

| **NPI** | **Date** | **Cumulative share** | **Region** | **Source** |
|---|---|---|---|---|
| School closure | 2020-03-16 | 1.00 | Zurich | https://www.admin.ch/gov/it/pagina-iniziale/documentazione/comunicati-stampa/comunicati-stampa-consiglio-federale.html?dyn_startDate=01.01.2015 |
| School closure | 2020-03-16 | 1.00 | Neuchâtel | https://www.admin.ch/gov/it/pagina-iniziale/documentazione/comunicati-stampa/comunicati-stampa-consiglio-federale.html?dyn_startDate=01.01.2015 |
| School closure | 2020-03-16 | 1.00 | Valais | https://www.admin.ch/gov/it/pagina-iniziale/documentazione/comunicati-stampa/comunicati-stampa-consiglio-federale.html?dyn_startDate=01.01.2015 |
| School closure | 2020-03-16 | 1.00 | Vaud | https://www.admin.ch/gov/it/pagina-iniziale/documentazione/comunicati-stampa/comunicati-stampa-consiglio-federale.html?dyn_startDate=01.01.2015 |
| School closure | 2020-03-16 | 1.00 | Ticino | https://www.admin.ch/gov/it/pagina-iniziale/documentazione/comunicati-stampa/comunicati-stampa-consiglio-federale.html?dyn_startDate=01.01.2015 |
| School closure | 2020-03-16 | 1.00 | Thurgau | https://www.admin.ch/gov/it/pagina-iniziale/documentazione/comunicati-stampa/comunicati-stampa-consiglio-federale.html?dyn_startDate=01.01.2015 |

| | | | | |
|---|---|---|---|---|
| School closure | 2020-03-16 | 1.00 | Aargau | https://www.admin.ch/gov/it/pagina-iniziale/documentazione/comunicati-stampa/comunicati-stampa-consiglio-federale.html?dyn_startDate=01.01.2015 |
| School closure | 2020-03-16 | 1.00 | Graubünden | https://www.admin.ch/gov/it/pagina-iniziale/documentazione/comunicati-stampa/comunicati-stampa-consiglio-federale.html?dyn_startDate=01.01.2015 |
| School closure | 2020-03-16 | 1.00 | St. Gallen | https://www.admin.ch/gov/it/pagina-iniziale/documentazione/comunicati-stampa/comunicati-stampa-consiglio-federale.html?dyn_startDate=01.01.2015 |
| School closure | 2020-03-16 | 1.00 | Appenzell Innerrhoden | https://www.admin.ch/gov/it/pagina-iniziale/documentazione/comunicati-stampa/comunicati-stampa-consiglio-federale.html?dyn_startDate=01.01.2015 |
| School closure | 2020-03-16 | 1.00 | Appenzell Ausserrhoden | https://www.admin.ch/gov/it/pagina-iniziale/documentazione/comunicati-stampa/comunicati-stampa-consiglio-federale.html?dyn_startDate=01.01.2015 |
| School closure | 2020-03-16 | 1.00 | Schaffhausen | https://www.admin.ch/gov/it/pagina-iniziale/documentazione/comunicati-stampa/comunicati-stampa-consiglio-federale.html?dyn_startDate=01.01.2015 |
| School closure | 2020-03-16 | 1.00 | Basel-Landschaft | https://www.admin.ch/gov/it/pagina-iniziale/documentazione/comunicati-stampa/comunicati-stampa-consiglio-federale.html?dyn_startDate=01.01.2015 |
| School closure | 2020-03-16 | 1.00 | Basel-Stadt | https://www.admin.ch/gov/it/pagina-iniziale/documentazione/comunicati-stampa/comunicati-stampa-consiglio-federale.html?dyn_startDate=01.01.2015 |

| | | | | |
|---|---|---|---|---|
| School closure | 2020-03-16 | 1.00 | Solothurn | https://www.admin.ch/gov/it/pagina-iniziale/documentazione/comunicati-stampa/comunicati-stampa-consiglio-federale.html?dyn_startDate=01.01.2015 |
| School closure | 2020-03-16 | 1.00 | Fribourg | https://www.admin.ch/gov/it/pagina-iniziale/documentazione/comunicati-stampa/comunicati-stampa-consiglio-federale.html?dyn_startDate=01.01.2015 |
| School closure | 2020-03-16 | 1.00 | Zug | https://www.admin.ch/gov/it/pagina-iniziale/documentazione/comunicati-stampa/comunicati-stampa-consiglio-federale.html?dyn_startDate=01.01.2015 |
| School closure | 2020-03-16 | 1.00 | Glarus | https://www.admin.ch/gov/it/pagina-iniziale/documentazione/comunicati-stampa/comunicati-stampa-consiglio-federale.html?dyn_startDate=01.01.2015 |
| School closure | 2020-03-16 | 1.00 | Nidwalden | https://www.admin.ch/gov/it/pagina-iniziale/documentazione/comunicati-stampa/comunicati-stampa-consiglio-federale.html?dyn_startDate=01.01.2015 |
| School closure | 2020-03-16 | 1.00 | Obwalden | https://www.admin.ch/gov/it/pagina-iniziale/documentazione/comunicati-stampa/comunicati-stampa-consiglio-federale.html?dyn_startDate=01.01.2015 |
| School closure | 2020-03-16 | 1.00 | Schwyz | https://www.admin.ch/gov/it/pagina-iniziale/documentazione/comunicati-stampa/comunicati-stampa-consiglio-federale.html?dyn_startDate=01.01.2015 |
| School closure | 2020-03-16 | 1.00 | Uri | https://www.admin.ch/gov/it/pagina-iniziale/documentazione/comunicati-stampa/comunicati-stampa-consiglio-federale.html?dyn_startDate=01.01.2015 |

| | | | | |
|---|---|---|---|---|
| School closure | 2020-03-16 | 1.00 | Lucerne | https://www.admin.ch/gov/it/pagina-iniziale/documentazione/comunicati-stampa/comunicati-stampa-consiglio-federale.html?dyn_startDate=01.01.2015 |
| School closure | 2020-03-16 | 1.00 | Bern | https://www.admin.ch/gov/it/pagina-iniziale/documentazione/comunicati-stampa/comunicati-stampa-consiglio-federale.html?dyn_startDate=01.01.2015 |
| School closure | 2020-03-16 | 1.00 | Geneva | https://www.admin.ch/gov/it/pagina-iniziale/documentazione/comunicati-stampa/comunicati-stampa-consiglio-federale.html?dyn_startDate=01.01.2015 |
| School closure | 2020-03-16 | 1.00 | Jura | https://www.admin.ch/gov/it/pagina-iniziale/documentazione/comunicati-stampa/comunicati-stampa-consiglio-federale.html?dyn_startDate=01.01.2015 |

| **NPI** | **Date** | **Cumulative share** | **Region** | **Source** |
|---|---|---|---|---|
| Venue closure | 2020-03-14 | 0.04 | Ticino | https://www4.ti.ch/dss/dsp/covid19/home/ |
| Venue closure | 2020-03-16 | 0.09 | Jura | https://www.jura.ch/CHA/SIC/Centre-medias/Communiques-2020/COVID-19-le-canton-du-Jura-prend-des-mesures-supplementaires-pour-proteger-la-population-et-enrayer-la-propagation-du-coronaviru.html |
| Venue closure | 2020-03-16 | 0.09 | Neuchâtel | https://www.ne.ch/medias/Pages/20200315-mesures-urgence-lutte-covid19-canton-de-neuchatel.aspx |
| Venue closure | 2020-03-16 | 0.09 | Graubünden | https://www.kantonsamtsblatt.gr.ch/it/efuc/00.045.026/publikation/ |
| Venue closure | 2020-03-17 | 1.00 | Valais | https://www.admin.ch/gov/it/pagina-iniziale/documentazione/comunicati-stampa/comunicati-stampa-consiglio-federale.html?dyn_startDate=01.01.2015 |

| | | | | |
|---|---|---|---|---|
| Venue closure | 2020-03-17 | 1.00 | Vaud | https://www.admin.ch/gov/it/pagina-iniziale/documentazione/comunicati-stampa/comunicati-stampa-consiglio-federale.html?dyn_startDate=01.01.2015 |
| Venue closure | 2020-03-17 | 1.00 | Thurgau | https://www.admin.ch/gov/it/pagina-iniziale/documentazione/comunicati-stampa/comunicati-stampa-consiglio-federale.html?dyn_startDate=01.01.2015 |
| Venue closure | 2020-03-17 | 1.00 | Aargau | https://www.admin.ch/gov/it/pagina-iniziale/documentazione/comunicati-stampa/comunicati-stampa-consiglio-federale.html?dyn_startDate=01.01.2015 |
| Venue closure | 2020-03-17 | 1.00 | St. Gallen | https://www.admin.ch/gov/it/pagina-iniziale/documentazione/comunicati-stampa/comunicati-stampa-consiglio-federale.html?dyn_startDate=01.01.2015 |
| Venue closure | 2020-03-17 | 1.00 | Appenzell Innerrhoden | https://www.admin.ch/gov/it/pagina-iniziale/documentazione/comunicati-stampa/comunicati-stampa-consiglio-federale.html?dyn_startDate=01.01.2015 |
| Venue closure | 2020-03-17 | 1.00 | Appenzell Ausserrhoden | https://www.admin.ch/gov/it/pagina-iniziale/documentazione/comunicati-stampa/comunicati-stampa-consiglio-federale.html?dyn_startDate=01.01.2015 |
| Venue closure | 2020-03-17 | 1.00 | Schaffhausen | https://www.admin.ch/gov/it/pagina-iniziale/documentazione/comunicati-stampa/comunicati-stampa-consiglio-federale.html?dyn_startDate=01.01.2015 |
| Venue closure | 2020-03-17 | 1.00 | Zurich | https://www.admin.ch/gov/it/pagina-iniziale/documentazione/comunicati-stampa/comunicati-stampa-consiglio-federale.html?dyn_startDate=01.01.2015 |

| | | | | |
|---|---|---|---|---|
| Venue closure | 2020-03-17 | 1.00 | Basel-Stadt | https://www.admin.ch/gov/it/pagina-iniziale/documentazione/comunicati-stampa/comunicati-stampa-consiglio-federale.html?dyn_startDate=01.01.2015 |
| Venue closure | 2020-03-17 | 1.00 | Solothurn | https://www.admin.ch/gov/it/pagina-iniziale/documentazione/comunicati-stampa/comunicati-stampa-consiglio-federale.html?dyn_startDate=01.01.2015 |
| Venue closure | 2020-03-17 | 1.00 | Fribourg | https://www.admin.ch/gov/it/pagina-iniziale/documentazione/comunicati-stampa/comunicati-stampa-consiglio-federale.html?dyn_startDate=01.01.2015 |
| Venue closure | 2020-03-17 | 1.00 | Zug | https://www.admin.ch/gov/it/pagina-iniziale/documentazione/comunicati-stampa/comunicati-stampa-consiglio-federale.html?dyn_startDate=01.01.2015 |
| Venue closure | 2020-03-17 | 1.00 | Glarus | https://www.admin.ch/gov/it/pagina-iniziale/documentazione/comunicati-stampa/comunicati-stampa-consiglio-federale.html?dyn_startDate=01.01.2015 |
| Venue closure | 2020-03-17 | 1.00 | Nidwalden | https://www.admin.ch/gov/it/pagina-iniziale/documentazione/comunicati-stampa/comunicati-stampa-consiglio-federale.html?dyn_startDate=01.01.2015 |
| Venue closure | 2020-03-17 | 1.00 | Obwalden | https://www.admin.ch/gov/it/pagina-iniziale/documentazione/comunicati-stampa/comunicati-stampa-consiglio-federale.html?dyn_startDate=01.01.2015 |
| Venue closure | 2020-03-17 | 1.00 | Schwyz | https://www.admin.ch/gov/it/pagina-iniziale/documentazione/comunicati-stampa/comunicati-stampa-consiglio-federale.html?dyn_startDate=01.01.2015 |

| | | | | |
|---|---|---|---|---|
| Venue closure | 2020-03-17 | 1.00 | Uri | https://www.admin.ch/gov/it/pagina-iniziale/documentazione/comunicati-stampa/comunicati-stampa-consiglio-federale.html?dyn_startDate=01.01.2015 |
| Venue closure | 2020-03-17 | 1.00 | Lucerne | https://www.admin.ch/gov/it/pagina-iniziale/documentazione/comunicati-stampa/comunicati-stampa-consiglio-federale.html?dyn_startDate=01.01.2015 |
| Venue closure | 2020-03-17 | 1.00 | Bern | https://www.admin.ch/gov/it/pagina-iniziale/documentazione/comunicati-stampa/comunicati-stampa-consiglio-federale.html?dyn_startDate=01.01.2015 |
| Venue closure | 2020-03-17 | 1.00 | Geneva | https://www.admin.ch/gov/it/pagina-iniziale/documentazione/comunicati-stampa/comunicati-stampa-consiglio-federale.html?dyn_startDate=01.01.2015 |
| Venue closure | 2020-03-17 | 1.00 | Basel-Landschaft | https://www.admin.ch/gov/it/pagina-iniziale/documentazione/comunicati-stampa/comunicati-stampa-consiglio-federale.html?dyn_startDate=01.01.2015 |
| **NPI** | **Date** | **Cumulative share** | **Region** | **Source** |
| **NPI** | **Date** | **Cumulative share** | **Region** | **Source** |
| Work ban | 2020-03-14 | 0.04 | Ticino | https://www4.ti.ch/dss/dsp/covid19/home/ |

**Table 13.** Sources for policies implemented across different Swiss regions
